# Genomic Epidemiology of SARS-CoV-2 in Esteio, Rio Grande do Sul, Brazil

**DOI:** 10.1101/2021.01.21.21249906

**Authors:** Vinícius Bonetti Franceschi, Gabriel Dickin Caldana, Amanda de Menezes Mayer, Gabriela Bettella Cybis, Carla Andretta Moreira Neves, Patrícia Aline Gröhs Ferrareze, Meriane Demoliner, Paula Rodrigues de Almeida, Juliana Schons Gularte, Alana Witt Hansen, Matheus Nunes Weber, Juliane Deise Fleck, Ricardo Ariel Zimerman, Lívia Kmetzsch, Fernando Rosado Spilki, Claudia Elizabeth Thompson

**Affiliations:** Center of Biotechnology, Graduate Program in Cell and Molecular Biology (PPGBCM), Universidade Federal do Rio Grande do Sul (UFRGS), Porto Alegre, RS, Brazil; Graduate Program in Health Sciences, Universidade Federal de Ciências da Saúde de Porto Alegre (UFCSPA), Porto Alegre, RS, Brazil; Department of Statistics, Universidade Federal do Rio Grande do Sul (UFRGS), Porto Alegre, RS, Brazil; Molecular Microbiology Laboratory, Universidade Feevale, Novo Hamburgo, RS, Brazil; Irmandade Santa Casa de Misericórdia de Porto Alegre, Porto Alegre, RS, Brazil; Department of Pharmacosciences, Universidade Federal de Ciências da Saúde de Porto Alegre (UFCSPA), Porto Alegre, RS, Brazil

**Keywords:** Covid-19, Severe acute respiratory syndrome coronavirus 2, Infectious Diseases, Sequencing, Molecular Epidemiology

## Abstract

Brazil is the third country most affected by Covid-19 pandemic. In spite of this, viral evolution in municipality resolution is poorly understood in Brazil and it is crucial to understand the epidemiology of viral spread. We identified four main circulating lineages in Esteio (Southern Brazil) and their relationship with global, national and regional lineages using phylogenetics and phylodynamics inferences from 21 SARS-CoV-2 genome sequences. We provided a comprehensive view of viral mutations from a time- and age-representative sampling from May to October 2020, in Esteio (RS, Brazil), highlighting two frequent mutations in Spike glycoprotein (D614G and V1176F), an emergent mutation (E484K) in Spike Receptor Binding Domain (RBD) characteristic of the South African lineage B.1.351, and the adjacent replacement of 2 amino acids in Nucleocapsid phosphoprotein (R203K and G204R). A significant viral diversity was evidenced with the identification of 80 different SNPs. The E484K replacement was found in two genomes (9.5%) from samples obtained in mid-October, which is to our best knowledge the earliest description of E484K harboring SARS-CoV-2 in South Brazil. This mutation identified in a small municipality from the RS state demonstrates that it was probably widely distributed in the Brazilian territory, but went unnoticed so far by the lack of genomic surveillance in Brazil. The introduction of E484K mutants shows temporal correlation with later increases in new cases in our state. Importantly, since it has been associated with immune evasion and enhanced interaction with hACE-2, lineages containing this substitution must be the subject of intense surveillance. Our date demonstrates multiple introductions of the most prevalent lineages (B.1.1.33 and B.1.1.248) and the major role of community transmission in viral spreading and the establishment of Brazilian lineages. This represents an important contribution to the epidemiology of SARS-CoV-2.

## Introduction

In December 2019, the causative agent of the Covid-19 pandemic (SARS-CoV-2), emerged in Wuhan, Hubei, China (Huang et al. 2020). As of 15 January, 2021, there are ∼93 million confirmed cases and ∼2 million deaths in 191 countries (Johns Hopkins Coronavirus Resource Center 2020). Unprecedented international efforts of viral sequencing have allowed the submission of ∼375,000 genomes in the Global Initiative on Sharing All Influenza Data (GISAID) up to date (Shu and McCauley 2017), which are now available for studies of genomic epidemiology to follow the evolutionary history and dynamics of SARS-CoV-2 through space and time. In this sense, some important studies were already conducted in highly-affected countries, including China (Lu et al. 2020), USA (Deng et al. 2020; Fauver et al. 2020; Ladner et al. 2020; Maurano et al. 2020), Italy (Bartolini et al. 2020), Netherlands (Oude Munnink et al. 2020), Australia (Rockett et al. 2020; Seemann et al. 2020), and Brazil (Candido et al. 2020b; Paiva et al. 2020; Xavier et al. 2020).

By using a nomenclature developed to capture local and global patterns of genetic diversity of the virus, two main lineages (A and B) were identified, both originated in Wuhan and with simultaneous spreading around the world (Rambaut et al. 2020). The dynamics of viral transmission in the Brazilian territory was investigated through the sequencing of ∼500 genomes until the end of April, 2020. It was determined that: (i) B.1 and derived lineages were predominant at the beginning of the pandemic; (ii) >100 independent international introductions occurred in the country; (iii) a significant movement of the virus among the Brazilian regions was observed after international travel restrictions; and (iv) non-pharmacological measures were able to reduce the reproduction number (R) from >4 to <1 (Candido et al. 2020b).

The genetic diversity of SARS-CoV-2 has been extensively studied, evidencing the presence of recurrent mutations across the world (Takahiko Koyama et al.; Laamarti et al. 2020; van Dorp et al. 2020), their impact on pathogenicity and transmissibility of the virus (Korber et al. 2020; Li et al. 2020; Toyoshima et al. 2020; Volz et al. 2020), and on the role of mutations in the development of new vaccines (Dearlove et al. 2020). In addition to SARS-CoV-2 mutations, co-infection with other pathogens may be associated with poor clinical outcomes, although these rates appear to be limited (Calcagno et al. 2020; Kim et al. 2020; Nowak et al. 2020; Peddu et al. 2020).

As of March 10, 2020, a 60-year-old man who had been in Italy, became the first confirmed case in the southernmost state of Brazil (Rio Grande do Sul − RS) (Rio Grande do Sul Department of Health, 2020), which is the most populous state in the South Region of Brazil and the fifth in the whole country (∼11.5 million inhabitants) (Brazilian Institute of Geography and Statistics - IBGE). Currently, Brazil has ∼8.9% of worldwide cases (∼8.3 million) and is the third worst-hit country (Johns Hopkins Coronavirus Resource Center, 2020). The RS State reported ∼500,792 cases and 9,838 deaths, with ∼7% of cases requiring hospitalization (Rio Grande do Sul Department of Health, 2020). The municipality of Esteio, located in the metropolitan region of RS capital, reported 5,347 cases and 142 deaths (Esteio Department of Health, 2020). As Esteio is a commuter town, many workers move to and return from the state capital every day.

Thus, we aimed to characterize the main circulating lineages in Esteio (RS, Brazil) and their relationship with global, national and regional lineages using phylogenetics and phylodynamics inference from 21 SARS-CoV-2 genome sequences, including the investigation of putative viral mutations related to poor outcomes. Additionally, due to our typical subtropical climate and therefore high occurrence of respiratory infections, we investigated the occurrence of co-infections with other viral pathogens in these samples. The choice of a small municipality as the target of this study was important since we could more easily and precisely follow infected individuals, allowing a more detailed surveillance on the spread of the virus and detection of variability.

## Results

SARS-CoV-2 genomes were obtained with an average coverage depth of 1,380.51× (range: 5.92× − 6,224.73×, median: 213.28×, standard deviation: 2,296.16×) (Supplemental File 1). All consensus genomes passed the quality control steps. Considering the 21 sequences (Table 1), 52.4% of the patients were female and the mean age was 41.3 years (range: 19-72 years). The mean Cycle threshold (Ct) values was 16.12 (range: 12.53-19.94). None of the patients reported interstate or international travels. Regarding clinical status, 90.5% of patients were classified as mild infection and 9.5% as moderate.

**Table 1.** Epidemiological data of the 21 sequenced samples from Esteio, RS, Brazil.

| GISAID Accession | Ct value | Collection month | Age range | Sex | Clinical status | Pangolin Lineage | Nextstrain Clade |
| --- | --- | --- | --- | --- | --- | --- | --- |
| EPI_ISL_831678 | 16.15 | May 2020 | 20-30 | M | Mild | B.1.1.33 | 20B |
| EPI_ISL_831474 | 16.48 | June 2020 | 20-30 | M | Mild | B.1.1.33 | 20B |
| EPI_ISL_831645 | 16.72 | June 2020 | 60+ | M | Moderate | B.1.1.248 | 20B |
| EPI_ISL_831646 | 16.45 | June 2020 | 50-60 | F | Mild | B.1.1.33 | 20B |
| EPI_ISL_831660 | 15.53 | June 2020 | 20-30 | F | Mild | B.1.1.248 | 20B |
| EPI_ISL_831681 | 16.72 | July 2020 | 40-50 | F | Mild | B.1.1 | 19A |
| EPI_ISL_831683 | 15.50 | July 2020 | 30-40 | F | Moderate | B.1.1.33 | 20B |
| EPI_ISL_831685 | 16.65 | July 2020 | 10-20 | F | Mild | B.1.1.33 | 20B |
| EPI_ISL_831688 | 15.52 | July 2020 | 40-50 | M | Mild | B.1.1.248 | 20B |
| EPI_ISL_831689 | 14.37 | August 2020 | 30-40 | M | Mild | B.1.1.248 | 20B |
| EPI_ISL_831892 | 15.12 | August 2020 | 60+ | M | Mild | B.1.1.33 | 20B |
| EPI_ISL_831898 | 14.14 | August 2020 | 40-50 | M | Mild | B.1.1.33 | 20B |
| EPI_ISL_831913 | 12.53 | August 2020 | 30-40 | F | Mild | B.1.1.49 | 20B |
| EPI_ISL_831938 | 15.32 | August 2020 | 50-60 | M | Mild | B.1.1.248 | 20B |
| EPI_ISL_831939 | 15.58 | September 2020 | 40-50 | M | Mild | B.1.1 | 20B |
| EPI_ISL_831940 | 17.10 | September 2020 | 20-30 | F | Mild | B.1.1.33 | 20B |
| EPI_ISL_832009 | 14.99 | September 2020 | 30-40 | F | Mild | B.1.1.248 | 20B |
| EPI_ISL_832010 | 18.31 | October 2020 | 20-30 | F | Mild | B.1.1.248 | 20B |
| EPI_ISL_832011 | 17.33 | October 2020 | 40 | M | Mild | B.1.1.248 | 20B |
| EPI_ISL_832012 | 19.94 | October 2020 | 60+ | F | Mild | B.1.1.33 | 20B |
| EPI_ISL_832013 | 18.15 | October 2020 | 40-50 | F | Mild | B.1.1 | 20B |

All samples were nasopharyngeal swabs collected from patients of the municipality of Esteio. Sample ID=Sample identifier; M=Male; F=Female.

### Virome analysis

To investigate whether the severity of the infection presented by the patients could be linked to co-infection with another respiratory viral pathogen, we analyzed the viral composition of these samples. We found through taxonomic classification at the level of nucleotides and amino acids that none of the investigated patients had a viral infection other than Covid-19. All samples had high assignment (>99%) to the *Betacoronavirus* genus.

### SARS-CoV-2 mutations found in the patient samples

The number of Single Nucleotide Polymorphisms (SNPs) per genome ranged from 1 to 19 (mean: 12.8, median: 14.0) (Figure 1A). All genomes were different from each other. We identified 80 different SNPs in the 21 genomes analyzed. Thirty two (40.0%) of them were observed in more than one sample (Supplemental Table 1). Of these, 18 (56.2%) were missense (non-synonymous). High frequency (>5 genomes) missense mutations were observed in the following positions (absolute nucleotide position: amino acid inside the gene): ORF1ab (C12053T: L3930F), Surface (S) glycoprotein (A23403G: D614G; G25088T: V1176F), ORF6 (T27299C: I33T), and Nucleocapsid (N) protein (GGG28881-28883ACC: RG203-204KR; T29148C: I292T) (Figure 1B). A mutation in the Receptor Binding Domain (RBD) of the Spike protein (G23012A: E484) was found in two genomes (9.5%).

**Figure 1.**
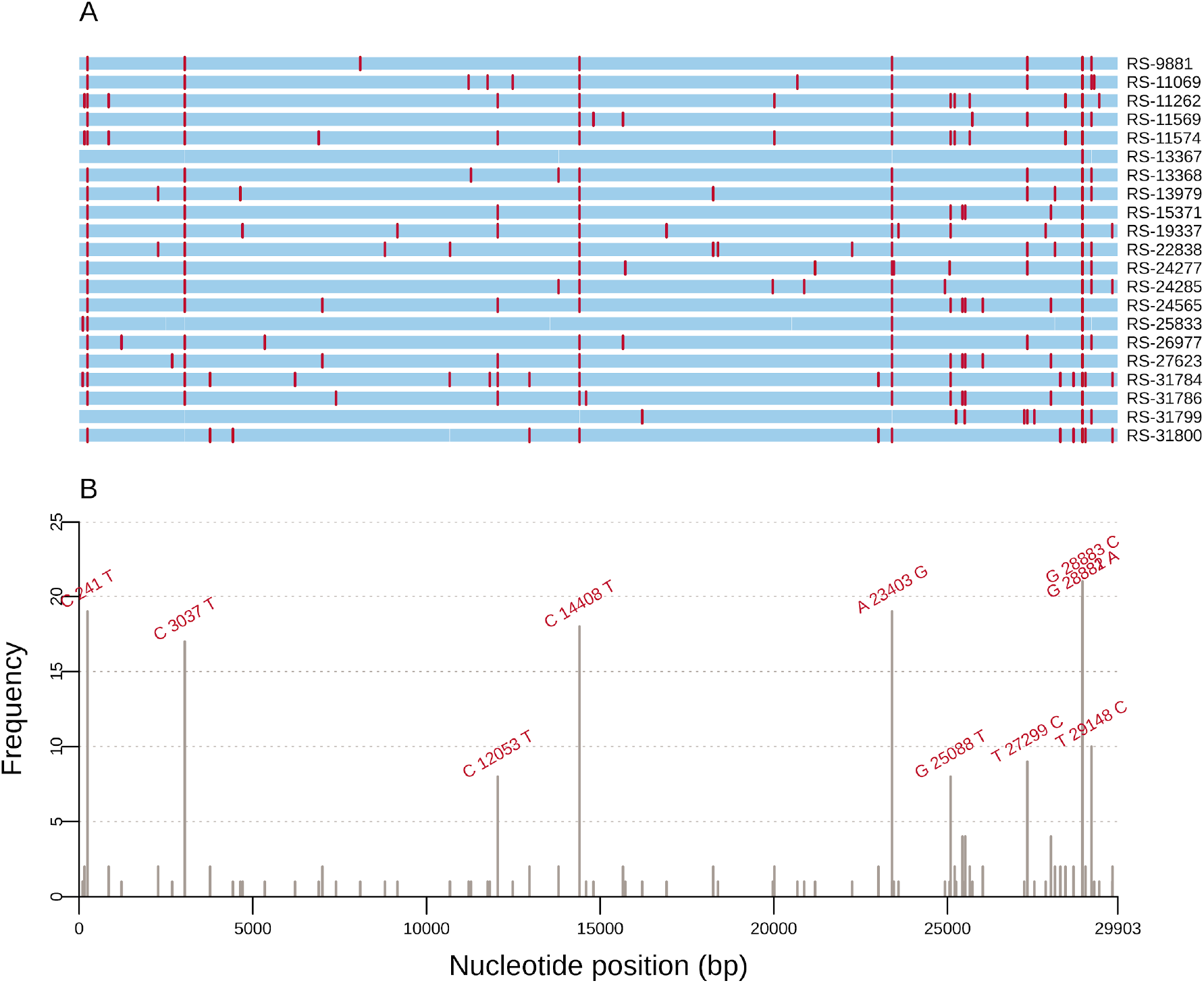
Mutations of SARS-CoV-2 genomes from patients of Esteio, RS, Brazil. (A)Genome map for the 21 genomes sequenced (indicated in the right). SNPs are colored in red. (B) Frequency of SNPs per SARS-CoV-2 genome position among the 21 genome sequences. These mutations correspond to the red lines in (A) and only those represented by >5 genomes are indicated above the bars, regardless of being synonymous or missense.

We were able to identify four different viral lineages, all descendants of lineage B (Table 1). Two lineages probably associated with community-transmission in Brazil, B.1.1.33 (n=9; 42.9%) and B.1.1.248 (n=8; 38.1%) were the most prevalent. All B.1.1.33 sequences shared T27299C (ORF6:I33T), GGG28881-28883AAC (N:RG203-204KR), and T29148C (N:I292T) mutations. All B.1.1.248 sequences shared C241T (5’ UTR), C3037T (ORF1ab nsp3:F924), C12053T (ORF1ab nsp7:L3930F), C14408T (ORF1ab RdRp:L4715), A23403G (S:D614G), G25088T (S:V1176F), and GGG28881-28883AAC (N:RG203-204KR) replacements.

Both lineages are represented by >80% of Brazilian sequences in global context. They are the most representative lineages in the South Region of Brazil and in the whole country (Figure 2, Supplemental Figures 1, 2 and 3). Three genomes were classified as B.1.1 lineage, which are the most globally widespread lineages characterized by RG203-204KR mutations in the nucleocapsid phosphoprotein (https://cov-lineages.org; Rambaut et al. 2020).

**Figure 2.**
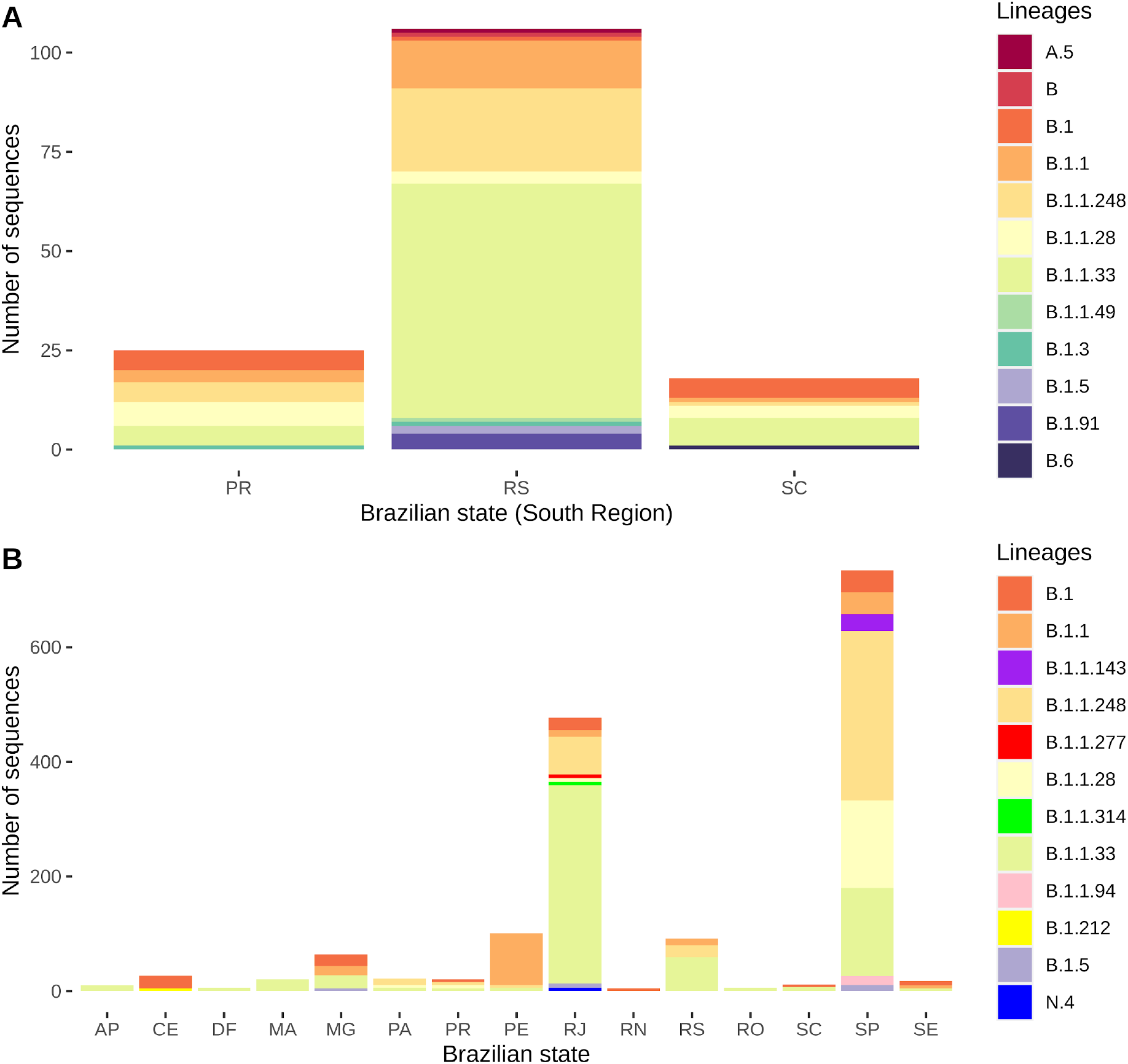
Most prevalent lineages in Brazil considering all 1778 Brazilian sequences available in the GISAID database. (A) All sampled viral lineages from Southern Brazil, including Paraná (PR), Rio Grande do Sul (RS) and Santa Catarina (SC) states. The RS sequences include the 21 viral genomes from this work and 85 from GISAID. (B) Most prevalent lineages from Brazilian states. Only states that have more ≥ 5 sequences labelled as the lineage under consideration were included. AP=Amapá; CE=Ceará; DF=Distrito Federal; MA=Maranhão; MG=Minas Gerais; PA=Pará; PE=Pernambuco; RJ=Rio de Janeiro; RN=Rio Grande do Norte; RO=Rondônia; SP=São Paulo; SE=Sergipe.

Finally, a relatively rare lineage (B.1.1.49) mostly found in Wales and Denmark was also assigned. After inspecting these sequences assigned to global lineages (B.1.1 and B.1.1.49), we verified that in all cases there were characteristic mutations of B.1.1.248 and B.1.1.33 lineages flagged as undetermined bases (N character; depth of coverage (DP) < 10) in the consensus genome. After reclassifying these sequences using low coverage variants, two were attributed to B.1.1 lineage, one to B.1.1.248 and one to B.1.1.33 (Supplemental File 2). Therefore, due to the absence of confirmed relationships with other countries and the presence of other defining-lineage variants with low coverage, it seems possible that these sequences are also the result of community transmission in Brazil and were not introduced independently in the municipality of Esteio from other countries.

We also detected two novel epidemiologically-related clusters until then unknown. Two patients had three unique mutations in genome positions 25207 (Spike), 25642 (ORF3a), and 28393 (Nucleocapsid), all synonymous substitutions. The patients both live in the same neighborhood, about 100 meters from each other and did the test in a 2-day interval. We also identified another cluster of four patients characterized by three unique mutations: 25429 (ORF3a: V13L), 25509 (ORF3a), and 27976 (ORF8: H28R). The tests of these patients were performed in a 3-month interval (July 15 - October 13). The two clusters are linked to viruses belonging to B.1.1.248 lineage, suggesting the existence of specific mutation signatures even within lineages.

### Phylogenetics analysis

After running the Nextstrain pipeline for quality control and subsampling, we obtained 3,758 time-, geographical- and genetic representative genomes to proceed phylogenetic inferences. Of these, 393 were from Africa, 800 were from Asia, 1,203 from Europe, 235 from North America, 127 from Oceania, and 1,000 from South America. Considering the latter, 609 were from Brazil and 98 from Rio Grande do Sul (21 from this study plus 77 from GISAID that passed the quality control criteria) (Supplemental File 3).

The time-resolved phylogeny confirmed that the majority of the time-representative sequences from Esteio are the result of community transmission within Brazil. The sequences grouped mostly in two clades (Figure 3A, Supplemental Figure 4) corresponding to lineages B.1.1.248 and B.1.1.33. Clade 1 comprised 147 sequences: 20 from RS, 58 from SP, 46 from RJ, 2 from PR, and 2 from SC, therefore mostly widespread in Brazilian states (Figure 3B). Clade 2 included 277 genomes: 50 from RS, 72 from RJ, 20 from SP, 5 from SC, 30 from Chile, 10 from Argentina, and 6 from Uruguay (Figure 3C). These results suggest a clade distributed through South American countries. Esteio sequences are relatively evenly distributed through these clades mostly represented by Brazilian and RS genomes. Exceptions to this observation were the two previously described epidemiologically-linked clusters, whose sequences grouped together in the B.1.1.248 lineage as expected (Figure 3C). Given the low mutation rate of SARS-CoV-2 (6.59×10^−4^, ∼19 mutations per year) (Supplemental Figure 5), we believe that this would indicate at least four introductions of lineage B.1.1.248 and seven introductions of lineage B.1.1.33 in the municipality of Esteio, probably from other locations in Brazil, and a national movement of the virus even to more distant places like the southernmost state of Brazil. Likewise, despite the large representativeness of Brazilian samples within these two major clades, we also found other sequences from Asia, Europe, Oceania, and South America. Therefore, sequences from these clades seem to have been directly transmitted from Brazil to other countries.

**Figure 3.**
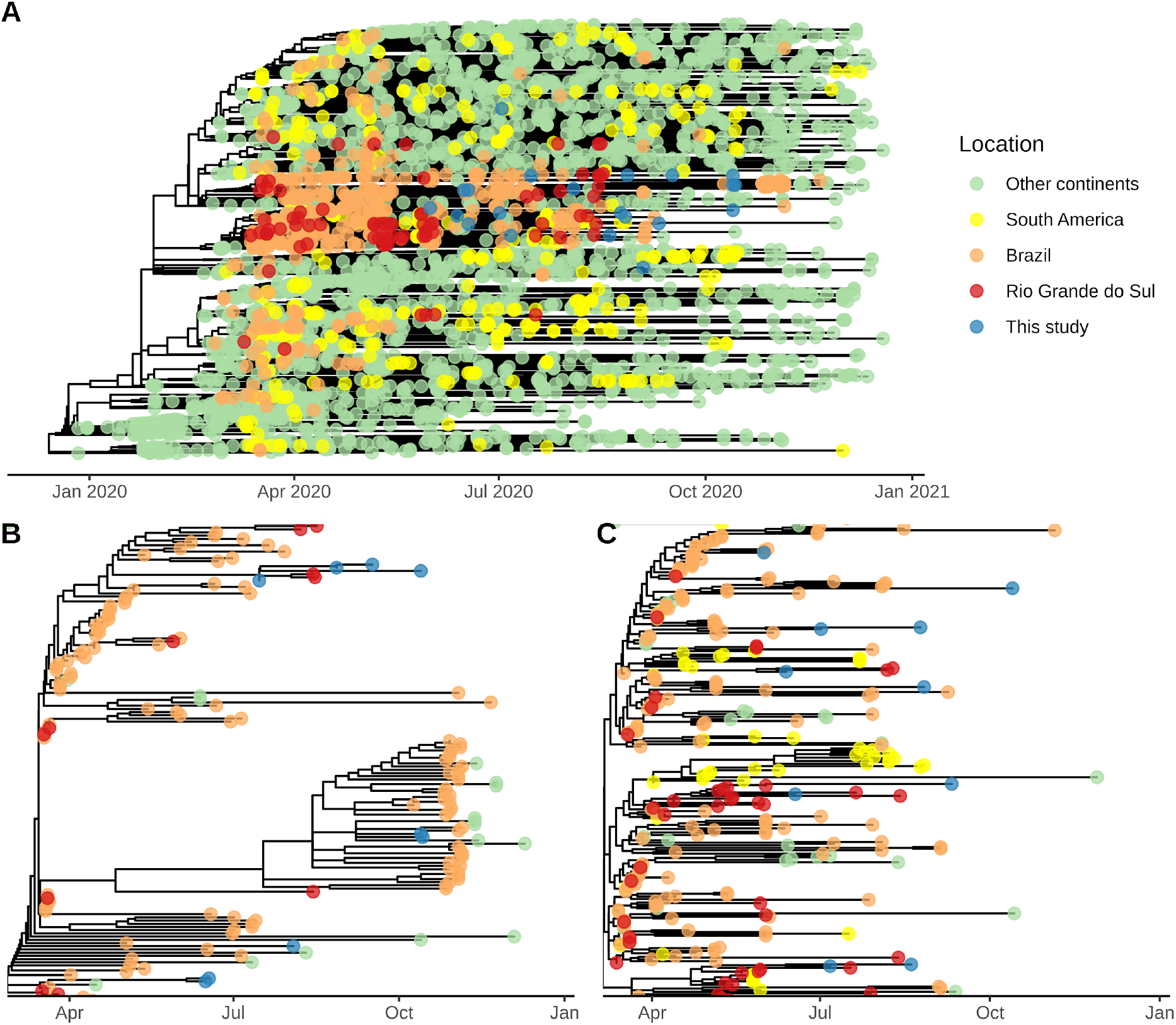
Time-resolved phylogeny inferred from our 21 sequences combined with other 3,758 worldwide genomes sampled between December 2019 and December 2020. (A) Worldwide distribution of sequences in the subsampled tree, highlighting genomes from this study. (B)Zoom-in on sequences from this study belonging to Clade 1 / lineage B.1.1.248 (upper clade in A). (C) Zoom-in on sequences from this study belonging to Clade 2 / lineage B.1.1.33 (bottom clade in A). In (B) and (C), the clade is initiated from the T_MRCA_ of the sequences presented in this study and it does not refer to all the sequences assigned to specific lineages.

**Figure 4.**
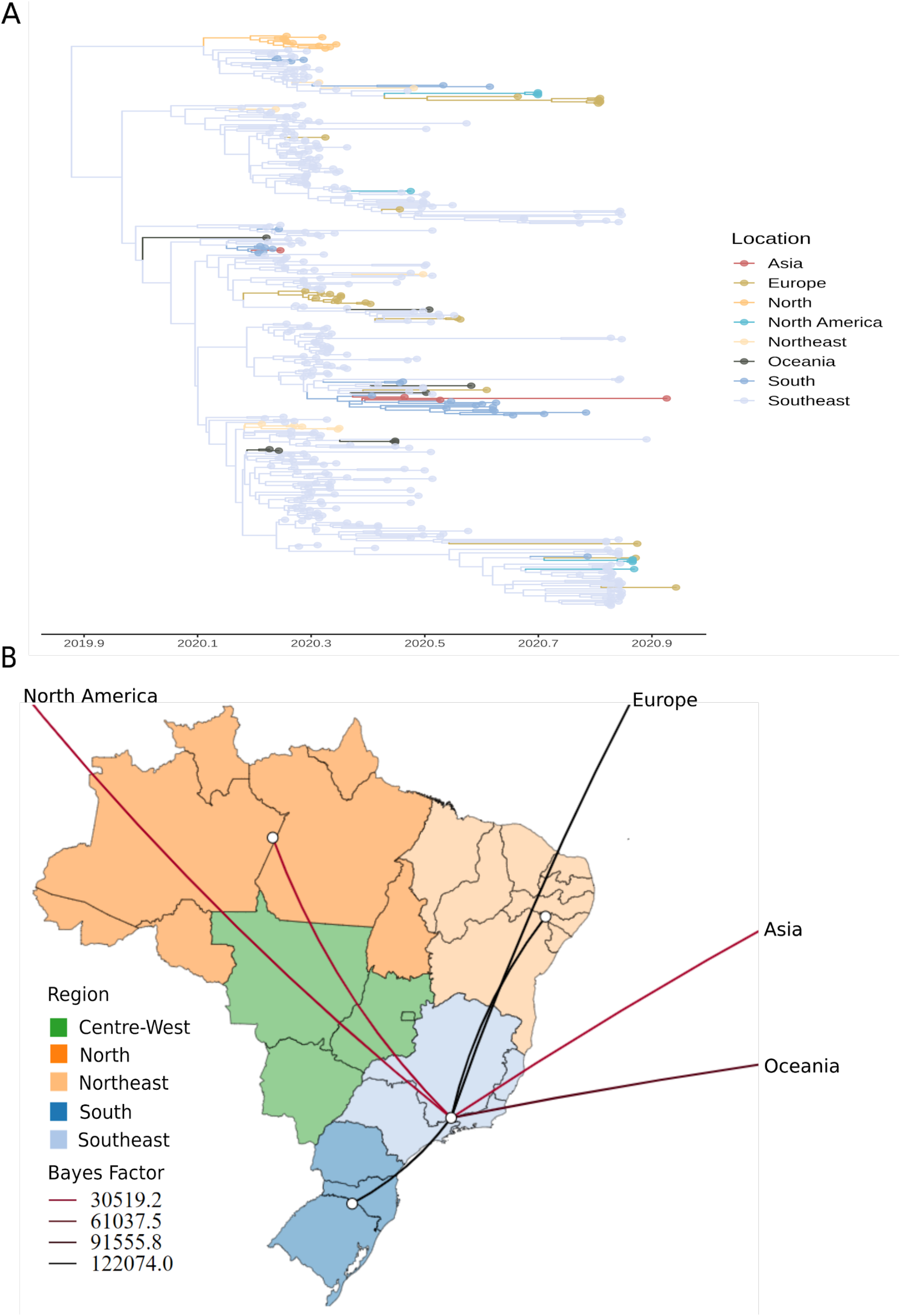
Spatiotemporal diffusion of B.1.1.248 lineage across Brazilian regions and other continents. (A) Time-resolved MCC tree of 405 genomes belonging to B.1.1.248 lineage. (B) Well-supported transition rates between Brazilian regions and other continents in discrete phylogeographic reconstructions using BSSVS procedure and Bayes Factor tests. Transition rates with posterior probabilities < 0.5 were cut-off.

**Figure 5.**
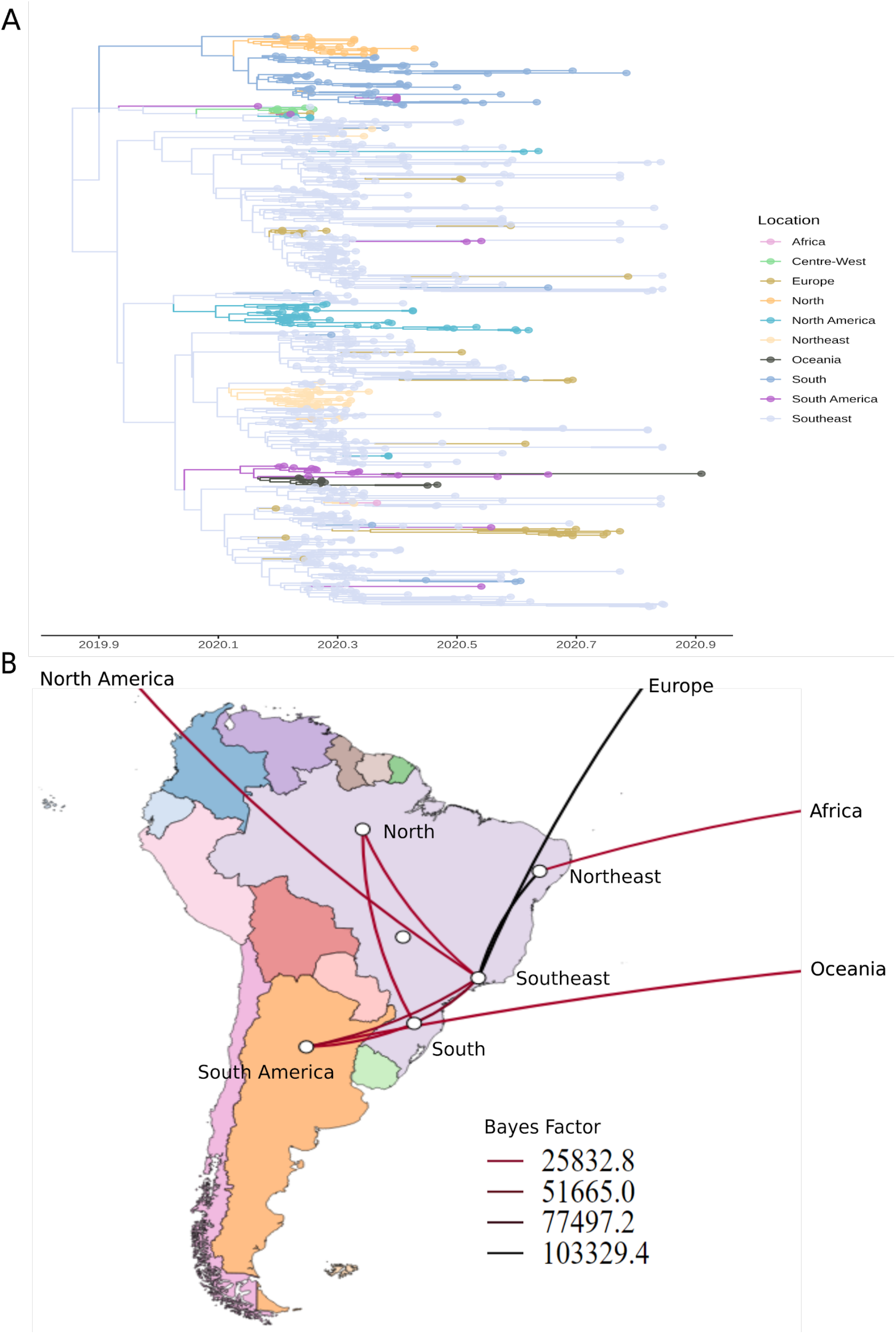
Spatiotemporal diffusion of B.1.1.33 lineage across Brazilian regions, South America and other continents. (A) Time-resolved MCC tree of 725 genomes belonging to B.1.1.33 lineage. (B) Well-supported transition rates between Brazilian regions and other continents in discrete phylogeographic reconstructions using BSSVS procedure and Bayes Factor tests. Transition rates with posterior probabilities < 0.5 were cut-off.

### Phylodynamics and phylogeographic analysis

Time-measured phylogeographic analysis highlighted the major contribution of Southeast in Brazilian and worldwide diffusion of both lineages (Figures 4 and 5). B.1.1.248 lineage was probably introduced from Southeast to Northern, Northeast, and Southern Brazil, as well as Asia, Europe, North America and Oceania (Bayes Factor (BF) > 30; Posterior Probability (PP) > 0.8) (Figure 4B). The four major clades from Southern Brazil in the B.1.1.248 Maximum-Clade Credibility (MCC) tree were probably introduced from Southeast with T_MRCA_ ranging from 26 February, 2020 (95% Highest Posterior Density (HPD): 25 February - 11 April) to 17 April (95% HPD: 17 April - 9 May) (Figure 4A).

B.1.1.33 lineage was probably exported from Southeast to Northern, Northeast, and Southern Brazil, as well as Europe, North and South America (BF > 30; PP > 0.8) (Figure 5B). Well-supported rates were also identified from Northeast to Africa (BF = 31.10; PP = 0.79) and from South America to Oceania (BF = 56.99; PP = 0.87). Viruses belonging to this lineage appear to have a major contribution in Southern Brazil epidemics, since its sequences formed a monophyletic clade with >50 sequences in the B.1.1.33 MCC tree (Figure 5A), whose T_MRCA_ was dated from January 29, 2020 (95% HPD: January 23 - March 02). Furthermore, this analysis confirmed that Southern Brazil is the most probable source of importations of available B.1.1.33 sequences to South-American countries (BF = 1467.97; PP = 0.99) and Northern Brazil (BF = 9.45; PP = 0.53) (Figure 5B).

## Discussion

In the present work, we accessed SARS-CoV-2 mutations, circulating lineages and phylogenetic patterns of SARS-CoV-2 from a time- and age-representative set of patients admitted in a municipal healthcare system from the Southern region of Brazil, the third country most affected by the Covid-19 pandemic. The SARS-CoV-2 spike (S) glycoprotein mediates the interaction with the Angiotensin-converting enzyme 2 (ACE2) receptor in the host cells and it is the primary target of neutralizing antibodies (Walls et al. 2020). A mutation in S (D614G) was recently associated with higher viral loads (Korber et al. 2020), increased replication on human lung epithelial cells (Plante et al. 2020), and younger age of patients (Volz et al. 2020), but not with the disease severity (Korber et al. 2020; Volz et al. 2020). This mutation is associated with an abolition in the hydrogen bond between the aspartate originally located at 614 position and a threonine residue in the 859 neighbouring promoter of the spike trimer, thus increasing the probability of RBD being found in the open state. This promotes the binding with the ACE2 receptor leading to enhanced infectivity (Plante et al. 2020). Importantly, this replacement was detected in 90.5% of our samples, compatible with the dominance of this variant currently in global context (221,700 sequences [Updated 14 December, 2020]; Singer et al. 2020). Also concerning the S protein, we identified the V1176F variant in 38.1% of our samples and in all of them there was a co-occurrence with D614G. Importantly, 349/535 (65.2%) of the sequences already isolated in the world including this replacement were from Brazil (Singer et al. 2020), indicating that this mutation may become fixed.

Another replacement in the RDB of S protein (E484K) was also assigned. Currently, 157 genomes have this mutation globally, 114 (72.6%) isolated from South Africa, where a new lineage (501Y.V2) characterized by three RBD mutations (K417N, E484K and N501Y) recently emerged (Tegally et al. 2020). Recent evidence showed that E484K replacement enables viral escape from neutralizing monoclonal antibodies or polyclonal sera (Baum et al. 2020; Greaney et al. 2020; Weisblum et al. 2020) facilitating reinfection by emerging lineages harboring this mutation as reported in Brazil (Nonaka et al. 2021). Importantly, Brazil/RS-31784 and Brazil/RS-31800 sequences had four of five shared mutations with a new lineage recently reported in the Rio de Janeiro state (synonymous C28253T (F120F; ORF8), missense G28628T (A119S) and G28975T (M234I) in N protein and C29754U (3’ UTR)), in addition to E484K RBD replacement (Voloch et al. 2020). They were sampled in late-October, as the first cases of the RJ lineage (Supplemental File 2), while our samples were collected in mid-October. Phylodynamics inferences pointed that this lineage emerged in early July, approximately four months before the detection of its first genomes (Voloch et al. 2020). Moreover, its identification in a small municipality from the RS state (located 1.5 thousand kilometers from RJ) demonstrates that it emerged months later and may be already widely distributed in the Brazilian territory, but went unnoticed so far by the lack of genomic surveillance in Brazil.

The substitution of a negatively charged amino acid (Glutamic Acid) for the positively charged lysine has a profound impact upon a highly flexible loop at the RBD. More specifically, it creates a strong ion interaction between lysine and amino acid 75 of hACE-2 (the main SARS-CoV-2 receptor). This link is not present in the wild type E484. Shifting key elements of RBD responsible for interactions may have a major impact in the risk of immune evasion. Also E484K could be related to enhanced infectivity, which may be associated with the rapid dissemination of these escape mutants (Nelson et al. 2021). The publication of a recent reinfection case of SARS-CoV-2 harboring E484K and the presence of this mutation in Covid-19 patients during the current second wave in Northern Brazil are highly suggestive that this mutation is critical for viral evolution and thus must be investigated thoroughly (Nonaka et al. 2021).

Additionally, a tri-nucleotide mutation in the Nucleocapsid gene (RG203KR) was also observed in high frequency in this study (∼100%), which is present in about 1/3 of the genomes available in GISAID (Singer et al. 2020). The missense mutation in ORF1ab (L3930F) was reported in other 429 sequences (125 from Brazil) and also in two sequences of lineage B.1.1.248 from the Philippines (Velasco et al. 2020). Interestingly, the I33F and I292T mutations that were found in ORF6 and N, respectively, have been considered dominant mutations in the SARS-CoV-2 sequences from Brazil (Franco-Muñoz et al. 2020; Singh et al. 2020). The co-occurrence of these mutations represents the signature of Clade 2, which were one of the three most prevalent Brazilian viral lineage groups in the beginning of the pandemic, highly widespread in 16 states from Brazil (Candido et al. 2020b). It is important to emphasize that functional studies are necessary to characterize the effect of each viral mutation on transmissibility and pathogenicity. It is expected that most mutations will not have a great impact on viral evolution and its relationship with the human host. However, some of them may increase the viral fitness or represent some viral advantage in the pathogen-host interaction and, consequently, may become fixed in the population.

Compared to three other studies conducted in Brazil (Candido et al. 2020b), in the states of Minas Gerais (Xavier et al. 2020) and Pernambuco (Paiva et al. 2020) in the early phase of pandemic — which showed the introduction of viral lineages from other continents (mainly Europe) by international returning travelers —, this study suggests a minor role of international lineages in the ongoing viral transmission in Esteio. We speculate a trend towards the perpetuation and diversification of the lineages found in this study (B.1.1.248 and B.1.1.33) inside Brazil, unless new importations massively occur and these new imported lineages begin to circulate. The dissemination of these lineages were also reported in the Uruguayan-Brazilian border, driving viral introductions mainly from Southeast and Southern Brazil (especially RS state) to Uruguay (Mir et al. 2021). In this study, we found consistent results, mainly regarding B.1.1.33 diffusion from Southern and Southeast to South-American countries (*e. g*. Argentina, Chile and Uruguay). These lineages have already formed new sublineages (https://cov-lineages.org/lineages.html). B.1.1.248 has evolved in N.1 (USA lineage), N.2 (French), N.3 (Argentinian), and N.4 (Chilean). Furthermore, B.1.1.248 has evolved in P.1 (Manaus lineage associated with a constellation of Spike mutations like B.1.1.7 and B.1.351 (Faria et al. 2021)) and P.2 (RJ lineage found in this study).

We built a time-resolved phylogeny prioritizing sequences that are genetically and spatially closer, but maintaining a global representativity of viral spread. This allowed us to confidently identify that our sequences fell into two main clades, with a broad presence of Brazilian and local sequences. We also inferred the spatiotemporal diffusion of these main lineages in regional and global context, finding the key role of Southeast in disseminating these lineages across Brazilian states and other continentes. We also found a broader clade represented by Southern Brazil sequences and its important contribution in disseminating B.1.1.33 to South-American countries. Moreover, we found four broader clades in the B.1.1.248 MCC phylogeny, suggesting multiple introduction events from Southeast followed by community transmission. It is also important to highlight that a large proportion of the samples available for Brazil come from the Southeast, which is expected to impact conclusions about the real epidemiological processes and to overestimate Southeast contribution in diffusion to other regions of Brazil. However, a study from the beginning of pandemic (February-March 2020) estimated that the main destinations of the international passengers arriving to Brazil were São Paulo (46.1%), Rio de Janeiro (21%) and Belo Horizonte (4.1%), three capitals from the Southeast and therefore routes for Covid-19 importation (Candido et al. 2020a). Moreover, during mid-February and mid-March, SARS-CoV-2 spread mostly locally and within-state borders. In contrast, during mid-March and mid-April there was an ignition of the epidemic from the Southeast region to other states (Candido et al. 2020b), which is consistent with our findings.

Some limitations should be considered. Firstly, it was not possible to analyze a larger sample size. Moreover, the low quantity of sequences from the RS state to contextualize our sequences limited the inference of events of introduction and movement of the virus with municipal and state resolution. Still in this respect, we have observed a dramatic drop in the sequencing efforts from Brazil after April (Candido et al. 2020b), which made it difficult to measure the main circulating lineages in the country during our investigation period (May-October, 2020) and may introduce confounding factors.

Nevertheless, our results provide a comprehensive view of viral mutations from a time- and age-representative sample from May to October 2020, highlighting two frequent mutations in Spike glycoprotein (D614G and V1176F), an emergent mutation in Spike RBD (E484K) characteristic of the South African lineage B.1.351, and the adjacent replacement of 2 amino acids in Nucleocapsid phosphoprotein (R203K and G204R). A significant viral diversity was evidenced by the absence of identical isolates in our samples. Furthermore, we identified patterns of SARS-CoV-2 viral diversity inside Southern Brazil, demonstrating the major role of community transmission in viral spreading and the establishment of Brazilian lineages. This fact was demonstrated by the dominance of lineages B.1.1.248 and B.1.1.33, widely distributed throughout the Brazilian states and with very low occurrence in other countries. In particular, to our best knowledge, we describe the earliest E484K harboring SARS-CoV-2 in Brazil. Since this mutation has been associated with loss of neutralizing activity from convalescent plasma (immune evasion) and enhanced interaction with hACE-2, lineages containing this substitution must be the subject of intense surveillance. Interestingly, Rio Grande do Sul state has lagged behind northern Brazil in COVID-19 cases. However, introduction of E484K mutants shows temporal correlation with later increases in new cases in our state. Unfortunately, the lack of consistent genomic data excludes us the capability of exploring further the connection between emergent mutations and the SARS CoV 2 spreading patterns. Our data show the value of correlating clinical, epidemiological and genomic information for the understanding of viral evolution and its spatial distribution over time. This is of paramount importance to better inform policy making strategies to fight Covid-19. More specifically, it is critical that immune strategies such as convalescent plasma and vaccines be tested against these new variants. Attempts to demonstrate activity against S mutans should be a priority effort for all vaccine and monoclonal antibody makers. Second generation immune therapies might have to be directed at more conservative neutralizing binding sites (such as S2 fusion domain) or elicit strong cellular response in order to keep on long term protection.

## Methods

### Ethics statement

Ethical approval was obtained from the Brazilian’s National Ethics Committee (Comissão Nacional de Ética em Pesquisa — CONEP) under process number 30934020.5.0000.0008. All samples were anonymized before received by the study investigators.

### Sample collection and clinical testing

Nasopharyngeal samples were obtained from patients of the Hospital São Camilo, Secretaria Municipal de Esteio and Vigilancia em Saude from Esteio, Rio Grande do Sul (RS), Brazil. Nasopharyngeal swabs were collected and placed in Viral Transport Medium (VTM, Copan Universal Transport Medium). Samples were transported to the Molecular Microbiology Laboratory from Feevale University and tested on the same day for SARS-CoV-2 by reverse-transcriptase quantitative polymerase chain reaction (RTq-PCR). Remnant samples were stored at −80°C. SARS-CoV-2 diagnosis was performed using the Charité RT-qPCR assays. The RT-PCR assay used WHO-recommended primers and probes targeting the E and RdRp genes (Corman et al. 2020).

We selected 21 samples with RT-qPCR positive results, collected from May 31 to October 13, 2020 from patients residing in the municipality of Esteio, RS, Brazil. We included patients who presented symptoms such as fever, cough, sore throat, dyspnea, anosmia, fatigue, diarrhea and/or vomiting (mild and moderate clinical status). Additionally, samples were selected based on cycle threshold (Ct) values ≤ 20. Electronic medical records were reviewed to compile epidemiological metadata (*e. g*., date of collection, sex, age, symptoms, exposure history, and clinical status).

### RNA extraction, library preparation and sequencing

We submitted the RT-qPCR positive for SARS-CoV-2 swabs to genomic RNA extraction. This process was performed in the automated nucleic acid purification system KingFisher™ Duo Prime Purification System (ThermoFisher Scientific, Waltham, USA) along with the MagMax™ CORE Nucleic Acid Purification Kit (ThermoFisher Scientific, Waltham, USA).

The extracted and purified genomic RNA was transcribed to cDNA using Maxima H Minus Double-Stranded cDNA Synthesis Kit, catalog number K2561 (ThermoFisher Scientific, Waltham, USA) following the manufacturer’s instructions. Library preparation was achieved using Nextera™ Flex for Enrichment with RNA Probes (Illumina, San Diego, USA). Briefly, we performed tagmentation - the process that fragments and tags the DNA with adapter sequences - in a pre-programmed thermocycler incubation temperature, until holding at 10 °C. This step uses the Enrichment Bead-Linked Transposomes (Enrichment BLT, eBLT) to tagment DNA followed by post tagmentation clean up. The PCR procedure adds pre-paired 10 base pair adapters and sequences required for sequencing cluster generation. The viral cDNA was used as input for multiple overlapping PCR reactions that spanned the viral genome using PCR Master Mix (Enhanced PCR Mix reagent and nuclease-free water). The amplified tagmented DNA was cleaned with AMPure XP magnetic beads (Beckman Coulter Inc., Indianapolis, USA) to remove shorter DNA fragments and other impurities. We then quantified the cleaned libraries employing Qubit dsDNA BR Assay Kit (Thermo Fisher Scientific, Waltham, USA).

Sequencing was performed on an Illumina Miseq® (Illumina, San Diego, USA) using Reagent Kit v3 with 150 cycles in a paired-end run, following the manufacturer’s instructions. All experiments were performed in a biosafety level 2 laboratory.

### Consensus calling

Reference mapping and consensus calling was performed using an in-house developed pipeline managed with Snakemake (Köster and Rahmann 2012). Briefly, quality control was performed FastQC v0.11.9 and low-quality reads and adapters were removed using Trimmomatic v0.39 (Bolger et al. 2014). PCR duplicates were discarded using Picard MarkDuplicates v2.23.8 (https://broadinstitute.github.io/picard/) with `REMOVE_DUPLICATES=true`. Reads were mapped to the reference SARS-CoV-2 genome (GenBank accession number NC_045512.2) using Burrows–Wheeler Aligner (BWA-MEM) v0.7.17 (Li and Durbin 2009) and unmapped reads were discarded. Consensus sequences were generated using bcftools mpileup combined with bcftools consensus v1.9 (Li 2011). Positions covered by fewer than 10 reads (DP<10) were considered a gap in coverage and converted to Ns. Coverage values for each genome were calculated using bedtools v2.26.0 (Quinlan 2014) and plotted using the karyoploteR v1.12.4 package (Gel and Serra 2017). Finally, we assessed genome consensus sequence quality using Nextclade v0.8.1 (https://clades.nextstrain.org/) and CoV-GLUE (http://cov-glue.cvr.gla.ac.uk/; Singer et al. 2020).

### Virome analysis

As the respiratory panel kit used allows the detection of ∼40 respiratory viral pathogens, the viral composition of each sample was verified using Kaiju v1.7.3 (Menzel et al. 2016) and Kraken v2.0.7-beta (Wood et al. 2019) against a reference database of viral sequences. The viral database for each tool was built with the following commands, respectively: `kaiju-makedb -s visuses`. and `kraken2-build --download-library viral`. Taxonomic classification interactive charts were visualized using Krona (Ondov et al. 2011).

### Mutation analysis

Sequence positions in this work refer to GenBank RefSeq sequence NC_045512.2, a genome isolated and sequenced from Wuhan (China), early in the pandemic. SNPs and insertions/deletions (INDELs) were assessed in each sample by using snippy variant calling and core genome alignment pipeline v4.6.0 (https://github.com/tseemann/snippy), which uses FreeBayes v1.3.2 (Garrison and Marth 2012) variant caller and snpEff v5.0 (Cingolani et al. 2012) to annotate and predict the effects of variants on genes and proteins. Genome map and histogram of SNPs were generated after running MAFFT v7.471 alignment using a modified code from Lu et al. 2020 (https://github.com/laduplessis/SARS-CoV-2_Guangdong_genomic_epidemiology/). Moreover, we identified global virus lineages using Nextclade v0.8.1 (https://clades.nextstrain.org/) and Pangolin v2 (https://github.com/cov-lineages/pangolin; Rambaut et al. 2020).

### Phylogenetics analysis

All available SARS-CoV-2 genomes (285,411 sequences) were obtained from GISAID on December 24, 2020. Available sequences were then subjected to analysis inside NextStrain ncov pipeline (https://github.com/nextstrain/ncov; Hadfield et al. 2018). Briefly, this pipeline uses the augur toolkit to (i) exclude short and low quality sequences or those with incomplete sampling date; (ii) align filtered sequences using MAFFT v7.471 (Katoh and Standley 2013); mask uninformative sites from the alignment; (iv) perform context subsampling using genetically closely-related genomes to our focal subset prioritizing sequences geographically closer to RS state, Brazil; (v) build maximum likelihood (ML) phylogenetics tree using IQ-TREE v2.0.3, employing the best-fit model of nucleotide substitution as indicated by ModelFinder (Nguyen et al. 2015); (vi) generate a time-scaled tree resolving polytomies and internal nodes with TreeTime v0.7.6, and under a strict clock under a skyline coalescent prior with a rate of 8×10^−4^ substitutions per site per year (Sagulenko et al. 2018); (vii) label clades, assign mutations and infer geographic movements; and (viii) export results to JSON format to enable interactive visualization through auspice.us. The ML tree was inspected in TempEst v1.5.3 (Rambaut et al. 2016) to investigate the temporal signal through regression of root-to-tip genetic divergence against sampling dates. No outliers were detected in this procedure.

### Phylodynamics and phylogeographic analysis

All global sequences (until December 24, 2020) belonging to lineages B.1.1.248 (n=405) and B.1.1.33 (n=725), found in high frequency in this study, were recovered from the filtered MAFFT alignment performed inside Nextstrain ncov pipeline in the previous step. The T_MRCA_ and the spatial diffusion of these important circulating lineages through Brazil were separately estimated for each lineage using a Bayesian Markov Chain Monte Carlo (MCMC) approach as implemented in BEAST v1.10.4 (Suchard et al. 2018), using the BEAGLE library v3 (Ayres et al. 2012) to save computational time. Time-scaled Bayesian trees were estimated in BEAST using: HKY+Γ nucleotide substitution model, a strict molecular clock model with a fixed substitution rate (8×10^−4^ subst/site/year) (Duchene et al. 2020) through a continuous time Markov Chain (CTMC) prior for the clock rate (Ferreira and Suchard 2008), and both a parametric exponential growth and a nonparametric skygrid priors (Gill et al. 2013).

Two MCMC chains were run for at least 120 million generations and convergence of the MCMC chains was inspected using Tracer v1.7.1 (Rambaut et al. 2018). After removal of 10% burn-in, log and tree files were combined using LogCombiner v1.10.4 (Suchard et al. 2018) to ensure stationarity and good mixing. Maximum clade credibility (MCC) summary trees were generated using TreeAnnotator v1.10.4 (Suchard et al. 2018). MCC trees were visualized using FigTree v1.4 (http://tree.bio.ed.ac.uk/software/figtree/) and additional annotations were performed in ggtree R package v2.0.4 (Yu et al. 2017).

Viral migrations across time were reconstructed using a reversible discrete asymmetric phylogeographic model (Lemey et al. 2009) in order to estimate locations of each internal node of the phylogeny. SpreaD3 (Bielejec et al. 2016) was used to map spatiotemporal information embedded in MCC trees. A discretization scheme of 10 possible states defined as Brazilian regions (Centre-West, North, Northeast, South, and Southeast) or other continents (Africa, Europe, North America, Oceania, and South America) was applied. Latitudes and longitudes were attributed to a randomly selected point next to the center of each region or continent. Location exchange rates that dominate the diffusion process were identified using the Bayesian stochastic search variable selection (BSSVS) procedure (Lemey et al. 2009) using Bayes factor tests to identify well-supported rates.

### Data access

Consensus genomes have been submitted to the GISAID database under accession numbers EPI_ISL_831474, EPI_ISL_831645, EPI_ISL_831646, EPI_ISL_831660, EPI_ISL_831678, EPI_ISL_831681, EPI_ISL_831683, EPI_ISL_831685, EPI_ISL_831688, EPI_ISL_831689, EPI_ISL_831892, EPI_ISL_831898, EPI_ISL_831913, EPI_ISL_831938, EPI_ISL_831939, EPI_ISL_831940, EPI_ISL_832013, EPI_ISL_832012, EPI_ISL_832011, EPI_ISL_832010, EPI_ISL_83209. The publicly available sequences used are acknowledged in the Supplemental File 3.

## Data Availability

Consensus genomes have been submitted to the GISAID database under accession numbers EPI_ISL_831471, EPI_ISL_831474, EPI_ISL_831645, EPI_ISL_831646, EPI_ISL_831660, EPI_ISL_831678, EPI_ISL_831681, EPI_ISL_831683, EPI_ISL_831685, EPI_ISL_831688, EPI_ISL_831689, EPI_ISL_831892, EPI_ISL_831898, EPI_ISL_831913, EPI_ISL_831938, EPI_ISL_831939, EPI_ISL_831940, EPI_ISL_832013, EPI_ISL_832012, EPI_ISL_832011, EPI_ISL_832010, EPI_ISL_83209. The publicly available sequences used are acknowledged in the Supplemental File 3. All data is available on the manuscript and supplementary files.

## Acknowledgements

We thank the Mayor’s Office, Health Department and São Camilo Hospital (Esteio, RS, Brazil), Leonardo Duarte Pascoal and Ana Regina Boll for their work in combating Covid-19, for providing the samples and epidemiological information of patients, and for the financial support for the sequencing; the Molecular Microbiology Laboratory of Feevale University for conducting the sequencing; Arnaldo Zaha, Macley Silva Cardoso, Maiara Monteiro Oliveira, Marilene Henning Vainstein, and Augusto Schrank for writing corrections and insightful discussions on the best way to present the data of this manuscript.

## Author contributions

CET and LK conceived the study. AMM, CAMN, GDC collected clinical samples and metadata. AMM, AWH, FRS, GDC, JDF, JSG, MD, MNW, PRA generated sequencing data. CET, GBC, LK, VBF contributed bioinformatics tools. CET, GBC, LK, PAGF, VBF performed the data analysis. CET, GBC, LK, RAZ, VBF wrote the manuscript.

