## Supplemental for "Genomic Epidemiology of SARS-CoV-2 in Esteio, Rio Grande do Sul, Brazil"

### Table of Contents


**Table of Contents** **1**

Supplemental Table 1. Mutations of genomes included in this study observed with frequency > 1. 2

Supplemental Figure 1. Brazilian circulating lineages over time (February-December, 2020). 4

Supplemental Figure 2. Occurence of (A) B.1.1.48 and (B) B.1.1.33 lineages around the world over time. 5

Supplemental Figure 3. Occurrence of the most prevalent lineages from this study (B.1.1.248 and B.1.1.33) around the world, and from a rare lineage that have been identified (B.1.1.49). 6

Supplemental Figure 5. Root-to-tip regression of genetic divergence against sampling dates. Sequences from this study are highlighted in blue. 8

**Supplemental Table 1**. Mutations of genomes included in this study observed with frequency > 1.

| Genomic change | Effect | Amino acid change | Gene/region | Product | no. samples | Frequency (%) |
|---|---|---|---|---|---|---|
| C147T | NA | NA | 5' UTR | NA | 2/21 | 9.5 |
| **C241T** | | | | | **19/21** | **90.5** |
| C850T | Synonymous | G195 | ORF1ab | nsp2 | 2/21 | 9.5 |
| A2276G | Missense | I671V | | nsp2 | 2/21 | 9.5 |
| **C3037T** | **Synonymous** | **F924** | | **nsp3** | **17/21** | **81.0** |
| T3766C | Synonymous | D1167 | | nsp3 | 2/21 | 9.5 |
| **C12053T** | **Missense** | **L3930F** | | **nsp7** | **8/21** | **38.1** |
| A12964G | Synonymous | G4233 | | nsp9 | 2/21 | 9.5 |
| T13804C | Synonymous | N4513 | | RdRp | 2/21 | 9.5 |
| **C14408T** | **Synonymous** | **L4715** | | **RdRp** | **18/21** | **85.7** |
| C15654T | Missense | T5130I | | RdRp | 2/19 | 9.5 |
| C18252T | Missense | T5996I | | 3'-to-5' exonuclease | 2/19 | 9.5 |
| C20016T | Missense | T6584I | | endoRNAse | 2/19 | 9.5 |
| G23012A | Missense | E484K | S | Surface glycoprotein | 2/19 | 9.5 |
| **A23403G** | **Missense** | **D614G** | | | **19/21** | **90.5** |
| **G25088T** | **Missense** | **V1176F** | | | **8/21** | **38.1** |
| C25207T | Synonymous | Y1215 | | | 2/21 | 9.5 |
| G25429T | Missense | V13L | ORF3a | ORF3a protein | 4/21 | 19.0 |
| C25509A | Synonymous | A39 | | | 4/21 | 19.0 |
| C25642T | Synonymous | L84 | | | 2/21 | 9.5 |
| A26019T | Synonymous | S209 | | | 2/21 | 9.5 |
| **T27299C** | **Missense** | **I33T** | **ORF6** | **ORF6 protein** | **9/21** | **42.9** |
| A27976G | Missense | H28R | ORF8 | ORF8 protein | 4/21 | 19.0 |
| C28093T | Missense | S67F | | | 2/21 | 9.5 |
| C28253T | Synonymous | F120 | | | 2/21 | 9.5 |
| T28393C | Synonymous | R40 | N | Nucleocapsid phosphoprotein | 2/21 | 9.5 |
| G28628T | Missense | A119S | | | 2/21 | 9.5 |
| **G28881A** | **Missense** | RG203-204KR | | | **20/21** | **95.2** |
| **G28882A** | | | | | **20/21** | **95.2** |
| **G28883C** | | | | | **21/21** | **100.0** |
| G28975T | Missense | M234I | | | 2/21 | 9.5 |
| **T29148C** | **Missense** | **I292T** | | | **10/21** | **47.6** |

Original bases or amino acids are represented before the genome coordinate, while the mutated ones are presented after. SNPs observed in more than 5 sequences are highlighted in bold. UTR= Unstranslated region; ORF=Open reading frame; S: Spike; N: Nucleocapsid; nsp: nonstructural protein; RdRp: RNA-dependent RNA polymerase.

**Supplemental Figure 1**. Brazilian circulating lineages over time (February-December, 2020).

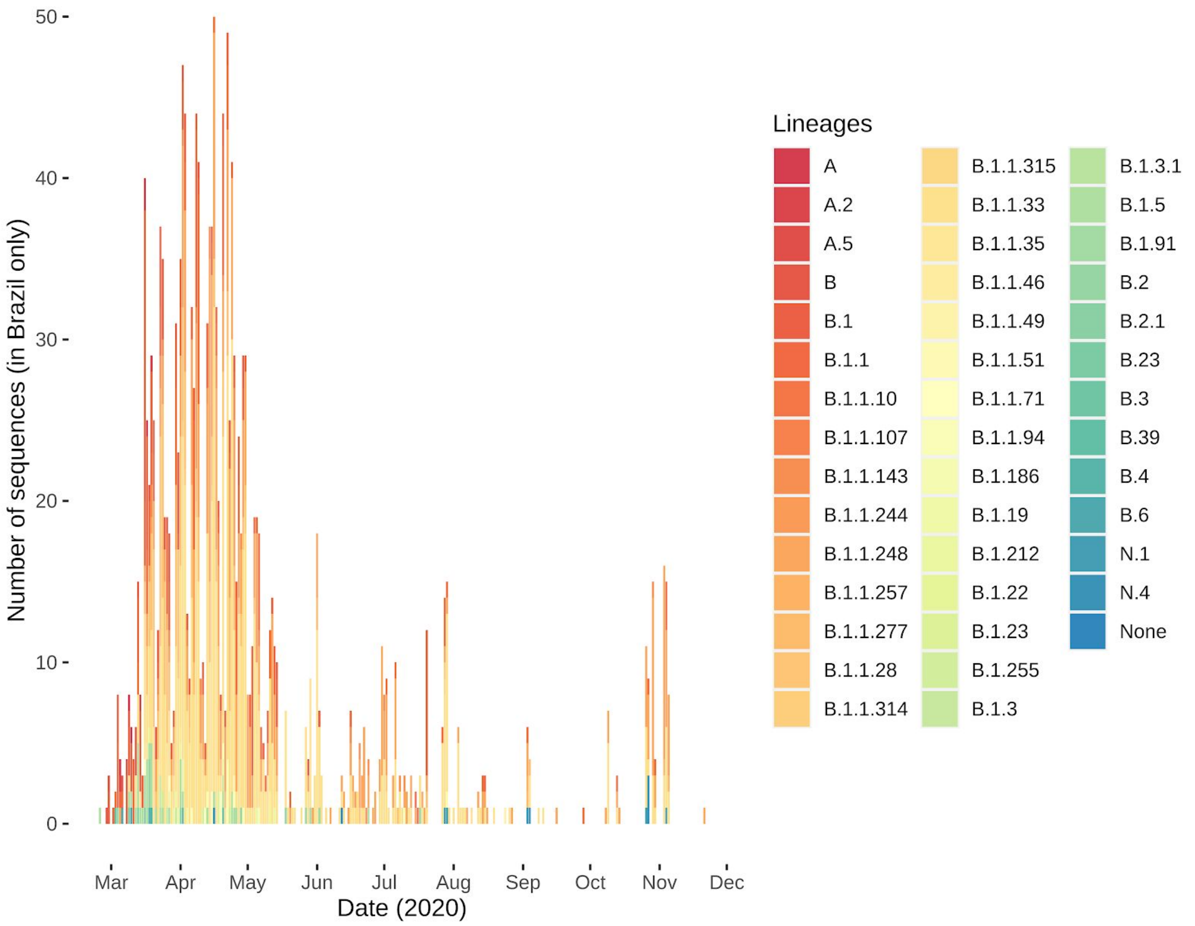

**Supplemental Figure 2**. Occurence of (A) B.1.1.48 and (B) B.1.1.33 lineages around the world over time.

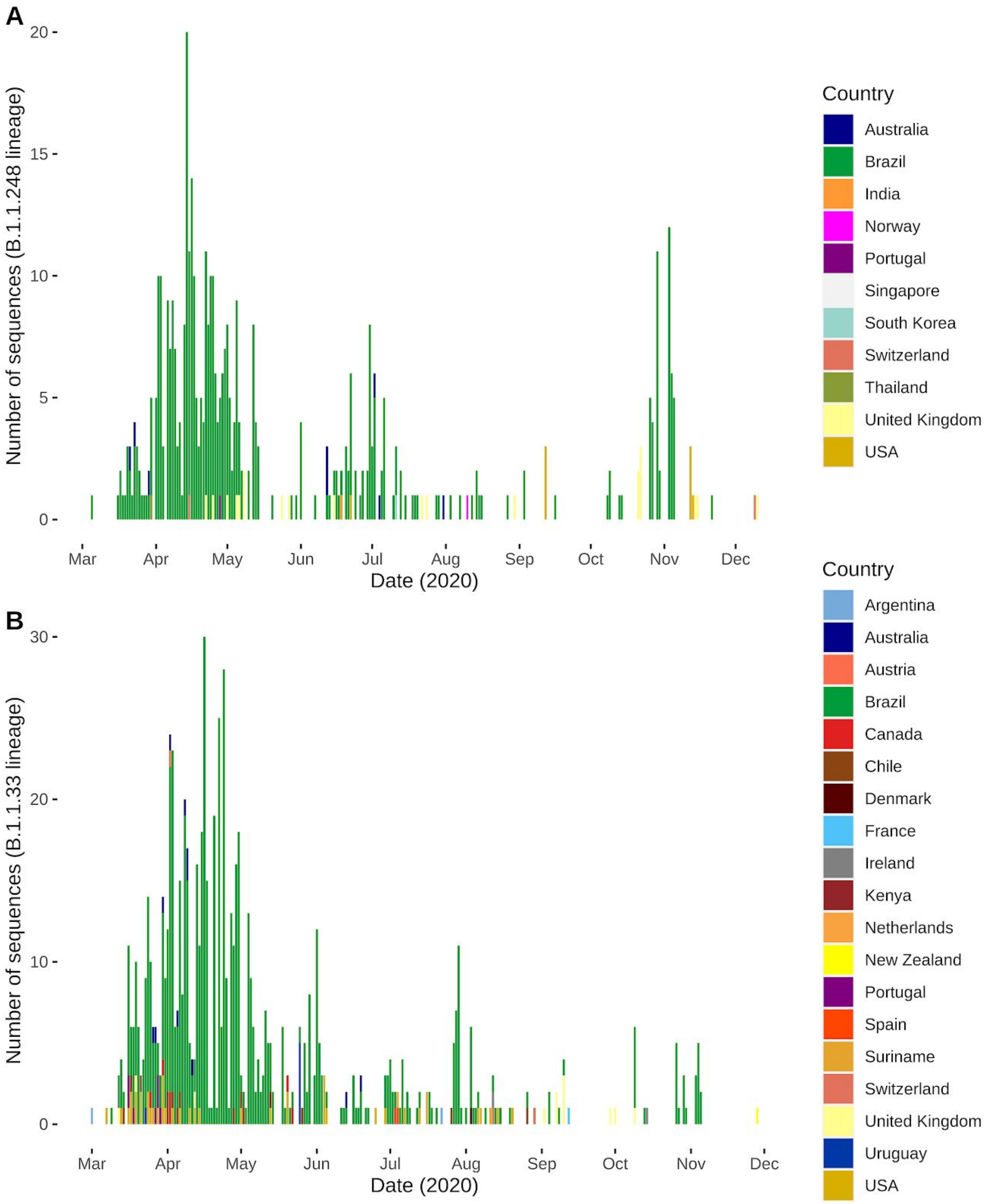

**Supplemental Figure 3**. Occurrence of the most prevalent lineages from this study (B.1.1.248 and B.1.1.33) around the world, and from a rare lineage that have been identified (B.1.1.49).

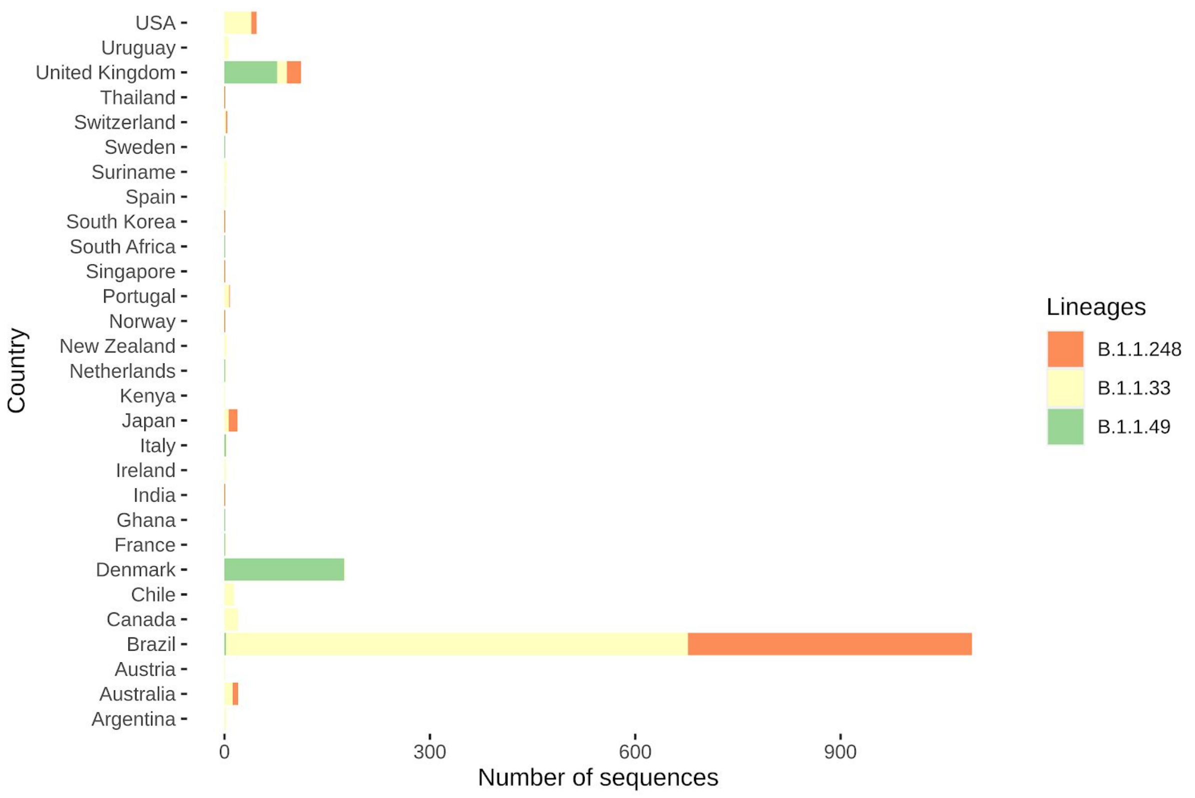

**Supplemental Figure 4.** Time-, geographical- and genomic representative tree generated after subsampling in the Nextstrain ncov pipeline, with tips colored by region of origin. The ring represents global phylogenetic lineages inferred using pangolin ([https://github.com/hCoV-2019/pangolin](https://github.com/hCoV-2019/pangolin)), and legend was ordered by lineage frequency in the tree.

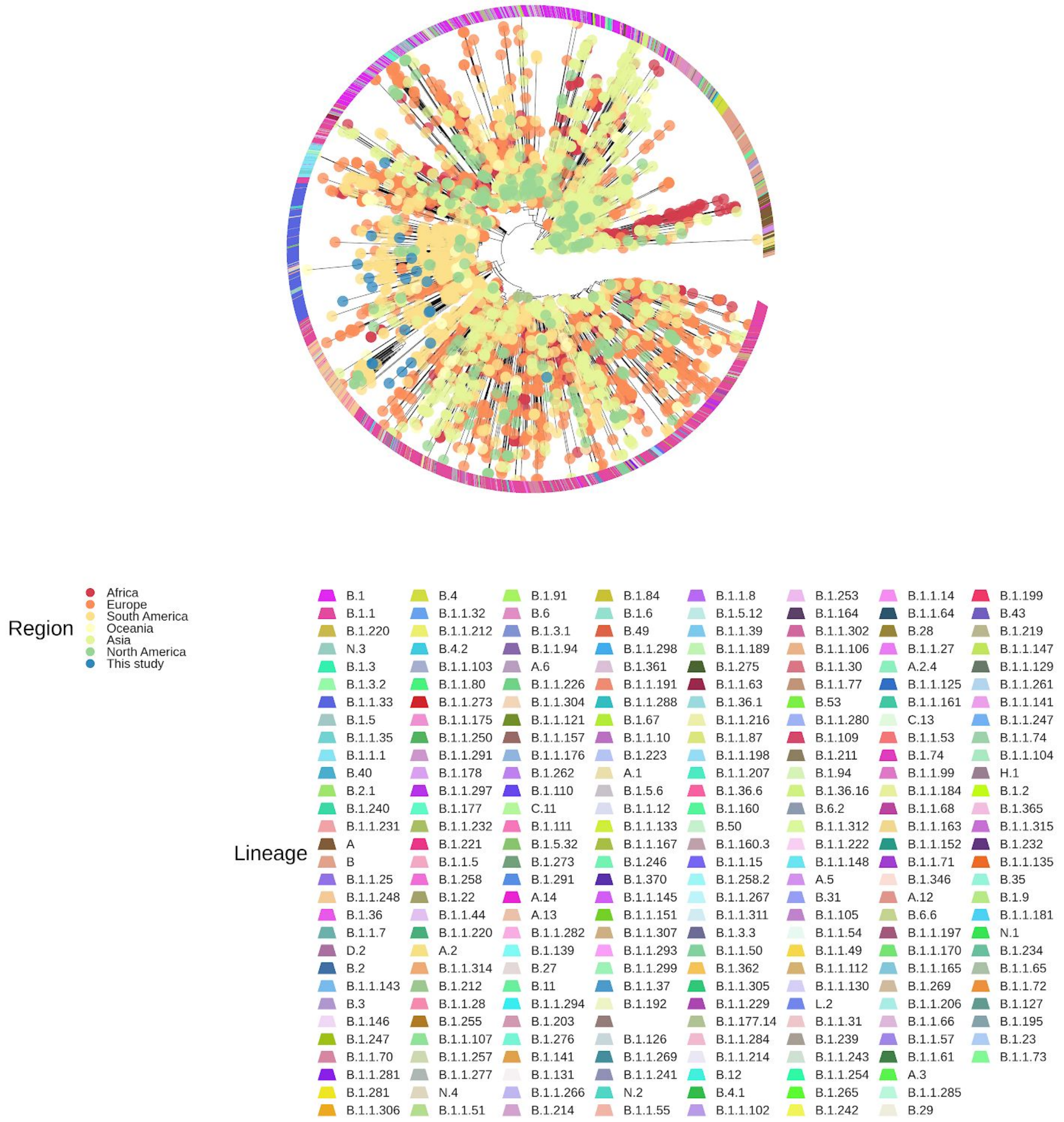

**Supplemental Figure 5.** Root-to-tip regression of genetic divergence against sampling dates. Sequences from this study are highlighted in cyan.

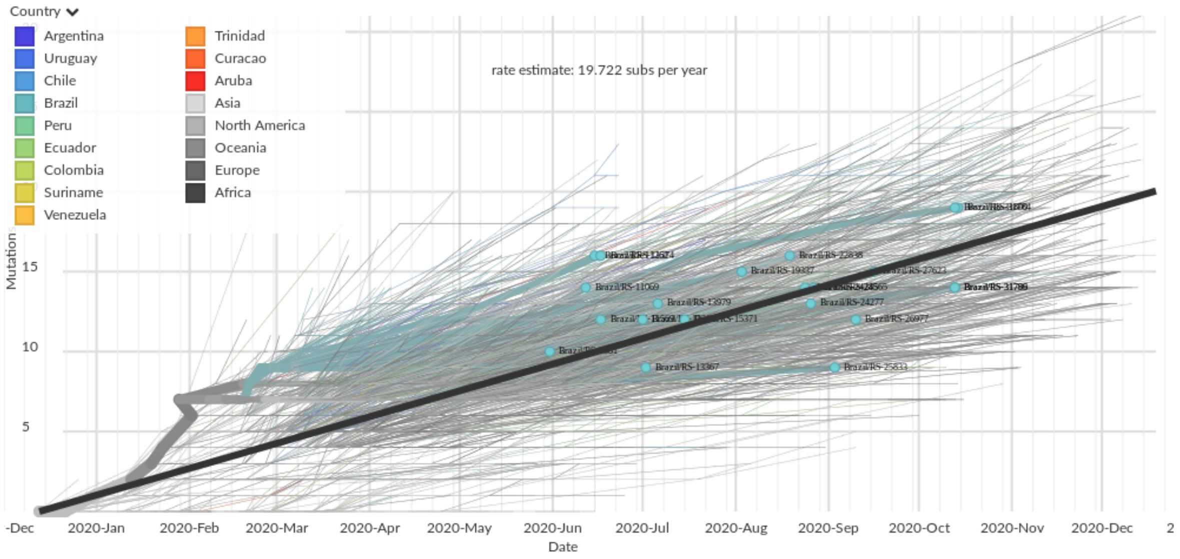
