## Supplemental File 1 for "Genomic Epidemiology of SARS-CoV-2 in Esteio, Rio Grande do Sul, Brazil"

#### Sample 1

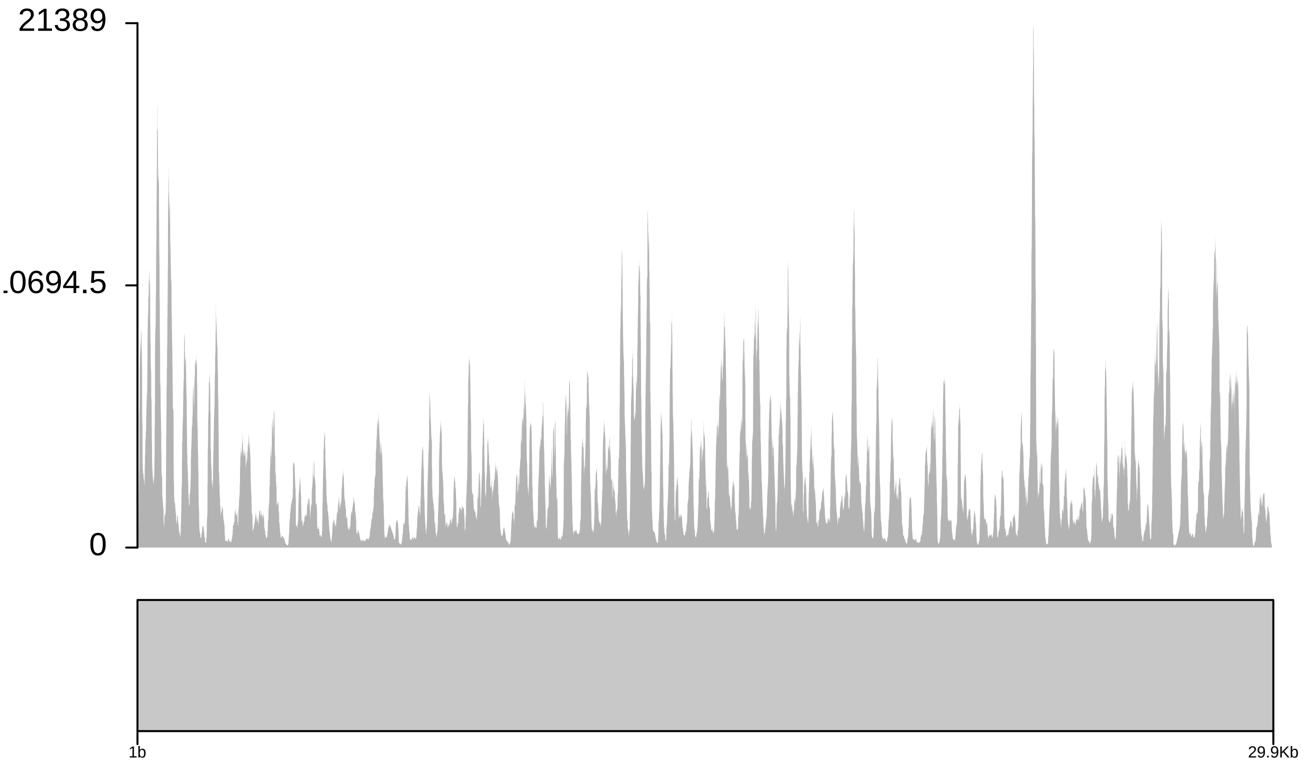

Sample 2

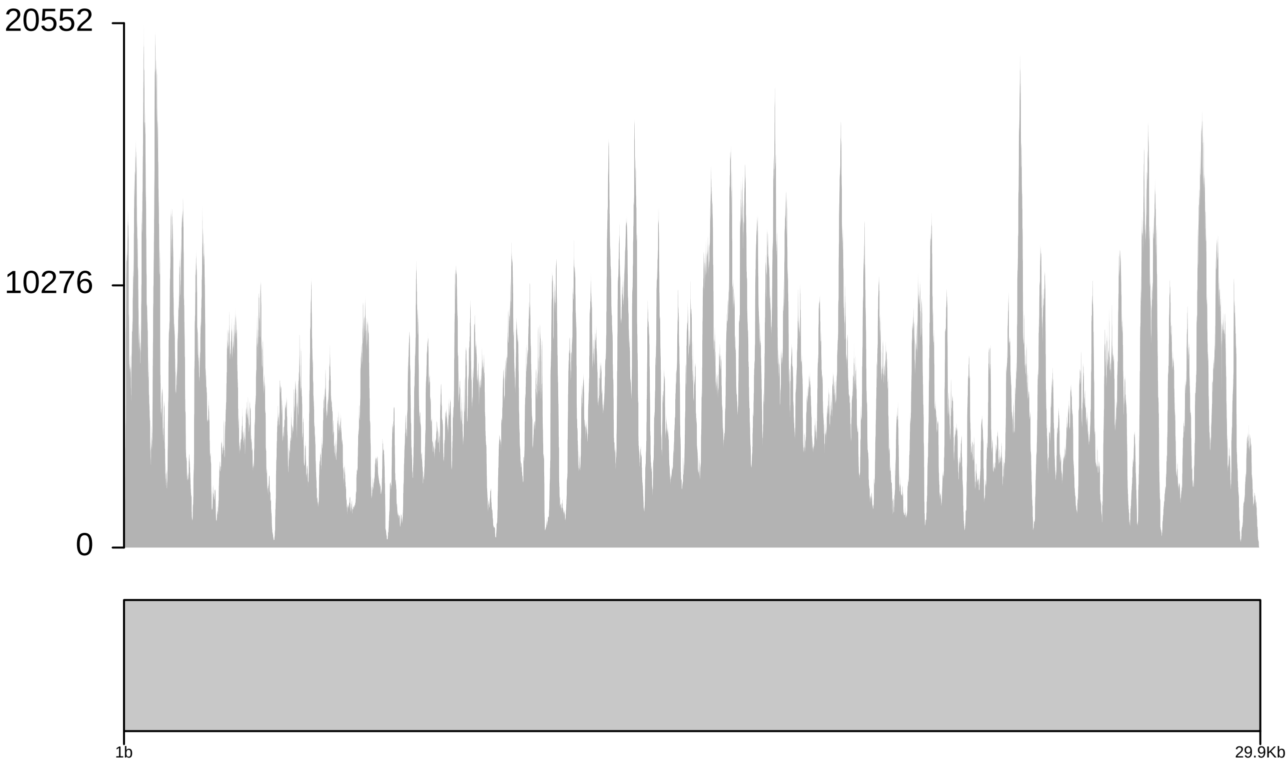

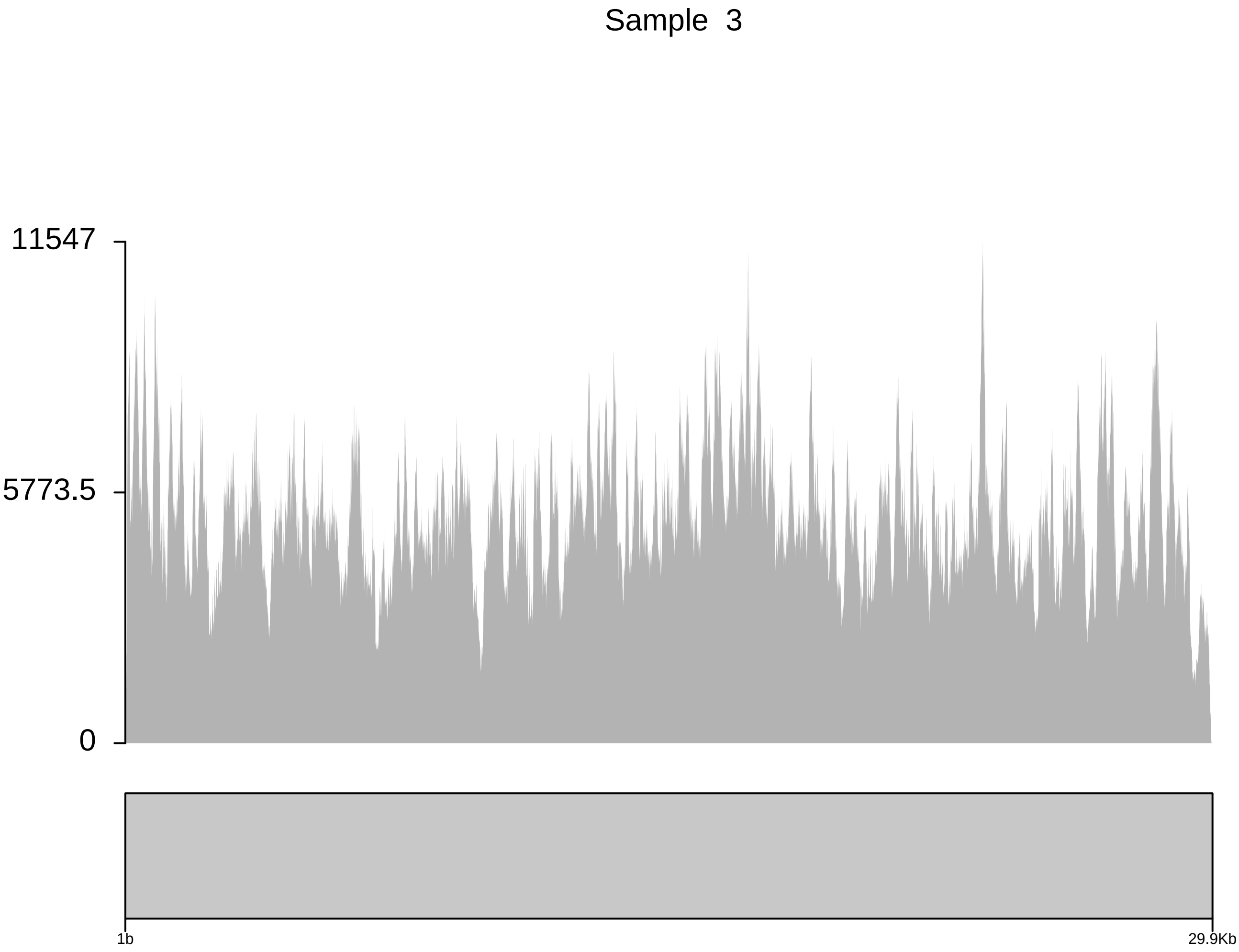
Sample 3
11547
5773.5
0
1b
29.9Kb

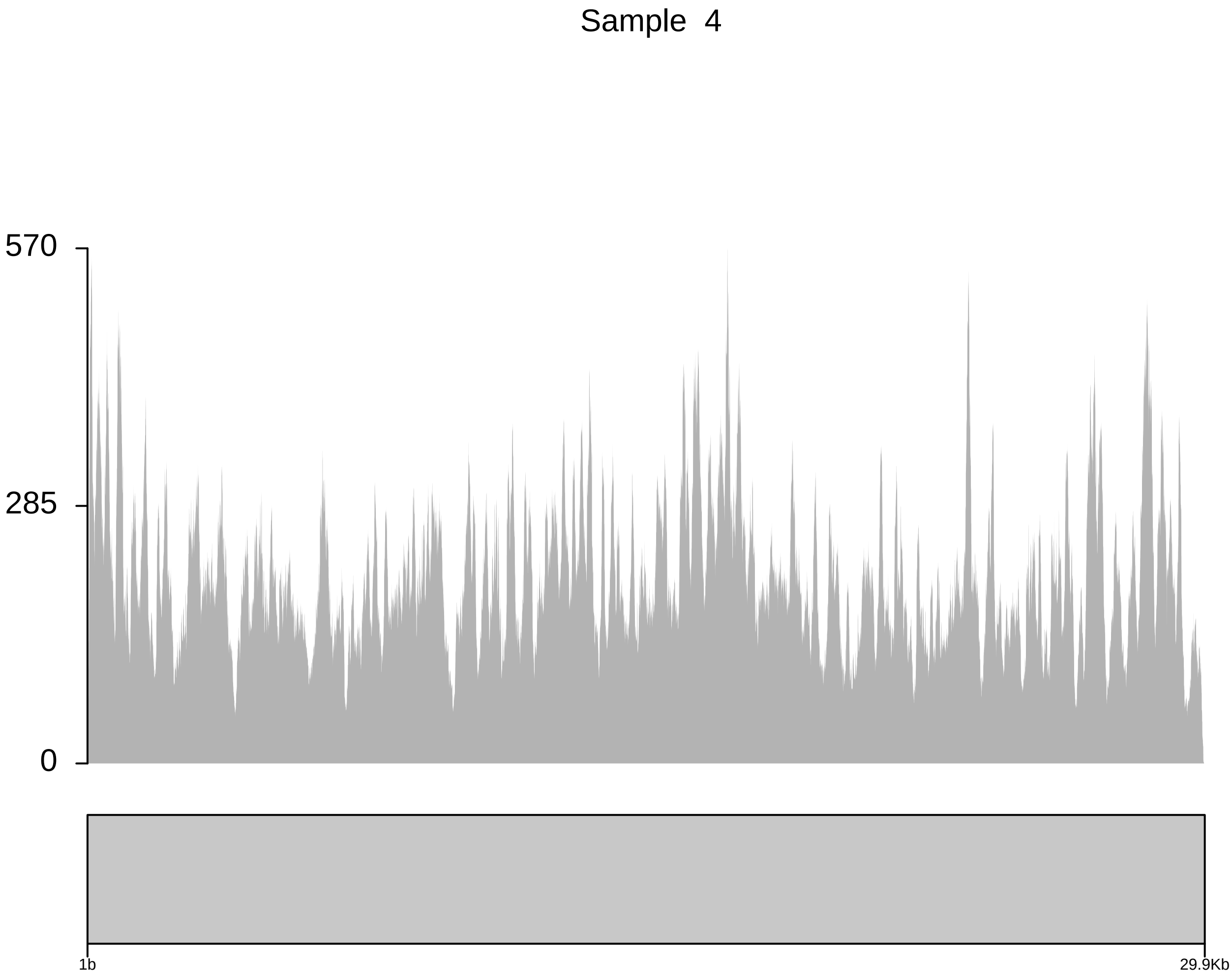

Sample 4
570
285
0
1b
29.9Kb

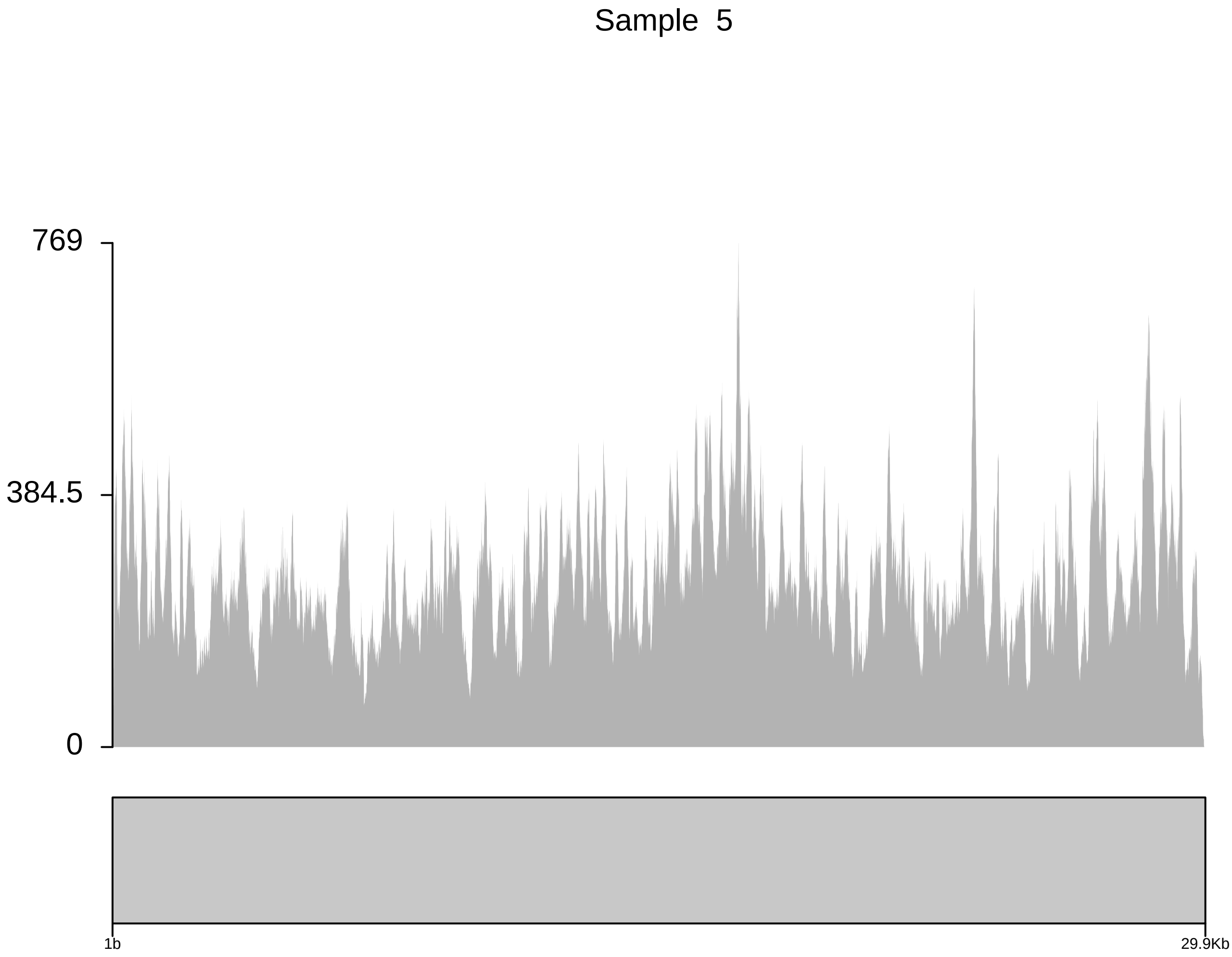
Sample 5
769
384.5
0
1b
29.9Kb

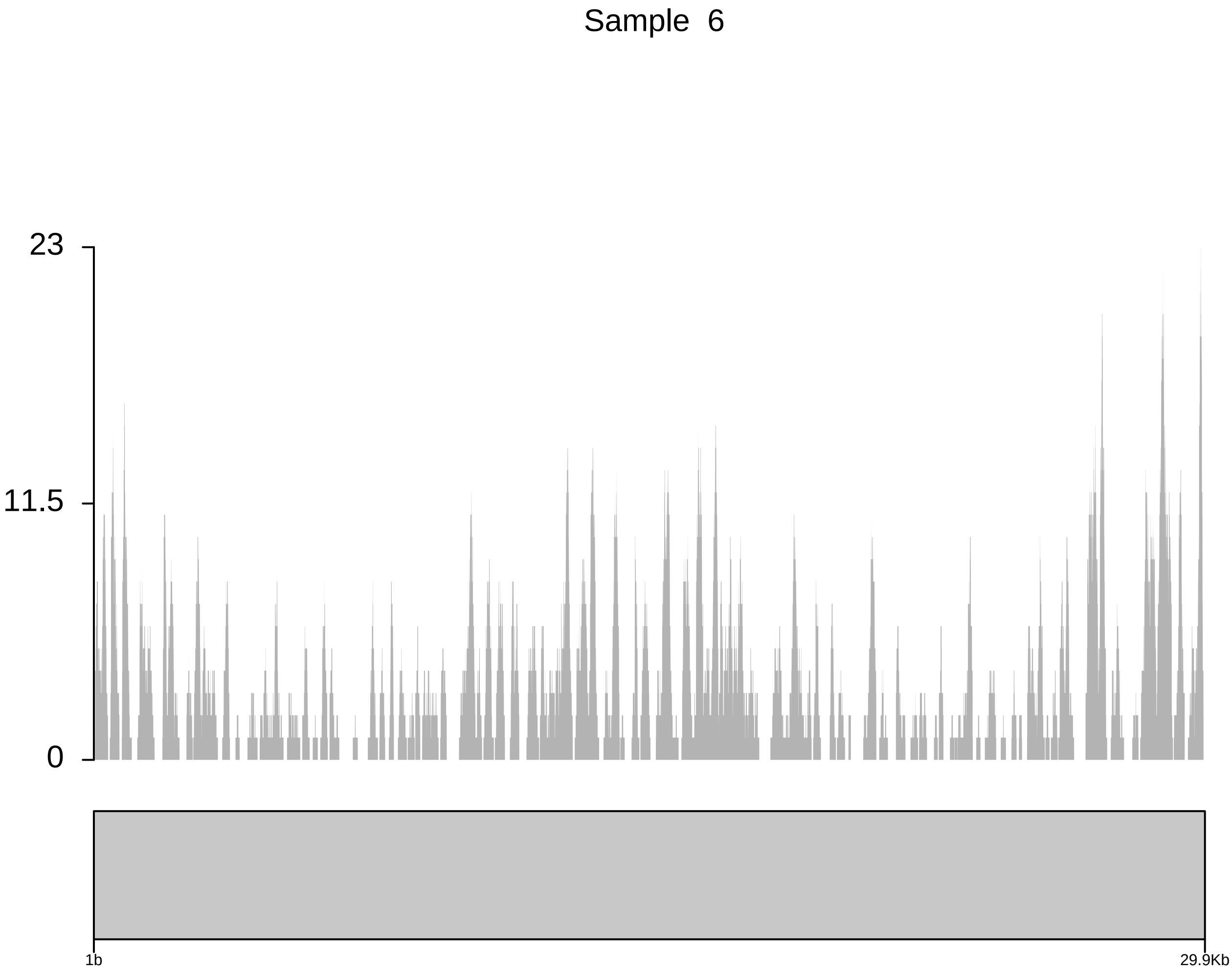
Sample 6
23
11.5
0
1b
29.9Kb

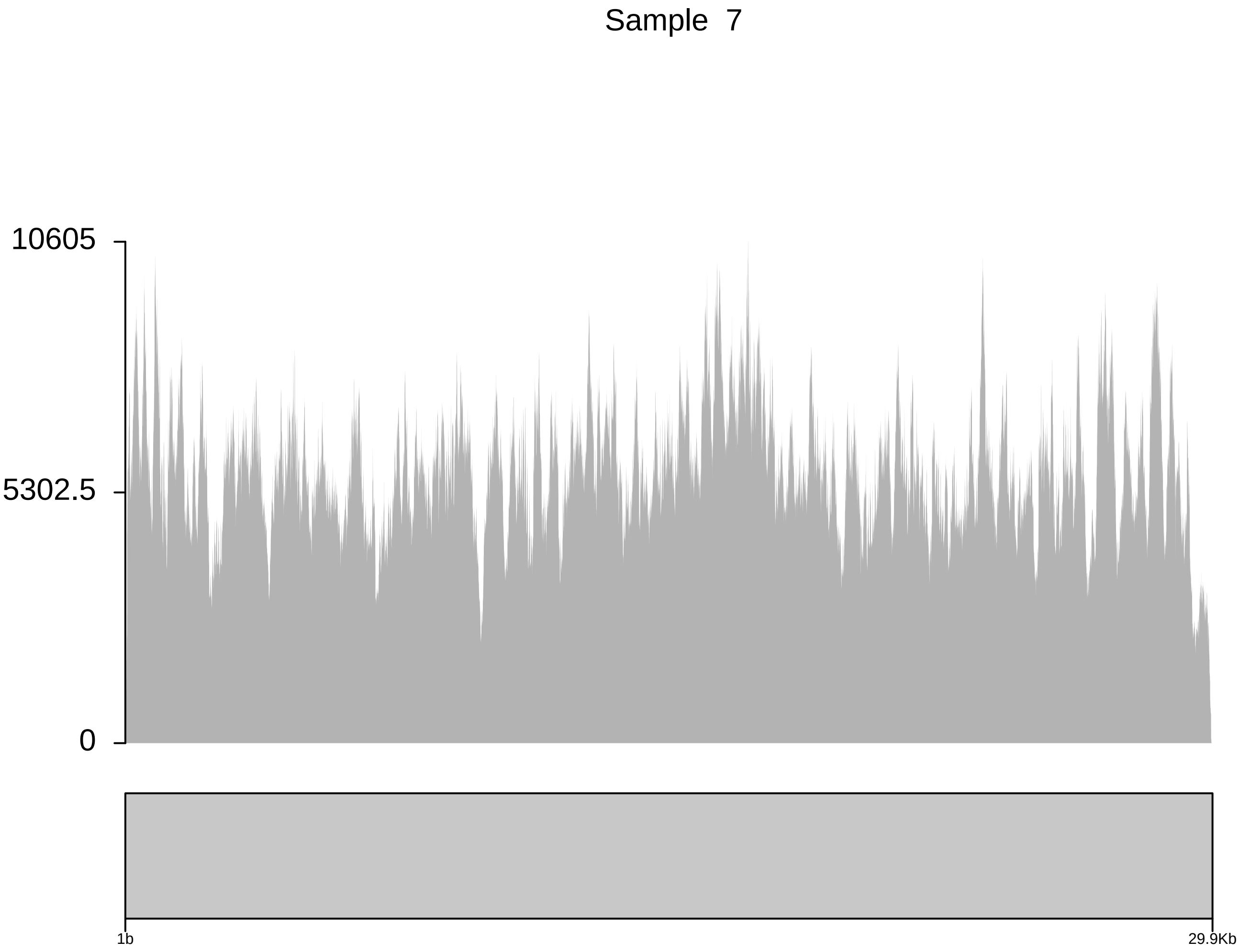

Sample 7
10605
5302.5
0
1b
29.9Kb

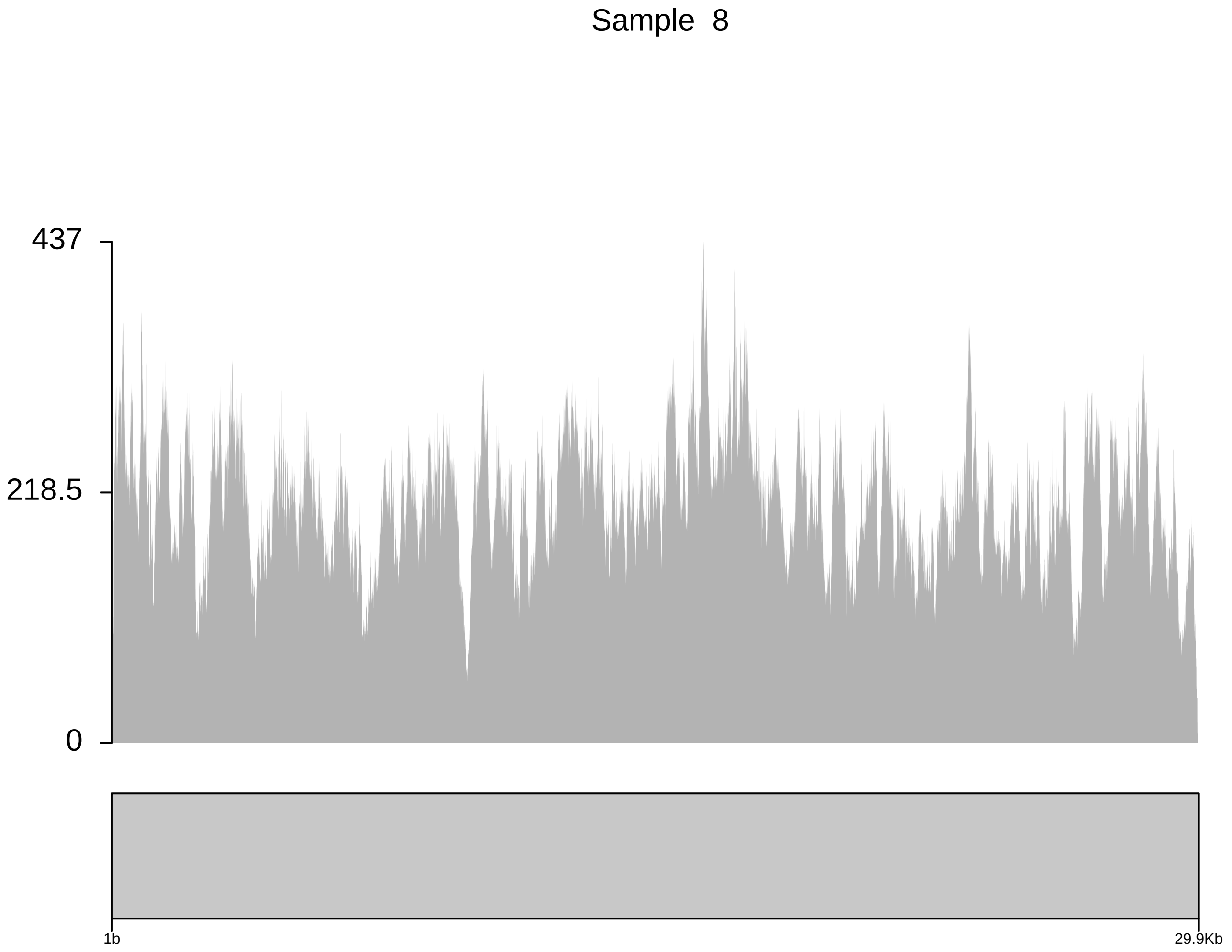
Sample 8
437
218.5
0
1b
29.9Kb

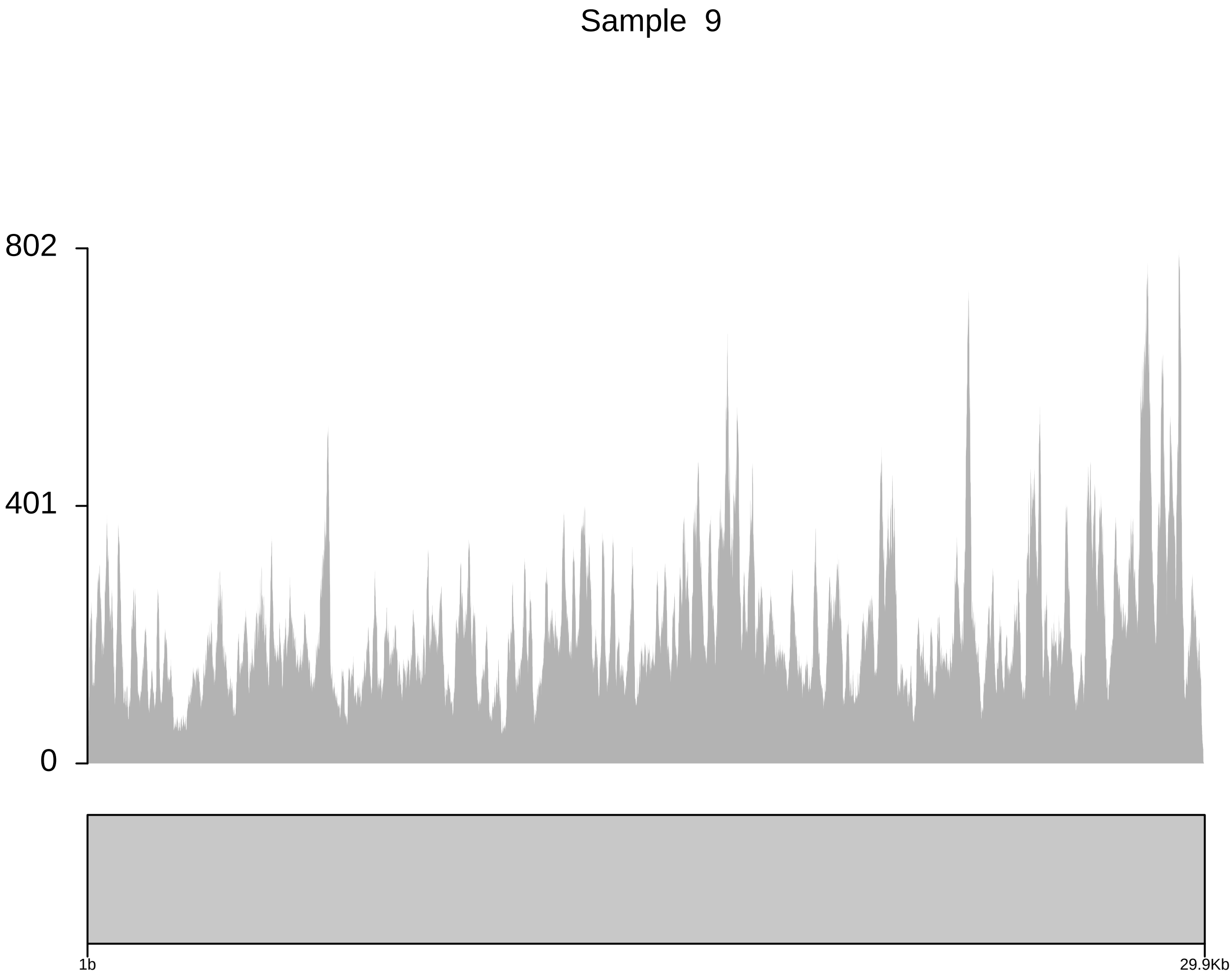

Sample 9
802
401
0
1b
29.9Kb

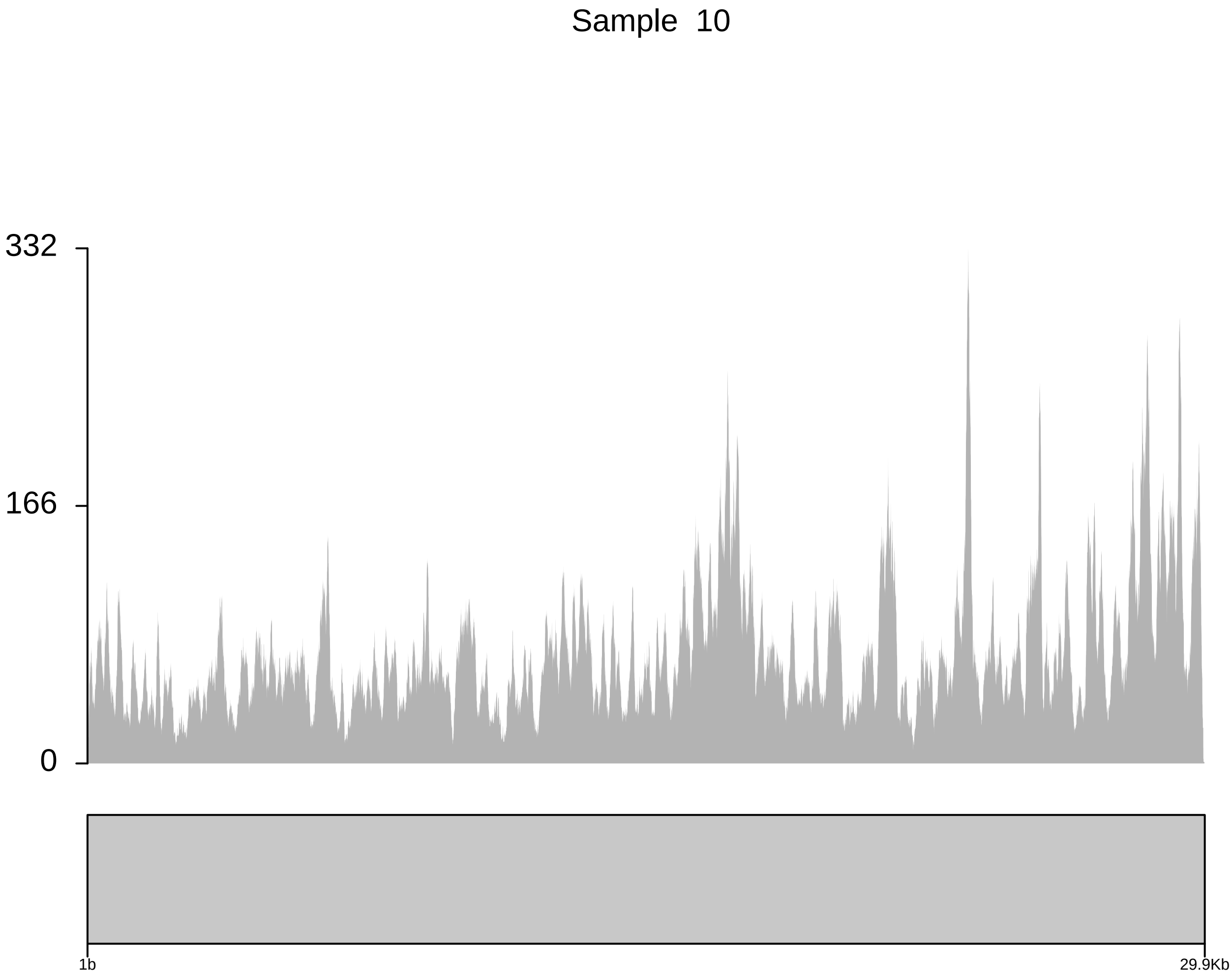
Sample 10
332
166
0
1b
29.9Kb

Sample 11

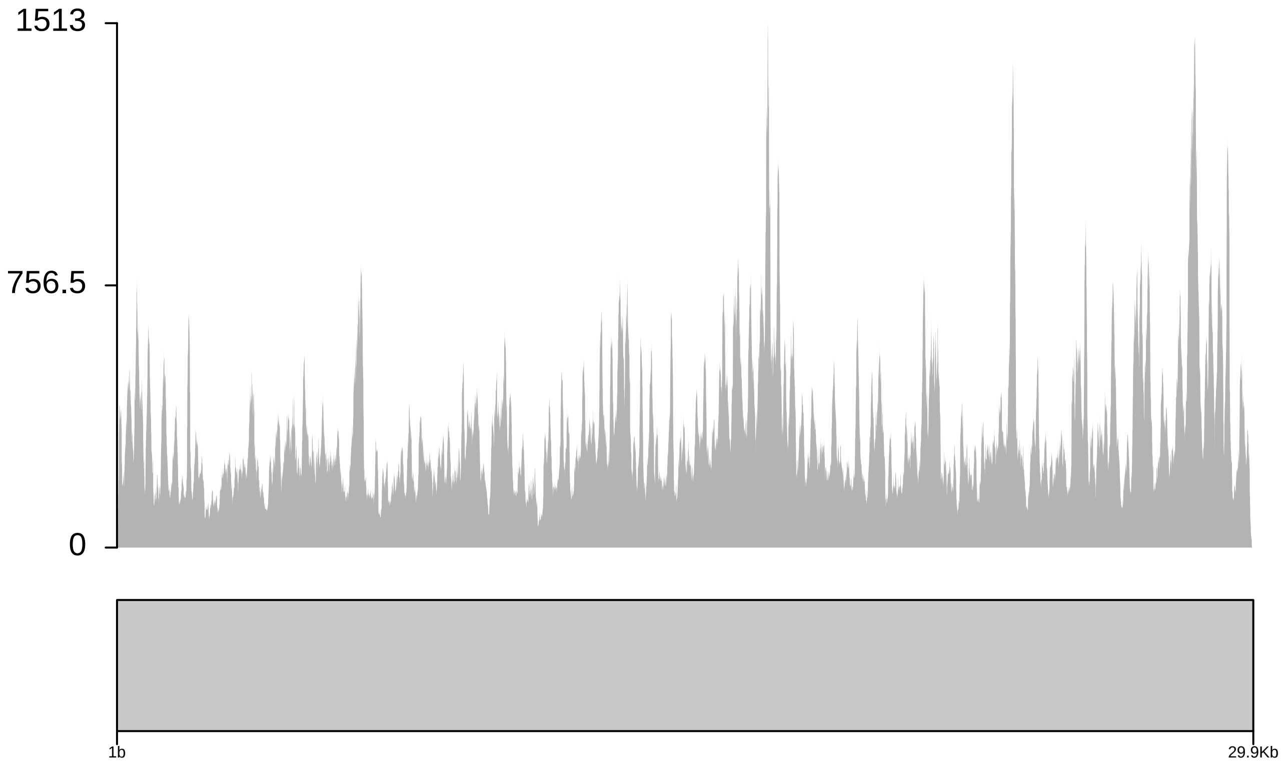

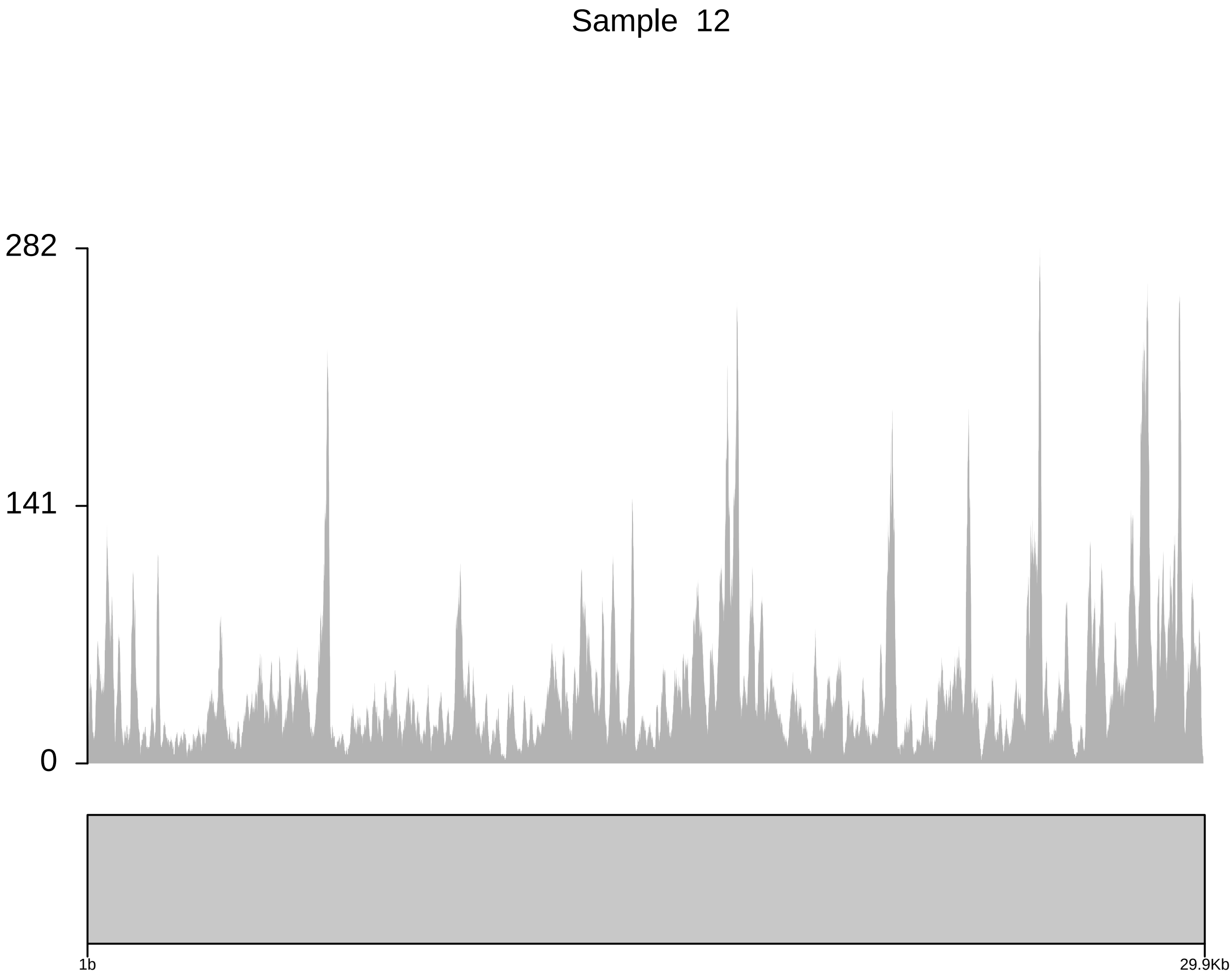
Sample 12
282
141
0
1b
29.9Kb

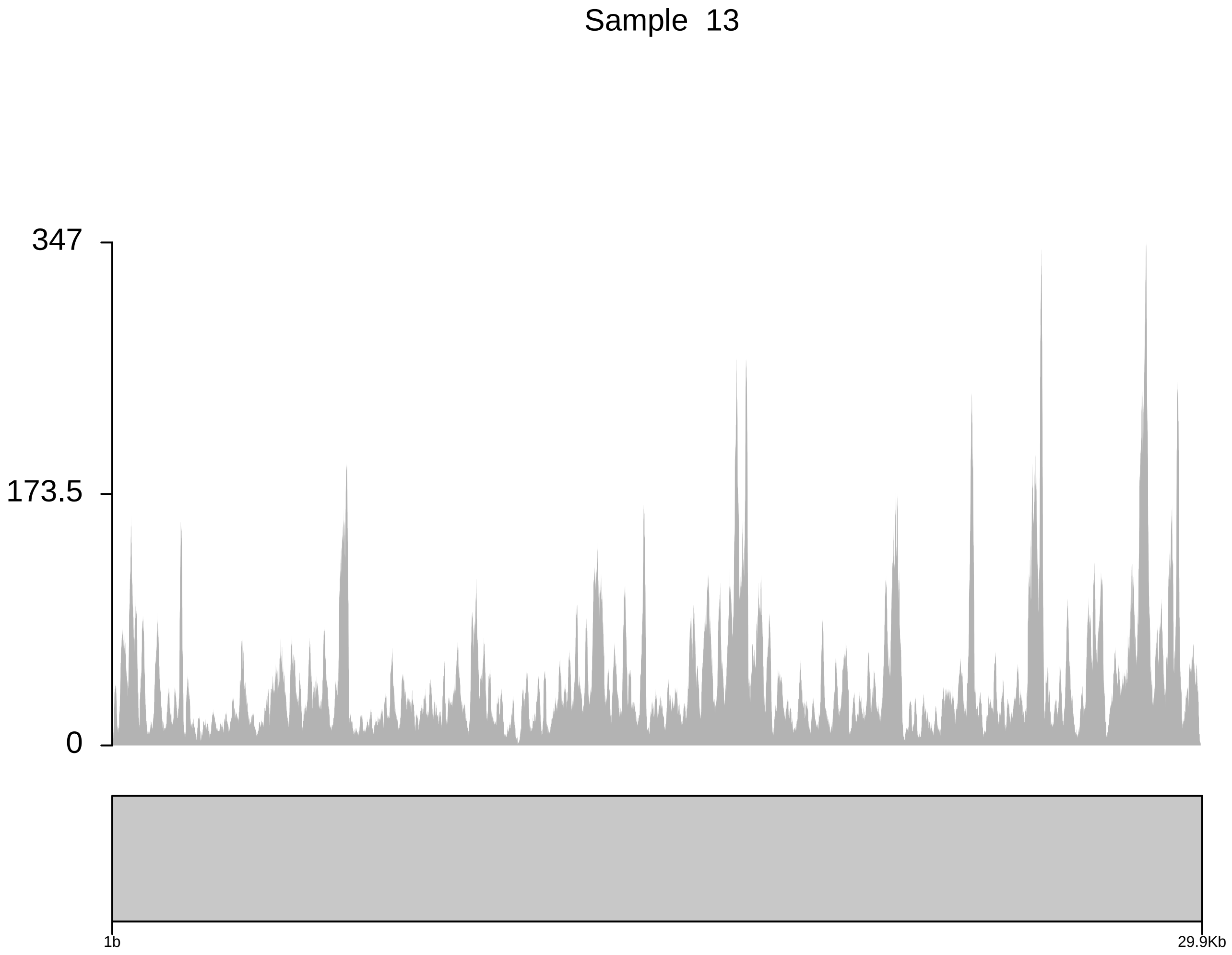
Sample 13
347
173.5
0
1b
29.9Kb

### Sample 14

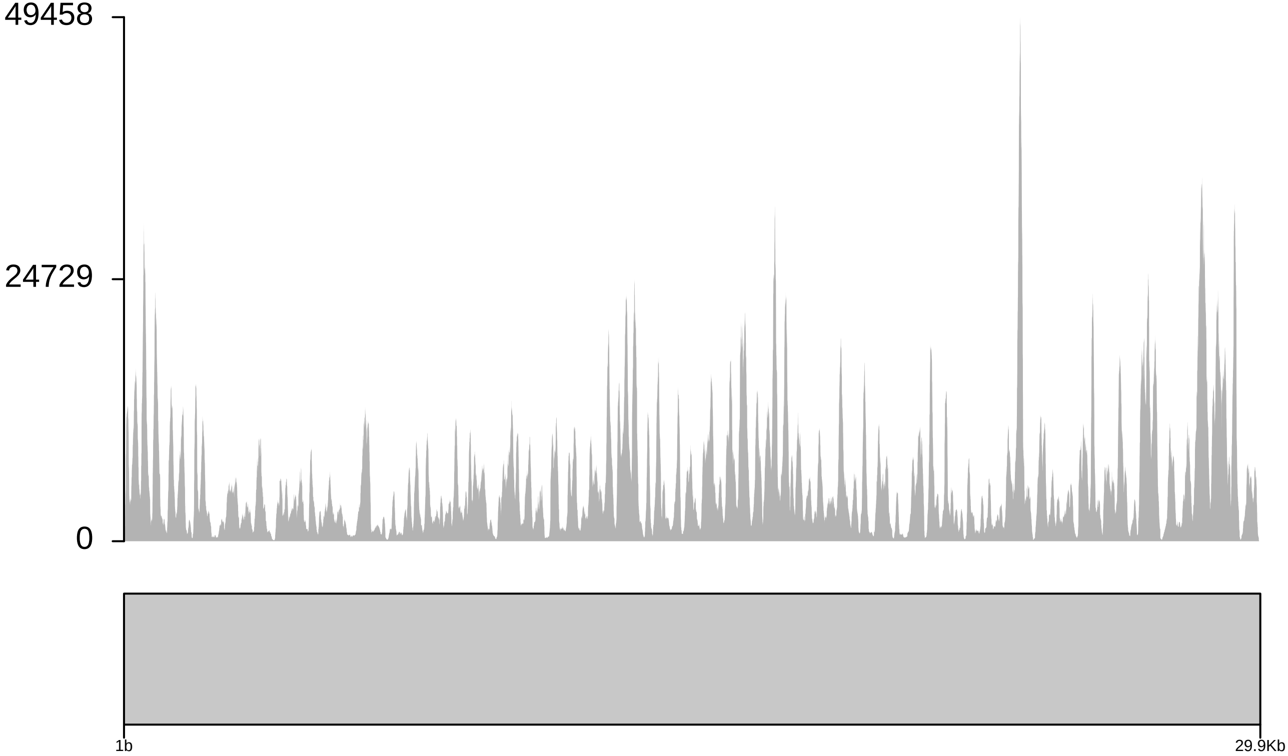

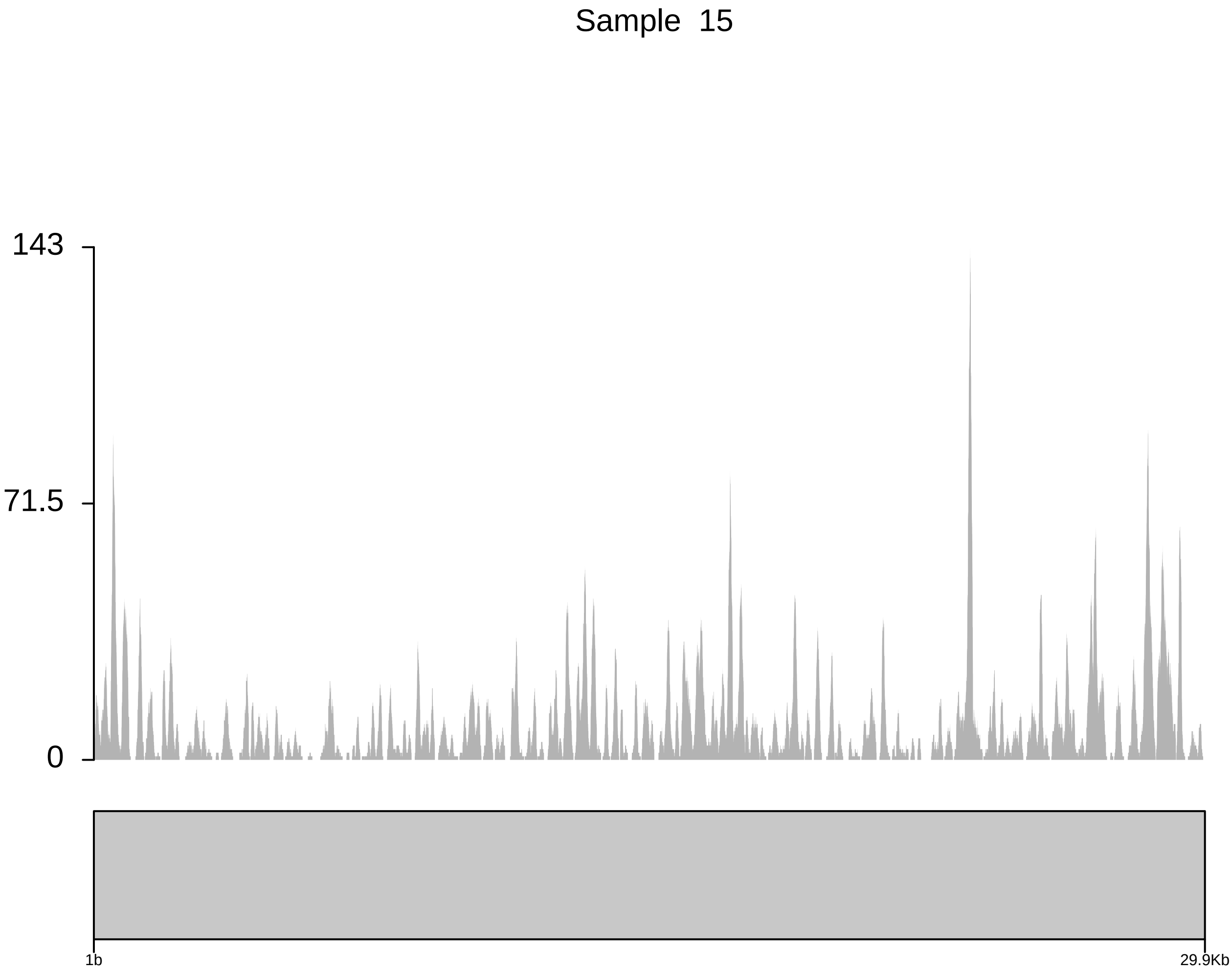
Sample 15
143
71.5
0
1b
29.9Kb

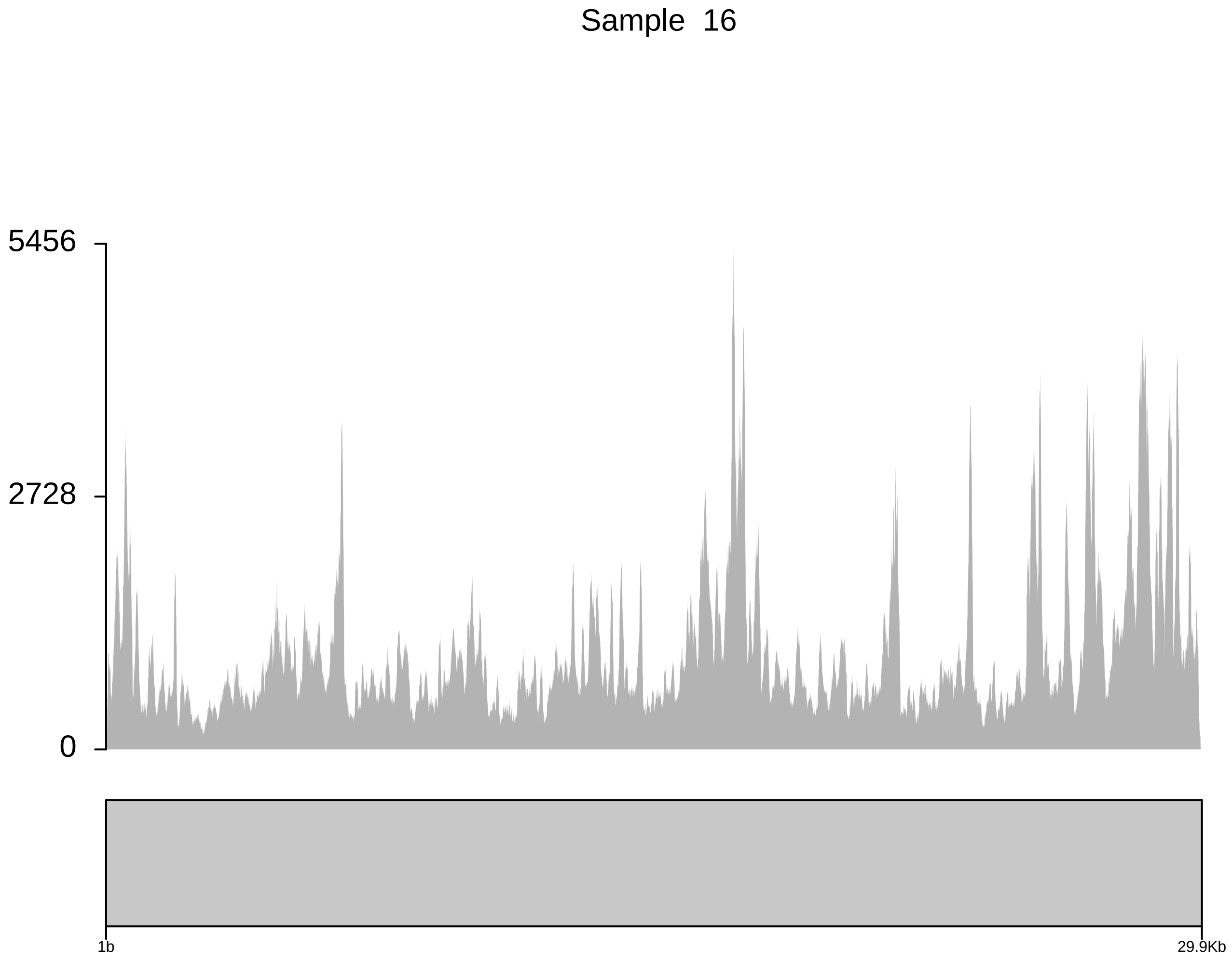
Sample 16
5456
2728
0
1b
29.9Kb

Sample 17

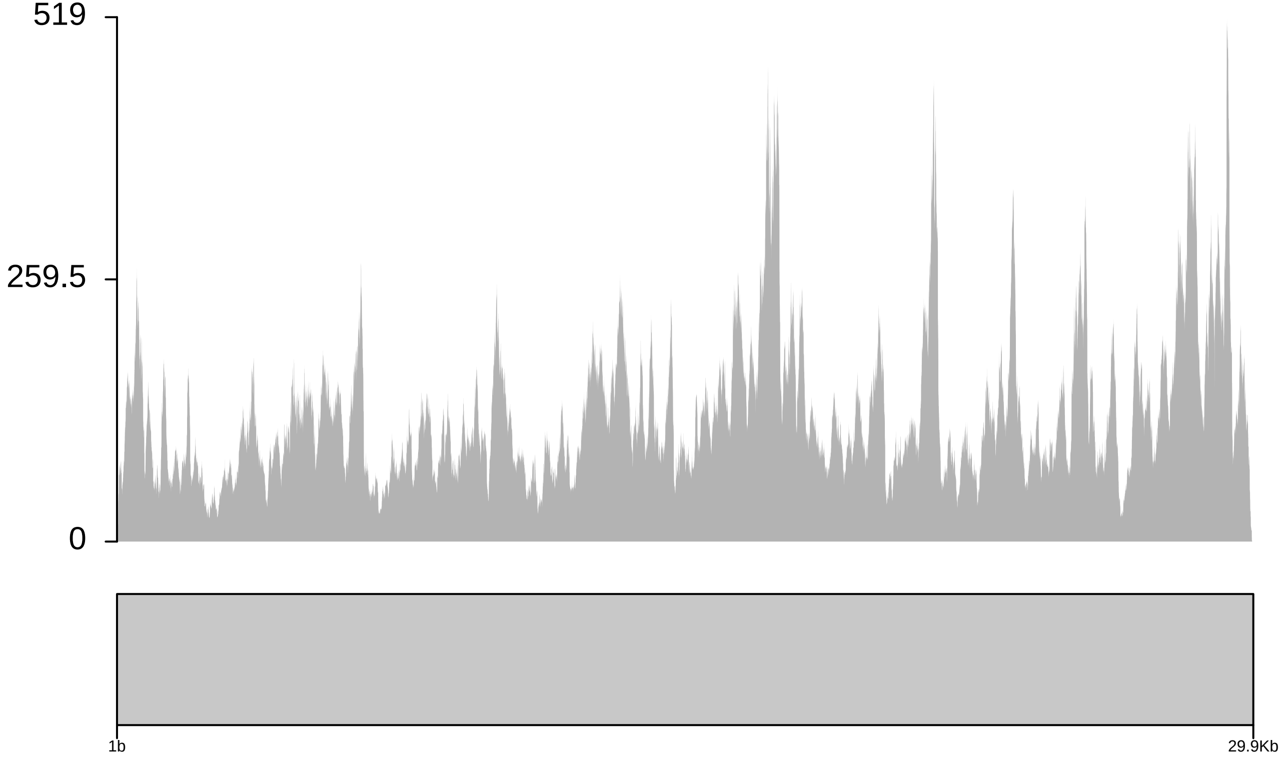

#### Sample 18

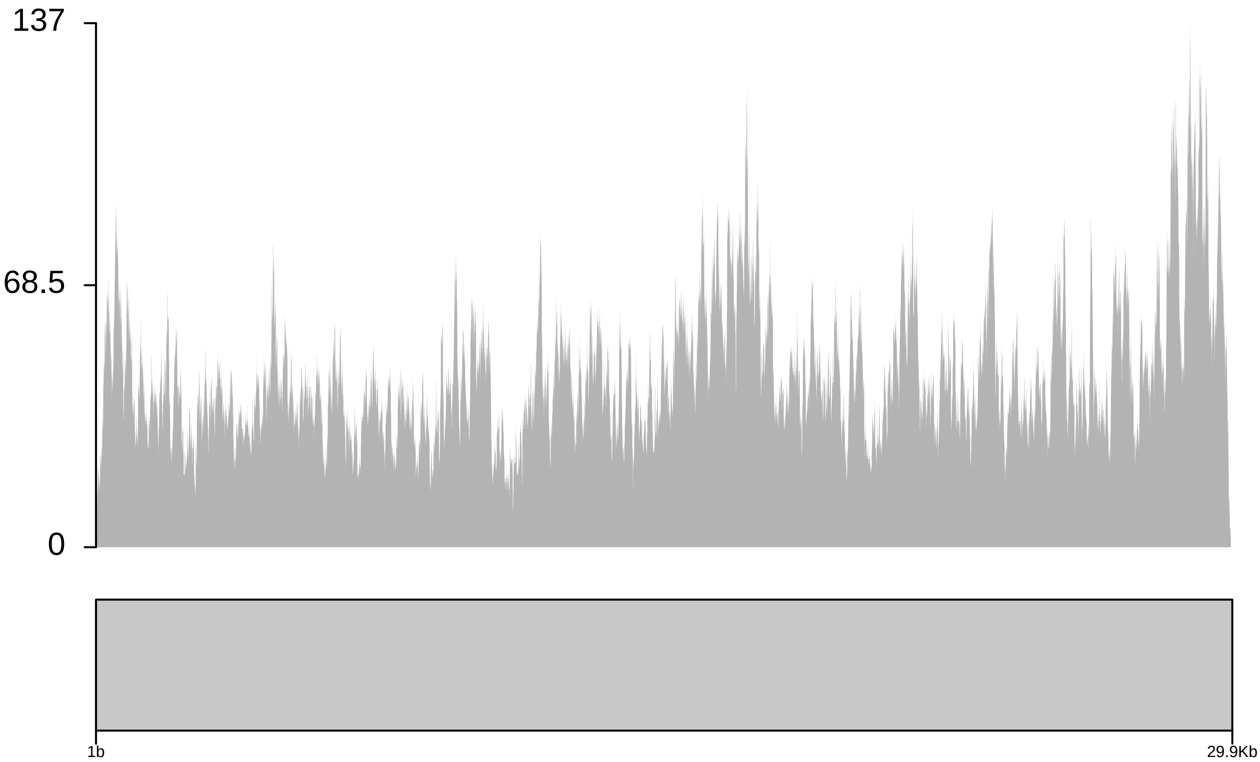

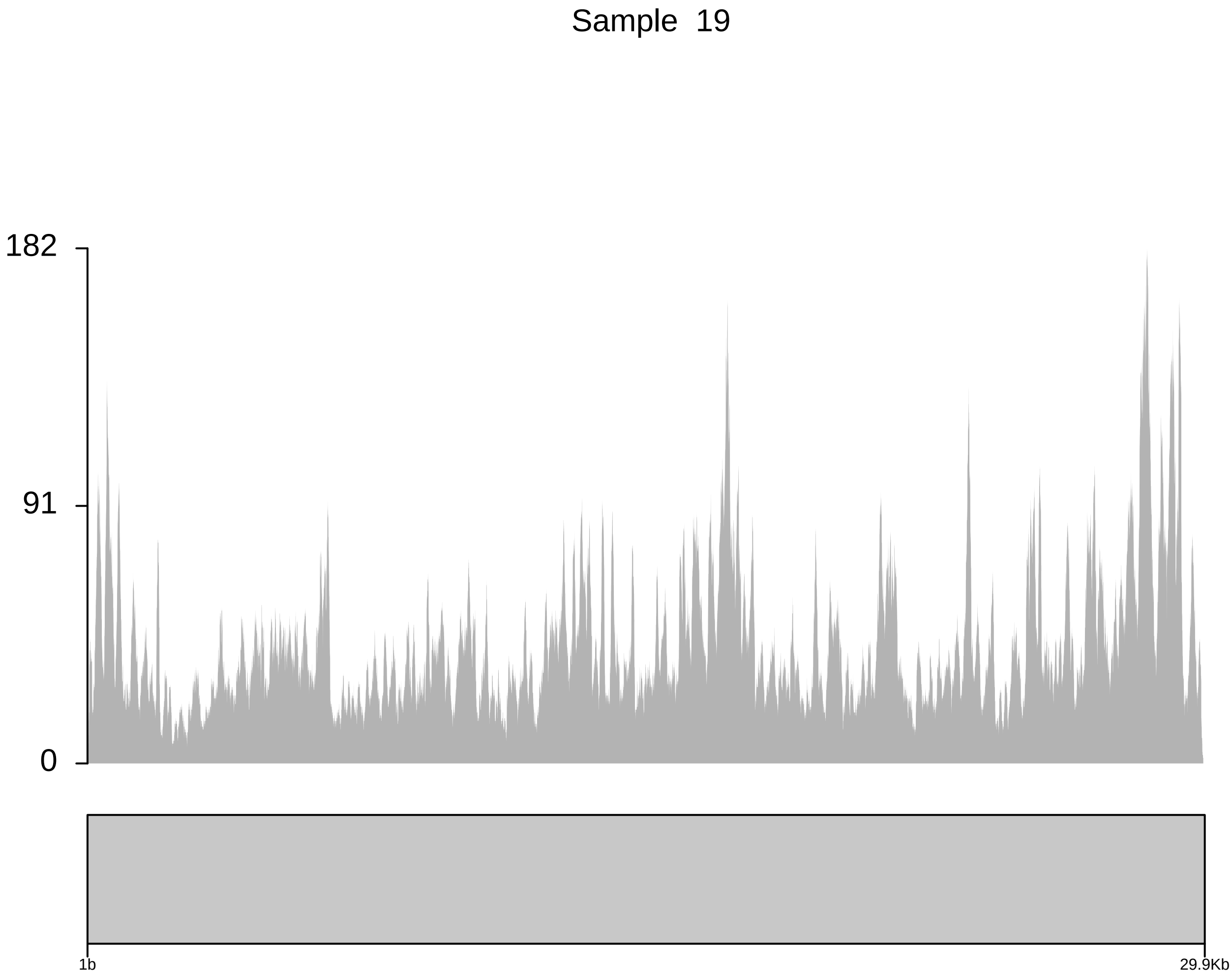

Sample 19
182
91
0
1b
29.9Kb

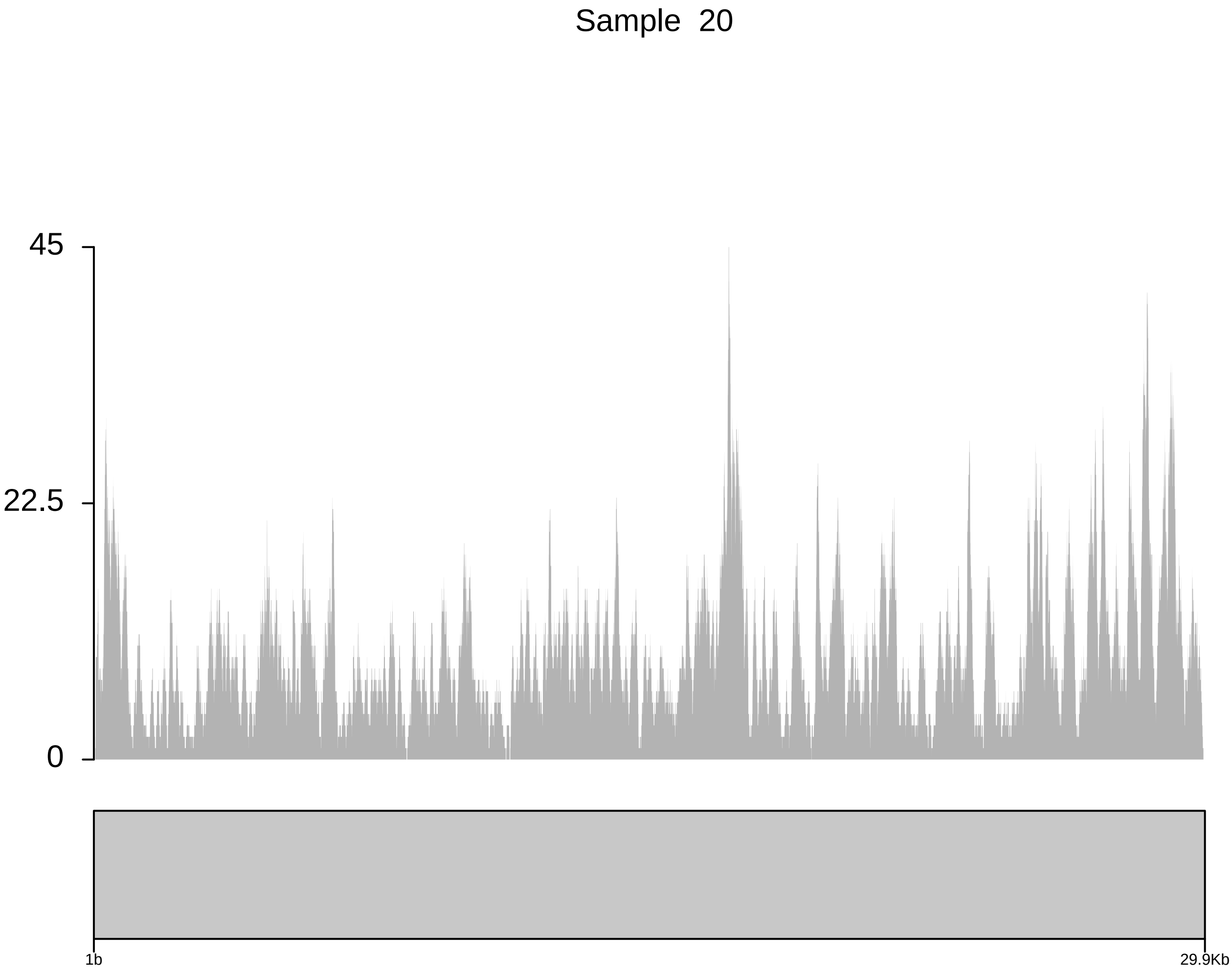
Sample 20
45
22.5
0
1b
29.9Kb

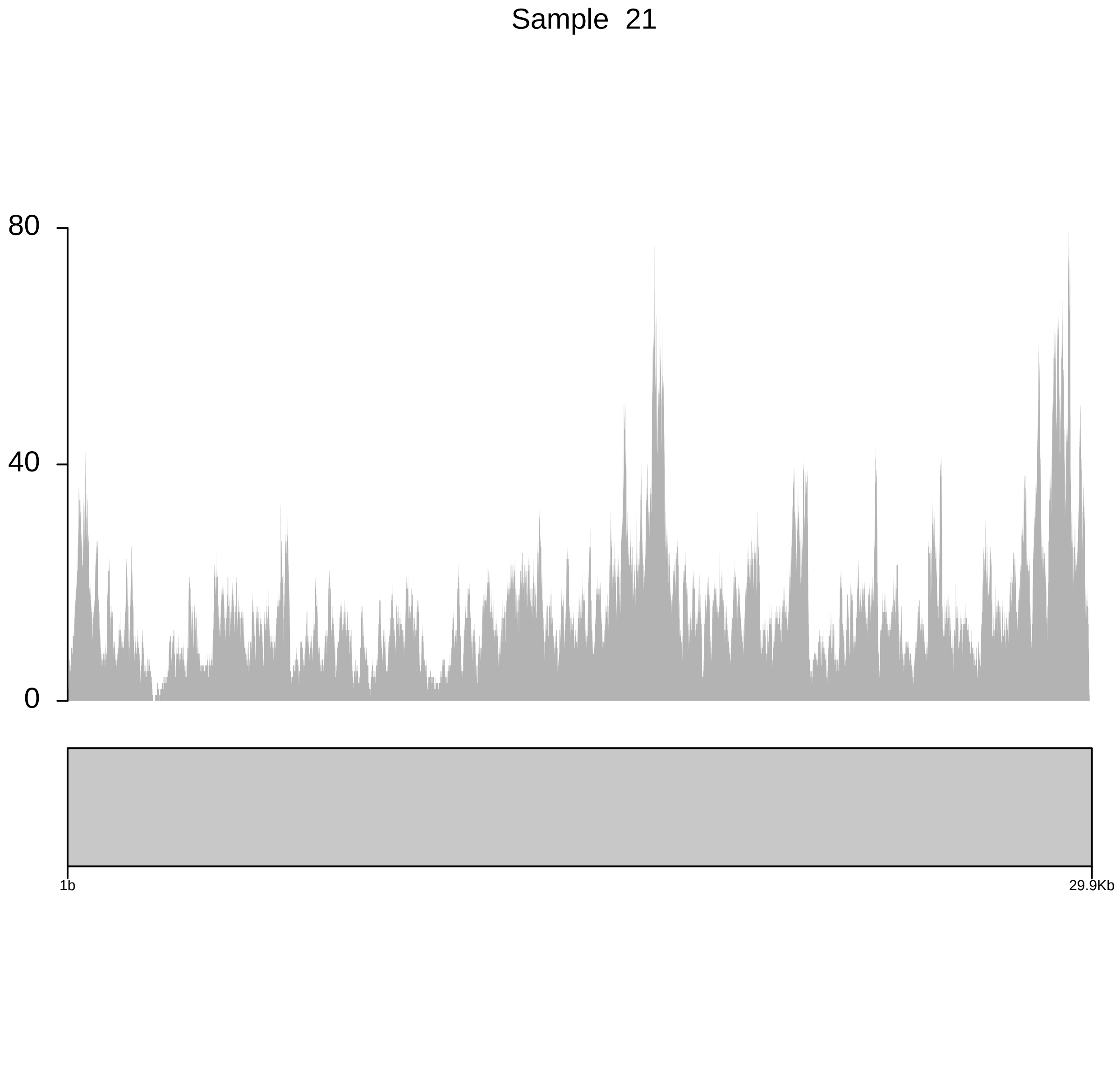
Sample 21
80
40
0
1b
29.9Kb
