## Supplemental File 2 for "Genomic Epidemiology of SARS-CoV-2 in Esteio, Rio Grande do Sul, Brazil"

**Supplemental File 2**. Detailed view of mutations across all samples. Nucleotide mutation positions are represented as rows. Each mutation has its own color. Cluster 1 and cluster 2 specific-mutations are colored in black and burnt yellow, respectively. Mutations related to Rio de Janeiro's new lineage are colored in red.

| Sample | 1 | 2 | 3 | 4 | 5 | 6 | 7 | 8 | 9 | 10 | 11 | 12 | 13 | 14 | 15 | 16 | 17 | 18 | 19 | 20 | 21 |
|---|---|---|---|---|---|---|---|---|---|---|---|---|---|---|---|---|---|---|---|---|---|
| Mutations | 241 | 241 | 147 | 241 | 147 | 28883 | 241 | 241 | 241 | 241 | 241 | 241 | 241 | 241 | 106 | 241 | 241 | 241 | 241 | 16207 | 241 |
| | 3037 | 3037 | 241 | 3037 | 241 | | 3037 | 2276 | 3037 | 3037 | 2276 | 3037 | 3037 | 3037 | 241 | 1218 | 2675 | 3037 | 3037 | 25245 | 3766 |
| | 8094 | 11211 | 850 | 14408 | 850 | | 11283 | 3037 | 12053 | 4706 | 3037 | 14408 | 13804 | 7005 | 23403 | 3037 | 3037 | 3766 | 7393 | 25494 | 4423 |
| | 14408 | 11758 | 3037 | 14805 | 3037 | | 13804 | 4644 | 14408 | 9167 | 8802 | 15720 | 14408 | 12053 | 28881 | 5339 | 7005 | 6218 | 12053 | 27205 | 12964 |
| | 23403 | 12484 | 12053 | 15654 | 6896 | | 14408 | 14408 | 23403 | 12053 | 10681 | 21191 | 19972 | 14408 | 28882 | 14408 | 12053 | 10667 | 14408 | 27299 | 14408 |
| | 27299 | 14408 | 14408 | 23403 | 12053 | | 23400 | 18252 | 25088 | 14408 | 14408 | 23403 | 20871 | 23403 | 28883 | 15654 | 14408 | 11824 | 14593 | 27501 | 23012 |
| | 28881 | 20679 | 20016 | 25718 | 14408 | | 27299 | 23403 | 25429 | 16912 | 18252 | 23455 | 23403 | 25088 | | 23403 | 23403 | 12053 | 23403 | 28881 | 23403 |
| | 28882 | 23403 | 23403 | 27299 | 20016 | | 28881 | 27299 | 25509 | 23403 | 18395 | 25062 | 24928 | 25429 | | 27299 | 25088 | 12964 | 25088 | 28882 | 28253 |
| | 28883 | 27299 | 25088 | 28881 | 23403 | | 28882 | 28093 | 27976 | 23593 | 22254 | 27299 | 28881 | 25509 | | 28881 | 25429 | 14408 | 25429 | 28883 | 28628 |
| | 29148 | 28881 | 25207 | 28882 | 25088 | | 28883 | 28881 | 28881 | 25088 | 23403 | 28881 | 28882 | 26019 | | 28882 | 25509 | 23012 | 25509 | 29148 | 28881 |
| | | 28882 | 25642 | 28883 | 25207 | | 29148 | 28882 | 28882 | 27828 | 27299 | 28882 | 28883 | 27976 | | 28883 | 26019 | 23403 | 27976 | | 28882 |
| | | 28883 | 28393 | 29148 | 25642 | | | 28883 | 28883 | 28881 | 28093 | 28883 | 29148 | 28881 | | 29148 | 27976 | 25088 | 28881 | | 28883 |
| | | 29148 | 28881 | | 28393 | | | 29148 | | 28882 | 28881 | 29148 | 29750 | 28882 | | | 28881 | 28253 | 28882 | | 28975 |
| | | 29224 | 28882 | | 28881 | | | | | 28883 | 28882 | | | 28883 | | | 28882 | 28628 | 28883 | | 29754 |
| | | | 28883 | | 28882 | | | | | 29744 | 28883 | | | | | | 28883 | 28881 | | | |
| | | | 29370 | | 28883 | | | | | | 29148 | | | | | | | 28882 | | | |

| | | | | | | | | | | | | | | | | | | | | | |
|---|---|---|---|---|---|---|---|---|---|---|---|---|---|---|---|---|---|---|---|---|---|
| | | | | | | | | | | | | | | | | | | 28883<br>28975<br>29754 | | | |
| Lineage | B.1.1.33 | B.1.1.33 | B.1.1.248 | B.1.1.33 | B.1.1.248 | B.1.1 | B.1.1.33 | B.1.1.33 | B.1.1.248 | B.1.1.248 | B.1.1.33 | B.1.1.33 | B.1.1.49 | B.1.1.248 | B.1.1 | B.1.1.33 | B.1.1.248 | B.1.1.248 | B.1.1.248 | B.1.1.33 | B.1.1 |
| # SNPs | 10 | 14 | 16 | 12 | 16 | 1 | 11 | 13 | 12 | 15 | 16 | 13 | 13 | 14 | 6 | 15 | 15 | 19 | 14 | 10 | 14 |
| Low depth SNPs (DP<10) | | | | | | 241 (9)<br>3037 (3)<br>10302 (1)<br>13804 (3)<br>15237 (4)<br>19255 (4)<br>23403 (3)<br>28881 (9)<br>28882 (9)<br>29148 (4) | | | | | | | 27299 (6) | | 2488 (1)<br>3037 (1)<br>3334 (2)<br>5835 (2)<br>6286 (5)<br>13560 (3)<br>20511 (2)<br>21396 (2)<br>26645 (4)<br>28093 (5)<br>29148 (5) | | | | | | 100 (7)<br>3037 (7)<br>10667 (2)<br>11074 (3)<br>11824 (7)<br>12053 (9)<br>25088 (9) |
| Lineage considering low depth SNPs | | | | | | B.1.1 | | | | | | | B.1.1.33 | | B.1.1 | | | | | | B.1.1.28 |
