## Supplemental File 3 for "Genomic Epidemiology of SARS-CoV-2 in Esteio, Rio Grande do Sul, Brazil"

| Author | n (strains) | strains | Publication title |
|---|---|---|---|
| Weijun Chen et al | 1 | Wuhan/WH01/2019 | Genomic characterisation and epidemiology of 2019 novel coronavirus: implications for virus origins and receptor binding |
| Cherng-Lih Perng et al | 10 | Taiwan/TSGH-37/2020,Taiwan/TSGH-34/2020,Taiwan/TSGH-35/2020,Taiwan/TSGH-31/2020,Taiwan/TSGH-27/2020,Taiwan/TSGH-36/2020,Taiwan/TSGH-23/2020,Taiwan/TSGH-24/2020,Taiwan/TSGH-05/2020,Taiwan/TSGH-03/2020 | - |
| Potdar V et al | 2 | India/MH-1-27/2020,India/MH-1-31/2020 | Full-genome sequences of the first two SARS-CoV-2 viruses from India |
| Jing Zhang et al | 2 | USA/WI1/2020,USA/CA-CDC-6/2020 | First 12 patients with coronavirus disease 2019 (COVID-19) in the United States |
| Caly L. et al | 5 | Australia/VIC14/2020,Australia/VIC02/2020,Australia/VIC03/2020,Australia/VIC876/2020,Australia/VIC156/2020 | Tracking the COVID-19 pandemic in Australia using genomics |
| Mak TM et al | 21 | Singapore/15/2020,Singapore/175/2020,Singapore/35/2020,Singapore/970/2020,Singapore/516/2020,Singapore/159/2020,Singapore/3Clin/2020,Singapore/910/2020,Singapore/684/2020,Singapore/687/2020,Singapore/795/2020,Singapore/607/2020,Singapore/903/2020,Singapore/864/2020,Singapore/865/2020,Singapore/880/2020,Singapore/879/2020,Singapore/891/2020,Singapore/889/2020,Singapore/295/2020,Singapore/376/2020 | - |
| Iglesias-Caballero et al | 9 | Spain/AN-ISCIII-2013382/2020,Spain/AN-ISCIII-2013675/2020,Andorra/202552/2020,Spain/CE-ISCIII-2016504/2020,Spain/CE-ISCIII-2016513/2020,Spain/MD-ISCIII-2014656/2020,Spain/CE-ISCIII-2016518/2020,Spain/CE-ISCIII-2016496/2020,Spain/CT-ISCIII-2013592/2020 | - |
| Amirtharaj Francis et al | 18 | UnitedArabEmirates/skmc-01319674/2020,UnitedArabEmirates/skmc-920168117/2020,UnitedArabEmirates/skmc-920569110/2020,UnitedArabEmirates/skmc-920568379/2020,UnitedArabEmirates/skmc-3819699/2020,UnitedArabEmirates/skmc-3819883/2020,UnitedArabEmirates/skmc-0381517/2020,UnitedArabEmirates/skmc-0532497/2020,UnitedArabEmirates/skmc-205223/2020,UnitedArabEmirates/skmc-3119719/2020,UnitedArabEmirates/skmc-3111681/2020,UnitedArabEmirates/skmc-3122376/2020,UnitedArabEmirates/skmc-3122201/2020,UnitedArabEmirates/skmc-3891025/2020,UnitedArabEmirates/skmc-3115511/2020,UnitedArabEmirates/skmc-3595813/2020,UnitedArabEmirates/skmc-3627476/2020,UnitedArabEmirates/skmc-227217/2020 | - |
| Jeong-Min Kim et al | 26 | SouthKorea/KCDC2002/2020,SouthKorea/KCDC07/2020,SouthKorea/KCDC2062/2020,SouthKorea/KCDC06/2020,SouthKorea/KCDC2291/2020,SouthKorea/KCDC2300/2020,SouthKorea/KCDC2001/2020,SouthKorea/KCDC12/2020,SouthKorea/KCDC2007/2020,SouthKorea/KCDC2004/2020,SouthKorea/KCDC2005/2020,SouthKorea/KCDC2196/2020,SouthKorea/KCDC2125/2020,SouthKorea/KCDC2194/2020,SouthKorea/KCDC2195/2020,SouthKorea/KCDC2534/2020,SouthKorea/KCDC2133/2020,SouthKorea/KCDC2134/2020,SouthKorea/KCDC2544/2020,SouthKorea/KCDC2704/2020,SouthKorea/KCDC2178/2020,SouthKorea/KCDC2186/2020,SouthKorea/KCDC2200/2020,SouthKorea/KCDC2558/2020,SouthKorea/KCDC2694/2020,SouthKorea/KCDC2695/2020 | - |

| | | | |
|---|---|---|---|
| Kuo-Chien Tsao et al | 2 | Taiwan/CGMH-CGU-02/2020,Taiwan/CGMH-CGU-01/2020 | SARS-CoV-2 genomic surveillance in Taiwan revealed novel ORF8-deletion mutant and clade possibly associated with infections in Middle East |
| Li et al | 1 | Cambodia/Kunming_kms-2/2020 | - |
| Mohd Noor Mat Isa et al | 8 | Malaysia/MKAK-CL-2020-5096/2020,Malaysia/MKAK-CL-2020-7554/2020,Malaysia/MKAK-CL-2020-5045/2020,Malaysia/MKAK-CL-2020-5049/2020,Malaysia/MKAK-CL-2020-5047/2020,Malaysia/MGI-MAEPS41/2020,Malaysia/MGI-MAEPS54/2020,Malaysia/MGI-MAEPS67/2020 | - |
| Monica Galiano et al | 8 | England/01/2020,England/02/2020,England/200641094/2020,England/200690300/2020,England/200690756/2020,England/200690245/2020,England/200690306/2020,England/20136085404/2020 | Preliminary analysis of SARS-CoV-2 importation & establishment of UK transmission lineages |
| Mee et al | 1 | England/2/2020 | - |
| Jeong-Min Kim et al | 1 | SouthKorea/KCDC03/2020 | Identification of Coronavirus Isolated from a Patient in Korea with COVID-19 |
| Le Quynh Mai et al | 3 | Vietnam/CM99/2020,Vietnam/38142/2020,Vietnam/39607/2020 | - |
| Ung Thi Hong Trang et al | 1 | Vietnam/VR03-38142/2020 | - |
| Sully Márquez et al | 18 | Ecuador/USFQ-556/2020,Ecuador/HEE-01/2020,Ecuador/USFQ-487/2020,Ecuador/USFQ-515/2020,Ecuador/USFQ-509/2020,Ecuador/USFQ-020/2020,Ecuador/USFQ-402/2020,Ecuador/USFQ-231/2020,Ecuador/USFQ-485/2020,Ecuador/USFQ-513/2020,Ecuador/USFQ-520/2020,Ecuador/USFQ-230/2020,Ecuador/USFQ-228/2020,Ecuador/USFQ-106/2020,Ecuador/USFQ-707/2020,Ecuador/USFQ-112/2020,Ecuador/USFQ-553/2020,Ecuador/USFQ-555/2020 | - |
| Danilo Franco et al | 20 | Panama/328723/2020,Panama/328709/2020,Panama/328719/2020,Panama/337660/2020,Panama/337467/2020,Panama/336897/2020,Panama/337442/2020,Panama/328941/2020,Panama/328927/2020,Panama/329198/2020,Panama/331837/2020,Panama/338677/2020,Panama/338657/2020,Panama/338674/2020,Panama/338687/2020,Panama/328677/2020,Panama/337640/2020,Panama/329117/2020,Panama/328844/2020,Panama/340837/2020 | Early transmission dynamics, spread, and genomic characterization of SARS-CoV-2 in Panama. |
| Valdinete Nascimento et al | 1 | Brazil/AM-02/2020 | COVID-19 in Latin America: Contrasting phylodynamic inference with epidemiological surveillance. |

| | | | |
|---|---|---|---|
| Nelson Gaburo Jr et al | 49 | Brazil/AM-L13-CD166/2020,Brazil/GO-L19-CD413/2020,Brazil/GO-L19-CD410/2020,Brazil/CE-L10-CD167/2020,Brazil/CE-L16-CD346/2020,Brazil/CE-L12-CD224/2020,Brazil/CE-L19-CD408/2020,Brazil/CE-L17-CD361/2020,Brazil/CE-L11-CD196/2020,Brazil/CE-L16-CD334/2020,Brazil/CE-L10-CD171/2020,Brazil/CE-L16-CD344/2020,Brazil/RN-L19-CD401/2020,Brazil/BA-L17-CD358/2020,Brazil/SP-L11-CD184-2/2020,Brazil/MT-L16-CD347/2020,Brazil/RJ-L10-CD154/2020,Brazil/SP-L12-CD202/2020,Brazil/SC-L16-CD314/2020,Brazil/RJ-L12-CD192/2020,Brazil/BA-L17-CD359/2020,Brazil/SP-L11-CD200/2020,Brazil/RS-L10-CD175/2020,Brazil/RJ-L10-CD158/2020,Brazil/RS-L11-CD180/2020,Brazil/MG-L11-CD183/2020,Brazil/MA-L19-CD403/2020,Brazil/MA-L19-CD407/2020,Brazil/SC-L17-CD322/2020,Brazil/MA-L17-CD363/2020,Brazil/RS-L19-CD390/2020,Brazil/RS-L17-CD370/2020,Brazil/SC-L17-CD372/2020,Brazil/RS-L19-CD412/2020,Brazil/MA-L19-CD406/2020,Brazil/MA-L19-CD405/2020,Brazil/SC-L19-CD399/2020,Brazil/PR-L11-CD179/2020,Brazil/CE-L16-CD332/2020,Brazil/SC-L16-CD350/2020,Brazil/PR-L21-CD448/2020,Brazil/PR-L16-CD333/2020,Brazil/PR-L17-CD367/2020,Brazil/PR-L17-CD366/2020,Brazil/PR-L17-CD368/2020,Brazil/PR-L19-CD389/2020,Brazil/CE-L21-CD447/2020,Brazil/PR-L17-CD365/2020,Brazil/PE-L20-CD432/2020 | Evolution and epidemic spread of SARS-CoV-2 in Brazil |
| Maria Dolores Ocete et al | 1 | Spain/VC-FISABIO-5/2020 | Phylodynamics of SARS-CoV-2 transmission in Spain |
| Abdaliyev Askar et al | 9 | Kazakhstan/7263/2020,Kazakhstan/16183/2020,Kazakhstan/7341/2020,Kazakhstan/17920/2020,Kazakhstan/22001/2020,Kazakhstan/18287/2020,Kazakhstan/18148/2020,Kazakhstan/38716/2020,Kazakhstan/33496/2020 | - |
| Carlos Cortes et al | 26 | EquatorialGuinea/253/2020,EquatorialGuinea/10071/2020,EquatorialGuinea/2892/2020,EquatorialGuinea/8971/2020,EquatorialGuinea/2274/2020,EquatorialGuinea/3350/2020,EquatorialGuinea/46787/2020,EquatorialGuinea/29422/2020,EquatorialGuinea/32447/2020,EquatorialGuinea/35052/2020,EquatorialGuinea/5117/2020,EquatorialGuinea/47665/2020,EquatorialGuinea/50236/2020,EquatorialGuinea/49849/2020,EquatorialGuinea/11514/2020,EquatorialGuinea/46215/2020,EquatorialGuinea/9134/2020,EquatorialGuinea/5508/2020,EquatorialGuinea/25536/2020,EquatorialGuinea/40288/2020,EquatorialGuinea/22908/2020,EquatorialGuinea/14513/2020,EquatorialGuinea/16990/2020,EquatorialGuinea/19424/2020,EquatorialGuinea/28974/2020,EquatorialGuinea/4361/2020 | - |
| Alexandr Shevtsov et al | 1 | Kazakhstan/16173/2020 | - |
| Pilailuk et al | 7 | Thailand/Bangkok_580/2020,Thailand/Samutprakarn_840/2020,Thailand/Nonthaburi_68/2020,Thailand/Phuket_247/2020,Thailand/NIH-1889/2020,Thailand/Nonthaburi_3472/2020,Thailand/Nonthaburi_2520/2020 | - |
| Rodpan et al | 4 | Thailand/SI202205-NT/2020,Thailand/SI201712-NT/2020,Thailand/SI200893-NT/2020,Thailand/SI200433-NT/2020 | - |
| Cao Minh Thắng et al | 1 | Vietnam/PIHCM-14/2020 | - |
| Ahmad Abou Tayoun et al | 7 | UnitedArabEmirates/L5630/2020,UnitedArabEmirates/L4280/2020,UnitedArabEmirates/L1014/2020,UnitedArabEmirates/L9766/2020,UnitedArabEmirates/L4184/2020,UnitedArabEmirates/L2409/2020,UnitedArabEmirates/L0881/2020 | Genomic surveillance and phylogenetic analysis reveal multiple introductions of SARS-CoV-2 into a global travel hub in the Middle East |
| Deborah A. Nickerson et al | 3 | USA/WA-S1317/2020,USA/WA-S2389/2020,USA/WA-S1649/2020 | - |
| Prado-Vivar et al | 2 | Ecuador/USFQ-165/2020,Ecuador/USFQ-167/2020 | - |
| Eden J-S et al | 2 | Australia/NSW01/2020,Australia/NSW03/2020 | Revealing COVID-19 transmission in Australia by SARS-CoV-2 genome sequencing and agent-based modeling |

| | | | |
|---|---|---|---|
| Ji-Rong Yang et al | 1 | Taiwan/3/2020 | SARS-CoV-2 genomic surveillance in Taiwan revealed novel ORF8-deletion mutant and clade possibly associated with infections in Middle East |
| Bert Vanmechelen et al | 3 | Belgium/GHB-03021/2020,Belgium/BA-02291/2020,Belgium/VDLH-030548-2/2020 | A phylodynamic workflow to rapidly gain insights into the dispersal history and dynamics of SARS-CoV-2 lineages |
| Tony Wawina-Bokalanga et al | 6 | Belgium/rega-0710751/2020,Belgium/DEJ-0403381/2020,Belgium/HI-0507421/2020,Belgium/Rega-0329233/2020,Belgium/rega-0513430/2020,Belgium/Rega-0331264/2020 | - |
| Hidalgo-Miranda A et al | 3 | Mexico/CMX-INMEGEN-12/2020,Mexico/CMX-INMEGEN-04/2020,Mexico/CMX-INMEGEN-07/2020 | - |
| Cecilia Salazar et al | 9 | Uruguay/UY-8M/2020,Uruguay/UY-272/2020,Uruguay/UY-242/2020,Uruguay/UY-162/2020,Uruguay/UY-957/2020,Uruguay/UY-652/2020,Uruguay/UY-667/2020,Uruguay/UY-650/2020,Uruguay/UY-651/2020 | - |
| Panzera et al | 1 | Uruguay/Mdeo-1/2020 | - |
| Irma López Martínez et al | 1 | Mexico/CMX-InDRE_03/2020 | Genomic analysis of early SARS-CoV-2 strains introduced in Mexico |
| Cecilia Salazar et al | 1 | Uruguay/UY-4/2020 | Multiple introductions, regional spread and local differentiation during the first week of COVID-19 epidemic in Montevideo, Uruguay |
| Maria Victoria Elizondo et al | 19 | Uruguay/UY-NYUMC869/2020,Uruguay/UY-NYUMC856/2020,Uruguay/UY-NYUMC860/2020,Uruguay/UY-NYUMC862/2020,Uruguay/UY-NYUMC863/2020,Uruguay/UY-NYUMC865/2020,Uruguay/UY-NYUMC933/2020,Uruguay/UY-NYUMC848/2020,Uruguay/UY-NYUMC871/2020,Uruguay/UY-NYUMC934/2020,Uruguay/UY-NYUMC872/2020,Uruguay/UY-NYUMC937/2020,Uruguay/UY-NYUMC936/2020,Uruguay/UY-NYUMC938/2020,Uruguay/UY-NYUMC939/2020,Uruguay/UY-NYUMC858/2020,Uruguay/UY-NYUMC931/2020,Uruguay/UY-NYUMC873/2020,Uruguay/UY-NYUMC857/2020 | - |
| Gisela Barrera Badillo et al | 1 | Mexico/QUE-InDRE-04/2020 | Genomic analysis of early SARS-CoV-2 strains introduced in Mexico |
| Carlos Padilla Rojas et al | 12 | Peru/LAL-INS-070/2020,Peru/LAL-INS-064/2020,Peru/LIM-INS-030/2020,Peru/LIM-INS-046/2020,Peru/LIM-INS-042/2020,Peru/LIM-INS-031/2020,Peru/LIM-INS-060/2020,Peru/ARE-INS-075/2020,Peru/LOR-INS-058/2020,Peru/ANC-INS-059/2020,Peru/ARE-INS-076/2020,Peru/LIM-INS-041/2020 | - |
| Rodriguez-Maldonado Abril et al | 1 | Mexico/QUE-InDRE-18/2020 | - |
| Darío García de Viedma et al | 6 | Spain/MD-IBV-99009630/2020,Spain/MD-IBV-99008612/2020,Spain/MD-IBV-99008616/2020,Spain/MD-IBV-99008619/2020,Spain/MD-IBV-99008615/2020,Spain/MD-IBV-99008700/2020 | - |

| | | | |
|---|---|---|---|
| Pablo Tsukayama et al | 42 | Peru/LIM-UPCH-0081/2020,Peru/LAM-UPCH-0006/2020,Peru/ARE-UPCH-0110/2020,Peru/LIM-UPCH-0056/2020,Peru/LIM-UPCH-0103/2020,Peru/LIM-UPCH-0025/2020,Peru/LIM-UPCH-0034/2020,Peru/AMZ-UPCH-0258/2020,Peru/AMZ-UPCH-0260/2020,Peru/LIM-UPCH-0182/2020,Peru/AMZ-UPCH-0277/2020,Peru/AMZ-UPCH-0261/2020,Peru/AMZ-UPCH-0263/2020,Peru/AMZ-UPCH-0270/2020,Peru/LIM-UPCH-0120/2020,Peru/LIM-UPCH-0165/2020,Peru/LIM-UPCH-0208/2020,Peru/LIM-UPCH-0184/2020,Peru/AMZ-UPCH-0268/2020,Peru/AMZ-UPCH-0255/2020,Peru/LIM-UPCH-0238/2020,Peru/LIM-UPCH-0242/2020,Peru/LIM-UPCH-0115/2020,Peru/LIM-UPCH-0176/2020,Peru/AMZ-UPCH-0259/2020,Peru/AMZ-UPCH-0264/2020,Peru/LIM-UPCH-0128/2020,Peru/LIM-UPCH-0147/2020,Peru/LIM-UPCH-0119/2020,Peru/AMZ-UPCH-0265/2020,Peru/LIM-UPCH-0194/2020,Peru/LIM-UPCH-0091/2020,Peru/LIM-UPCH-0137/2020,Peru/LIM-UPCH-0149/2020,Peru/LIM-UPCH-0123/2020,Peru/LIM-UPCH-0207/2020,Peru/AMZ-UPCH-0275/2020,Peru/AMZ-UPCH-0257/2020,Peru/LIM-UPCH-0243/2020,Peru/LIM-UPCH-0148/2020,Peru/LIM-UPCH-0145/2020,Peru/CAL-UPCH-0071/2020 | - |
| Laura Pérez-Lago et al | 3 | Spain/MD-IBV-004897/2020,Spain/MD-IBV-006379/2020,Spain/MD-IBV-004963/2020 | - |
| Kwabena O. Duedu et al | 4 | Ghana/UHAS-G541/2020,Ghana/UHAS-G441/2020,Ghana/UHAS-K674/2020,Ghana/UHAS-H009/2020 | - |
| Placide Mbala-Kingebeni et al | 8 | DRC/300/2020,DRC/253/2020,DRC/82/2020,DRC/1516/2020,DRC/2827/2020,DRC/2580/2020,DRC/248/2020,DRC/521/2020 | Phylogenetic analysis of SARS-CoV-2 in DRC |
| Goba et al | 7 | SierraLeone/KGH-G-8626/2020,SierraLeone/KGH-G-8668/2020,SierraLeone/KGH-G-8604/2020,SierraLeone/KGH-G-8548/2020,SierraLeone/KGH-G-8627/2020,SierraLeone/KGH-G-8607/2020,SierraLeone/KGH-G-8549/2020 | - |
| Fares et al | 2 | Tunisia/COV0010-12/2020,Tunisia/COV0425/2020 | First whole genome sequences and phylogenetic analysis of SARS-CoV-2 virus isolates during COVID-19 outbreak in Tunisia, North Africa. |
| Yadouleton et al | 9 | Benin/1950/2020,Benin/2012/2020,Benin/461/2020,Benin/843/2020,Benin/1409/2020,Benin/1408/2020,Benin/501/2020,Benin/1022/2020,Benin/197/2020 | Diagnostics and spread of SARS-CoV-2 in Western Africa: An observational laboratory-based study from Benin |
| Ngoi et al | 8 | Ghana/84162_S45/2020,Ghana/36523_S23/2020,Ghana/35183_S24/2020,Ghana/2666_S32/2020,Ghana/2852_S30/2020,Ghana/83637_S29/2020,Ghana/81219_S34/2020,Ghana/35019_S18/2020 | - |
| Handrick et al | 1 | Tunisia/MHT_2/2020 | - |
| Hesham Elghazaly et al | 7 | Egypt/MASRI-009/2020,Egypt/MASRI-014/2020,Egypt/MASRI-007/2020,Egypt/MASRI-005/2020,Egypt/MASRI-008/2020,Egypt/MASRI-012/2020,Egypt/MASRI-013/2020 | - |

| | | | |
|---|---|---|---|
| Bas Oude Munnink et al | 74 | Bahrein/BAH-24/2020,Bahrein/BAH-12/2020,Netherlands/FL-EMC-28/2020,Netherlands/GE-EMC-215/2020,Netherlands/ZH-EMC-394/2020,Netherlands/ZH-EMC-354/2020,Bahrein/BAH-02/2020,Bahrein/BAH-05/2020,Bahrein/BAH-08/2020,Bahrein/BAH-26/2020,Bahrein/BAH-21/2020,Bahrein/BAH-19/2020,Bahrein/BAH-09/2020,Netherlands/ZuidHolland_60/2020,Netherlands/NA_551/2020,Netherlands/NB-EMC-241/2020,Suriname/SR-32/2020,Suriname/SR-59/2020,Suriname/SR-47/2020,Suriname/SR-42/2020,Suriname/SR-70/2020,Suriname/SR-05/2020,Suriname/SR-69/2020,Suriname/SR-37/2020,Suriname/SR-41/2020,Suriname/SR-48/2020,Suriname/SR-60/2020,Suriname/SR-10/2020,Suriname/SR-11/2020,Suriname/SR-51/2020,Suriname/SR-39/2020,Suriname/SR-40/2020,Suriname/SR-15/2020,Suriname/SR-36/2020,Netherlands/UT-EMC-180/2020,Netherlands/ZH-EMC-1042/2020,Netherlands/GE-EMC-183/2020,Netherlands/ZH-EMC-663/2020,Netherlands/ZH-EMC-303/2020,Suriname/SR-63/2020,Suriname/SR-46/2020,Suriname/SR-58/2020,Suriname/SR-56/2020,Suriname/SR-62/2020,Netherlands/ZH-EMC-868/2020,Netherlands/NB-EMC-318/2020,Netherlands/NH-EMC-41/2020,Netherlands/ZuidHolland_65/2020,Netherlands/NA_542/2020,Netherlands/ZH-EMC-632/2020,Netherlands/ZH-EMC-891/2020,Netherlands/un-EMC-780/2020,Netherlands/GE-EMC-432/2020,Netherlands/GE-EMC-413/2020,Bahrein/BAH-23/2020,Netherlands/ZH-EMC-894/2020,Netherlands/ZH-EMC-360/2020,Netherlands/NA_354/2020,Netherlands/OV-EMC-55/2020,Netherlands/NA_440/2020,Netherlands/ZH-EMC-1056/2020,Netherlands/ZH-EMC-1090/2020,Netherlands/UT-EMC-57/2020,Netherlands/OV-EMC-59/2020,Netherlands/OV-EMC-60/2020,Netherlands/ZH-EMC-812/2020,Netherlands/NB-EMC-439/2020,Netherlands/ZE-EMC-85/2020,Netherlands/ZH-EMC-804/2020,Netherlands/ZH-EMC-726/2020,Netherlands/GE-EMC-417/2020,Netherlands/ZH-EMC-261/2020,Netherlands/un-EMC-757/2020,Netherlands/ZH-EMC-598/2020 | - |
| Fadwa Alofi et al | 4 | SaudiArabia/KAUST-JEDDAH467/2020,SaudiArabia/KAUST-JEDDAH468/2020,SaudiArabia/KAUST-JEDDAH469/2020,SaudiArabia/KAUST-JEDDAH466/2020 | - |
| Haruka Abe et al | 4 | Gabon/ITM-K011/2020,Gabon/ITM-K027/2020,Gabon/ITM-K028/2020,Gabon/ITM-K023/2020 | - |
| Chantal Akoua-Koffi et al | 22 | CotedIvoire/BKE0016/2020,CotedIvoire/BKE0011/2020,CotedIvoire/BKE0761/2020,CotedIvoire/BKE0862/2020,CotedIvoire/BKE0965/2020,CotedIvoire/BKE0820/2020,CotedIvoire/BKE0906/2020,CotedIvoire/BKE0172/2020,CotedIvoire/BKE0613/2020,CotedIvoire/BKE0317/2020,CotedIvoire/BKE0356/2020,CotedIvoire/BKE0042/2020,CotedIvoire/BKE0430/2020,CotedIvoire/BKE0405/2020,CotedIvoire/BKE0753/2020,CotedIvoire/BKE0178/2020,CotedIvoire/BKE0916/2020,CotedIvoire/BKE0880/2020,CotedIvoire/BKE0018/2020,CotedIvoire/C01-2459/2020,CotedIvoire/BKE0994/2020,CotedIvoire/BKE0992/2020 | - |
| Issa Abu-Dayyeh et al | 16 | Jordan/IR-ALSR-2578/2020,Jordan/SR-0336/2020,Jordan/SR-037/2020,Jordan/SR-051/2020,Jordan/SR-056/2020,Jordan/SR-0333/2020,Jordan/SR-0338/2020,Jordan/AM-ALSR-2586/2020,Jordan/AM-ALSR-2579/2020,Jordan/AM-ALSR-2585/2020,Jordan/MA-ALSR-2581/2020,Jordan/MA-ALSR-2583/2020,Jordan/SR-034/2020,Jordan/SR-055/2020,Jordan/SR-041/2020,Jordan/SR-052/2020 | - |
| Kouriba et al | 5 | Mali/M002698/2020,Mali/M002616/2020,Mali/M002704/2020,Mali/M002826/2020,Mali/M002673/2020 | - |

| | | | |
|---|---|---|---|
| Abdoul-Salam Ouedraogo et al | 28 | BurkinaFaso/2-009/2020,BurkinaFaso/2-012/2020,BurkinaFaso/3535/2020,BurkinaFaso/4307/2020,BurkinaFaso/3953/2020,BurkinaFaso/4215/2020,BurkinaFaso/4312/2020,BurkinaFaso/583/2020,BurkinaFaso/1941/2020,BurkinaFaso/609/2020,BurkinaFaso/588/2020,BurkinaFaso/1447/2020,BurkinaFaso/590/2020,BurkinaFaso/2173/2020,BurkinaFaso/2-043-1764/2020,BurkinaFaso/2-060-1985/2020,BurkinaFaso/2679/2020,BurkinaFaso/1174/2020,BurkinaFaso/2-032-1197/2020,BurkinaFaso/3567/2020,BurkinaFaso/3952/2020,BurkinaFaso/2163/2020,BurkinaFaso/2453/2020,BurkinaFaso/2481/2020,BurkinaFaso/608/2020,BurkinaFaso/1124/2020,BurkinaFaso/2-035-1347/2020,BurkinaFaso/2-042-1650/2020 | - |
| Rong Liu et al | 10 | UnitedArabEmirates/1003/2020,UnitedArabEmirates/0064/2020,UnitedArabEmirates/1046/2020,UnitedArabEmirates/0989/2020,UnitedArabEmirates/0919/2020,UnitedArabEmirates/0945/2020,UnitedArabEmirates/0821/2020,UnitedArabEmirates/0120/2020,UnitedArabEmirates/0198/2020,UnitedArabEmirates/0195/2020 | - |
| Victor M Corman et al | 45 | Germany/DE-SH-ChVir8595/2020,Germany/DE-NW-ChVir1683/2020,Moldova/ChVir7770/2020,BosniaandHerzegovina/ChVir7402/2020,Montenegro/ChVir-1265622002/2020,Montenegro/ChVir-1268662002/2020,BosniaandHerzegovina/ChVir7353/2020,BosniaandHerzegovina/ChVir7350/2020,BosniaandHerzegovina/ChVir7348/2020,BosniaandHerzegovina/ChVir7362/2020,Moldova/ChVir7771/2020,BosniaandHerzegovina/ChVir7382/2020,BosniaandHerzegovina/ChVir7387/2020,BosniaandHerzegovina/ChVir7396/2020,Moldova/ChVir7768/2020,CzechRepublic/ChVir1630/2020,Germany/DE-BE-ChVir1716/2020,Moldova/ChVir7776/2020,Moldova/ChVir7777/2020,BosniaandHerzegovina/ChVir7367/2020,Germany/DE-SH-ChVir8328/2020,Germany/DE-SH-ChVir9939/2020,Montenegro/ChVir-1796572002/2020,Montenegro/ChVir-1807132002/2020,Montenegro/ChVir-1794332002/2020,Montenegro/ChVir-1793232002/2020,CzechRepublic/ChVir1912/2020,Germany/DE-BE-ChVir8329/2020,Germany/DE-SH-ChVir9635/2020,Germany/DE-SH-ChVir9634/2020,Germany/DE-SH-ChVir20012/2020,Germany/DE-SH-ChVir20784/2020,Montenegro/ChVir-1267392002/2020,Germany/DE-SH-ChVir20797/2020,Germany/DE-SH-ChVir9984/2020,Germany/DE-SH-ChVir9953/2020,Germany/DE-SH-ChVir9944/2020,Germany/DE-SH-ChVir20807/2020,Germany/DE-SH-ChVir8192/2020,Germany/DE-SH-ChVir8194/2020,Germany/DE-SH-ChVir8179/2020,Montenegro/ChVir-1800902002/2020,Germany/DE-SH-ChVir8311/2020,Germany/DE-BE-ChVir9006/2020,Germany/DE-BE-ChVir7622/2020 | - |
| Massab Umair et al | 4 | Pakistan/NIH-44905/2020,Pakistan/NIH-HAS001/2020,Pakistan/NIH-45090/2020,Pakistan/NIH-45143/2020 | - |
| Dan Lule Bugembe et al | 10 | Uganda/UG017/2020,Uganda/UG001/2020,Uganda/UG002/2020,Uganda/UG020/2020,Uganda/UG016/2020,Uganda/UG019/2020,Uganda/UG015/2020,Uganda/UG014/2020,Uganda/UG009/2020,Uganda/UG003/2020 | Main Routes of Entry and Genomic Diversity of SARS-CoV-2, Uganda |

| | | | |
|---|---|---|---|
| Oluniyi P.E. et al et al | 33 | Nigeria/ED04-CV158/2020,Nigeria/ED02-CV156/2020,Nigeria/OS-CV287/2020,Nigeria/OS-CV302/2020,Nigeria/OS-CV301/2020,Nigeria/LA-CV253/2020,Nigeria/LA-CV370/2020,Nigeria/ED18-CV161/2020,Nigeria/LA-CV389/2020,Nigeria/ED25-CV163/2020,Nigeria/ED82-CV208/2020,Nigeria/ED48-CV193/2020,Nigeria/OS-CV303/2020,Nigeria/EK291-CV115/2020,Nigeria/AK-CV317/2020,Nigeria/OS-CV296/2020,Nigeria/OS-CV306/2020,Nigeria/OS-CV299/2020,Nigeria/BO-CV333/2020,Nigeria/OS-CV297/2020,Nigeria/FC-CV250/2020,Nigeria/OY636-CV192/2020,Nigeria/OS-CV294/2020,Nigeria/OS-CV292/2020,Nigeria/OS-CV307/2020,Nigeria/OS-CV304/2020,Nigeria/LA-CV379/2020,Nigeria/OY627-CV184/2020,Nigeria/LA-CV261/2020,Nigeria/LA-CV371/2020,Nigeria/OG-CV260/2020,Nigeria/FC-CV356/2020,Nigeria/OS-CV293/2020 | - |
| PathWest Laboratory Medicine WA Microbial Surveillance Unit et al | 3 | Australia/WA201/2020,Australia/WA203/2020,Australia/WA405/2020 | - |
| Tze Minn Mak et al | 31 | Singapore/1390/2020,Singapore/1013/2020,Singapore/472cul/2020,Singapore/1389/2020,PapuaNewGuinea/8/2020,PapuaNewGuinea/3/2020,PapuaNewGuinea/5/2020,PapuaNewGuinea/4/2020,PapuaNewGuinea/2/2020,PapuaNewGuinea/1/2020,PapuaNewGuinea/10/2020,PapuaNewGuinea/13/2020,PapuaNewGuinea/9/2020,PapuaNewGuinea/7/2020,PapuaNewGuinea/11/2020,PapuaNewGuinea/6/2020,PapuaNewGuinea/12/2020,Singapore/1444/2020,Singapore/1428/2020,Singapore/1462/2020,Singapore/1438/2020,Singapore/1451/2020,Singapore/1391/2020,Singapore/1101/2020,Singapore/1132/2020,Singapore/1108/2020,Singapore/1456/2020,Singapore/1083/2020,Singapore/1090/2020,Singapore/1440/2020,Singapore/1447/2020 | - |
| Placide Mbala-Kingebeni et al | 11 | DRC/04187/2020,DRC/04186/2020,DRC/16133/2020,DRC/16145/2020,DRC/16143/2020,DRC/16191/2020,DRC/6392/2020,DRC/10506/2020,DRC/10502/2020,DRC/10532/2020,DRC/7488/2020 | - |
| Augustina Sylverken et al | 2 | Ghana/TTH10/2020,Ghana/TTH11/2020 | - |
| Tsuyoshi Sekizuka et al | 7 | Japan/PG-0016/2020,Japan/PG-0015/2020,Japan/PG-0425/2020,Japan/PG-0541/2020,Japan/PG-0547/2020,Japan/PG-0546/2020,Japan/PG-0552/2020 | A genome epidemiological study of SARS-CoV-2 introduction into Japan |
| Yoong Min CHONG et al | 5 | Malaysia/6359/2020,Malaysia/0784/2020,Malaysia/0956/2020,Malaysia/7618/2020,Malaysia/8454/2020 | - |
| Rockett R et al | 3 | Australia/NSW02/2020,Australia/NSW10/2020,Australia/NSW08/2020 | Revealing COVID-19 transmission in Australia by SARS-CoV-2 genome sequencing and agent-based modeling |
| Shari Tyson et al | 1 | Canada/ON_VIDO-01/2020 | - |
| Amrit S. Boese et al | 1 | Canada/ON_ON-VIDO-01-2/2020 | - |
| Andrés E Castillo et al | 12 | Chile/LI-ISPCH-2/2020,Chile/RM-ISPCH-71/2020,Chile/RM-ISPCH-73/2020,Chile/RM-ISPCH-66/2020,Chile/RM-ISPCH-76/2020,Chile/RM-ISPCH-52/2020,Chile/RM-ISPCH-56/2020,Chile/RM-ISPCH-74/2020,Chile/RM-ISPCH-23/2020,Chile/RM-ISPCH-40/2020,Chile/RM-ISPCH-42/2020,Chile/RM-ISPCH-38/2020 | - |
| Ranjit Sah et al | 1 | Nepal/61/2020 | Insights into The Codon Usage Bias of 13 Severe Acute Respiratory Syndrome Coronavirus 2 (SARS-CoV-2) Isolates from Different Geo-locations |
| Shiou-Hwei Yeh et al | 1 | Taiwan/NTU02/2020 | Insights into The Codon Usage Bias of 13 Severe Acute Respiratory Syndrome Coronavirus 2 (SARS-CoV-2) Isolates from Different Geo-locations |
| Suppiah.J et al | 5 | Malaysia/IMR_WC627/2020,Malaysia/IMR_WC085/2020,Malaysia/IMR_WC1097/2020,Malaysia/IMR_WC1098/2020,Malaysia/IMR_WC1170/2020 | - |

| | | | |
|---|---|---|---|
| Nurdyana Abdul Rahman et al | 5 | Singapore/1nan/2020,Singapore/316/2020,Singapore/538/2020,Singapore/318/2020,Singapore/1060/2020 | - |
| Danielle E Anderson et al | 2 | Singapore/6/2020,Singapore/2/2020 | Discovery of a 382-nt deletion during the early evolution of SARS-CoV-2 |
| Anna Uehara et al | 1 | USA/CA2/2020 | First 12 patients with coronavirus disease 2019 (COVID-19) in the United States |
| Oskar Karlsson Lindsjo et al | 11 | Sweden/20-02114/2020,Sweden/20-04736/2020,Sweden/20-04851/2020,Sweden/20-04850/2020,Sweden/20-04852/2020,Sweden/20-50185/2020,Sweden/20-51988/2020,Sweden/20-51277/2020,Sweden/20-14377/2020,Sweden/20-14518/2020,Sweden/20-08909/2020 | - |
| Mélanie Albert et al | 2 | France/IDF-0372/2020,France/IDF-2792/2020 | Introductions and early spread of SARS-CoV-2 in France |
| Mélanie Albert et al | 2 | France/IDF-0372-isl/2020,France/IDF-0626/2020 | Whole genome and phylogenetic analysis of two SARS-CoV-2 strains isolated in Italy in January and February 2020: additional clues on multiple introductions and further circulation in Europe |
| Olivier Terrier et al | 2 | France/IDF-0386-islP1/2020,France/IDF-0386-islP3/2020 | Whole genome and phylogenetic analysis of two SARS-CoV-2 strains isolated in Italy in January and February 2020: additional clues on multiple introductions and further circulation in Europe |
| Maria R. Capobianchi et al | 1 | Italy/LAZ-INMI1-isl/2020 | Whole genome and phylogenetic analysis of two SARS-CoV-2 strains isolated in Italy in January and February 2020: additional clues on multiple introductions and further circulation in Europe |
| Kenneth Siu-Sing LEUNG et al | 5 | HongKong/HKPU17_2201/2020,HongKong/HKPU28_3001/2020,HongKong/HKPU60-0802/2020,HongKong/HKPU57-0702/2020,HongKong/HKPU66-2501/2020 | A territory-wide study of early COVID-19 outbreak in Hong Kong community: A clinical, epidemiological and phylogenomic investigation |
| Samira Al-Maruqi et al | 7 | Oman/RESP-20-4028/2020,Oman/RESP-20-3700/2020,Oman/RESP-20-6560/2020,Oman/RESP-20-837/2020,Oman/RESP-20-797/2020,Oman/RESP-20-3080/2020,Oman/810962/2020 | - |
| Seemann et al | 4 | Australia/VIC1820/2020,Australia/VIC10678/2020,Australia/VIC12604/2020,Australia/VIC9319/2020 | - |
| Caly L. et al | 5 | Australia/VIC2387/2020,Australia/VIC3049/2020,Australia/VIC3521/2020,Australia/VIC16197/2020,Australia/VIC11900/2020 | - |
| Tsuyoshi Sekizuka et al | 2 | Japan/AI-I-004/2020,Japan/NA-20-05-1/2020 | - |
| Ivars Silamiķelis et al | 39 | Latvia/040/2020,Latvia/044/2020,Latvia/116/2020,Latvia/010/2020,Latvia/115/2020,Latvia/019/2020,Latvia/156/2020,Latvia/112/2020,Latvia/132/2020,Latvia/193/2020,Latvia/190/2020,Latvia/053/2020,Latvia/084/2020,Latvia/071/2020,Latvia/083/2020,Latvia/085/2020,Latvia/038/2020,Latvia/020/2020,Latvia/037/2020,Latvia/030/2020,Latvia/054/2020,Latvia/042/2020,Latvia/011/2020,Latvia/149/2020,Latvia/147/2020,Latvia/171/2020,Latvia/172/2020,Latvia/195/2020,Latvia/034/2020,Latvia/028/2020,Latvia/035/2020,Latvia/059/2020,Latvia/067/2020,Latvia/058/2020,Latvia/065/2020,Latvia/068/2020,Latvia/196/2020,Latvia/179/2020,Latvia/163/2020 | - |
| Shakeel et al | 1 | Pakistan/KHI1/2020 | - |
| Ying Tao et al | 2 | USA/CA-CDC-5/2020,USA/IL1/2020 | First 12 patients with coronavirus disease 2019 (COVID-19) in the United States |

| | | | |
|---|---|---|---|
| Lukas Zemaitis et al | 20 | Lithuania/KU-LUHS-Eilnr10/2020,Lithuania/MR-LUHS-Eilnr19/2020,Lithuania/MR-LUHS-Eilnr31/2020,Lithuania/MR-LUHS-Eilnr65/2020,Lithuania/MR-LUHS-Eilnr67/2020,Lithuania/MR-LUHS-Eilnr462/2020,Lithuania/MR-LUHS-Eilnr422/2020,Lithuania/MR-LUHS-Eilnr427/2020,Lithuania/MR-LUHS-Eilnr203/2020,Lithuania/MR-LUHS-Eilnr201/2020,Lithuania/MR-LUHS-Eilnr442/2020,Lithuania/MR-LUHS-Eilnr228/2020,Lithuania/MR-LUHS-Eilnr227/2020,Lithuania/MR-LUHS-Eilnr249/2020,Lithuania/MR-LUHS-Eilnr247/2020,Lithuania/MR-LUHS-Eilnr414/2020,Lithuania/MR-LUHS-Eilnr230/2020,Lithuania/MR-LUHS-Eilnr172/2020,Lithuania/MR-LUHS-Eilnr173/2020,Lithuania/MR-LUHS-Eilnr189/2020 | - |
| Rima R Prasetya et al | 1 | Indonesia/JI-ITD-3101NT/2020 | - |
| Ji-Rong Yang et al | 2 | Taiwan/5/2020,Taiwan/8/2020 | - |
| Alan K.L. Tsang et al | 6 | HongKong/VB20017970-2/2020,HongKong/VM20106598/2020,HongKong/VM20107468/2020,HongKong/VB20175856/2020,HongKong/VM20108307/2020,HongKong/VM20008964-2/2020 | - |
| To et al | 3 | HongKong/HKU-902b/2020,HongKong/HKU-902a/2020,HongKong/HKU-911a/2020 | - |
| Zhang et al | 1 | Wuhan/Hu-1/2019 | A new coronavirus associated with human respiratory disease in China |
| Fan et al | 2 | Wuhan/IME-WH04/2020,Wuhan/IME-WH02/2020 | - |
| Cao et al | 1 | Vietnam/19-01S/2020 | Insights into The Codon Usage Bias of 13 Severe Acute Respiratory Syndrome Coronavirus 2 (SARS-CoV-2) Isolates from Different Geo-locations |
| Nguyen et al | 1 | Vietnam/19-02S/2020 | - |
| Mak et al | 1 | Singapore/1/2020 | Whole genome and phylogenetic analysis of two SARS-CoV-2 strains isolated in Italy in January and February 2020: additional clues on multiple introductions and further circulation in Europe |
| Erik A Karlsson et al | 1 | Cambodia/0012/2020 | Whole genome and phylogenetic analysis of two SARS-CoV-2 strains isolated in Italy in January and February 2020: additional clues on multiple introductions and further circulation in Europe |
| Peng Zhou et al | 1 | Wuhan/WIV06/2019 | - |
| Lili Ren et al | 1 | Wuhan/IPBCAMS-WH-04/2019 | Genomic diversity of SARS-CoV-2 in Coronavirus Disease 2019 patients |
| Kelvin K.W. To et al | 13 | HongKong/HKU-200723-060/2020,HongKong/HKU-200723-051/2020,HongKong/HKU-200723-053/2020,HongKong/HKU-200723-105/2020,HongKong/HKU-200723-003/2020,HongKong/HKU-200723-106/2020,HongKong/HKU-200723-073/2020,HongKong/HKU-200723-109/2020,HongKong/HKU-200723-078/2020,HongKong/HKU-200723-056/2020,HongKong/HKU-200723-098/2020,HongKong/HKU-200723-092/2020,HongKong/HKU-200723-071/2020 | - |
| Pilailuk et al | 2 | Thailand/61/2020,Thailand/74/2020 | Early transmission patterns of coronavirus disease 2019 (COVID-19) in travellers from Wuhan to Thailand, January 2020 |
| María Rodríguez et al | 3 | Spain/MD-HLP-113/2020,Spain/MD-HLP-95/2020,Spain/MD-HLP-77/2020 | - |
| Krista Queen et al | 3 | USA/CA-CDC-8/2020,USA/CA-CDC-9/2020,Jamaica/JM-CDC-0078/2020 | - |

| | | | |
|---|---|---|---|
| Adam Meijer et al | 25 | Curacao/CW-RIVM-10308/2020,Curacao/CW-RIVM-10309/2020,Curacao/CW-RIVM-10093/2020,Curacao/CW-RIVM-10092/2020,Curacao/CW-RIVM-10094/2020,Curacao/CW-RIVM-10100/2020,Aruba/AW-RIVM-10101/2020,Aruba/AW-RIVM-10122/2020,Aruba/AW-RIVM-10115/2020,Aruba/AW-RIVM-10113/2020,Aruba/AW-RIVM-10114/2020,Aruba/AW-RIVM-10390/2020,Aruba/AW-RIVM-10388/2020,Aruba/AW-RIVM-10389/2020,Aruba/AW-RIVM-10392/2020,Curacao/CW-RIVM-10364/2020,Curacao/CW-RIVM-10365/2020,Curacao/CW-RIVM-10366/2020,Aruba/AW-RIVM-10326/2020,Aruba/AW-RIVM-10391/2020,Netherlands/NH-RIVM-10073/2020,Netherlands/NoordHolland_10001/2020,Netherlands/NH-RIVM-20344/2020,Netherlands/ZuidHolland_10004/2020,Netherlands/GE-RIVM-20216/2020 | - |
| Ortwin Adams et al | 4 | Germany/NW-HHU-02-1/2020,Germany/NW-HHU-03/2020,Germany/NW-HHU-05/2020,Germany/NW-HHU-06/2020 | Genetic structure of SARS-CoV-2 in Western Germany reflects clonal superspreading and multiple independent introduction events |
| Kathrine Stene-Johansen et al | 44 | Norway/2498/2020,Norway/1986/2020,Norway/2087/2020,Norway/1494/2020,Norway/1455/2020,Norway/1443/2020,Norway/1493/2020,Norway/1694/2020,Norway/1379/2020,Norway/2534/2020,Norway/1955/2020,Norway/2356-2/2020,Norway/2200/2020,Norway/2812/2020,Norway/5327/2020,Norway/3069/2020,Norway/3365/2020,Norway/2850/2020,Norway/2846/2020,Norway/2856/2020,Norway/5487/2020,Norway/5334/2020,Norway/5171/2020,Norway/5643/2020,Norway/3756/2020,Norway/2958/2020,Norway/3620/2020,Norway/2781/2020,Norway/2893/2020,Norway/4059/2020,Norway/3376/2020,Norway/3378/2020,Norway/3272/2020,Norway/3278/2020,Norway/4125/2020,Norway/4040/2020,Norway/3833/2020,Norway/3602/2020,Norway/5121/2020,Norway/4779/2020,Norway/4817/2020,Norway/2855/2020,Norway/2854/2020,Norway/2853/2020 | - |
| Michael Carr et al | 36 | Ireland/D-NVRL-AIIDV0328v1/2020,Ireland/D-NVRL-AIIDV0328v2/2020,Ireland/CO-NVRL-20134/2020,Ireland/D-NVRL-71IRL15701/2020,Ireland/LK-NVRL-19935/2020,Ireland/CO-NVRL-20IRL79223/2020,Ireland/D-NVRL-20IRL87996/2020,Ireland/D-NVRL-AIIDV0091v1/2020,Ireland/LS-NVRL-75IRL70478/2020,Ireland/CO-NVRL-75IRL73676/2020,Ireland/CO-NVRL-75IRL73629/2020,Ireland/D-NVRL-AIIDM0047v1/2020,Ireland/G-NVRL-75IRL85497/2020,Ireland/D-NVRL-72IRL12139/2020,Ireland/WW-NVRL-24052/2020,Ireland/CN-NVRL-71IRL96106/2020,Ireland/D-NVRL-AIIDM0017v1/2020,Ireland/D-NVRL-20IRL27755/2020,Ireland/LH-NVRL-20IRL26172/2020,Ireland/D-NVRL-AIIDV0019v1/2020,Ireland/MH-NVRL-72IRL14688/2020,Ireland/D-NVRL-72IRL38771/2020,Ireland/D-NVRL-72IRL56952/2020,Ireland/MO-NVRL-73IRL31845/2020,Ireland/MH-NVRL-73IRL22943/2020,Ireland/LH-NVRL-73IRL72600/2020,Ireland/LH-NVRL-73IRL72673/2020,Ireland/CN-NVRL-73IRL72730/2020,Ireland/WW-NVRL-AIIDV0073v2/2020,Ireland/WW-NVRL-AIIDV0073v1/2020,Ireland/D-NVRL-20IRL36562/2020,Ireland/D-NVRL-71IRL07158/2020,Ireland/D-NVRL-20IRL43282/2020,Ireland/D-NVRL-71IRL21349/2020,Ireland/LH-NVRL-20IRL52333/2020,Ireland/WW-NVRL-AIIDV0073v4/2020 | - |
| Alexandra Popa et al | 12 | Austria/CeMM0002/2020,Austria/CeMM0120/2020,Austria/CeMM0639/2020,Austria/CeMM0553/2020,Austria/CeMM0255/2020,Austria/CeMM0386/2020,Austria/CeMM0352/2020,Austria/CeMM0003/2020,Austria/CeMM0143/2020,Austria/CeMM0146/2020,Austria/CeMM0045/2020,Austria/CeMM0530/2020 | Mutational dynamics and transmission properties of SARS-CoV-2 superspreading events in Austria |
| Ivan Ivanov et al | 13 | Bulgaria/15/2020,Bulgaria/23/2020,Bulgaria/41/2020,Bulgaria/40/2020,Bulgaria/36/2020,Bulgaria/17/2020,Bulgaria/06/2020,Bulgaria/37/2020,Bulgaria/10/2020,Bulgaria/34/2020,Bulgaria/27/2020,Bulgaria/16/2020,Bulgaria/21/2020 | - |

| | | | |
|---|---|---|---|
| Elizabeth Batty et al | 9 | Thailand/Bangkok-CONI-0231/2020,Thailand/Bangkok-0084/2020,Thailand/Bangkok-0078/2020,Thailand/Bangkok-CONI-0322/2020,Thailand/Bangkok-CONI-0323/2020,Thailand/Bangkok-0077/2020,Thailand/Bangkok-0081/2020,Thailand/Bangkok-0071/2020,Thailand/Bangkok-0087/2020 | - |
| Ndongo Dia et al | 20 | Senegal/306/2020,Senegal/003/2020,Senegal/073/2020,Senegal/087/2020,Senegal/082/2020,Senegal/600/2020,Senegal/83403/2020,Senegal/65526/2020,Senegal/6427/2020,Senegal/3331/2020,Senegal/6481/2020,Senegal/9369/2020,Senegal/83493/2020,Senegal/64932/2020,Senegal/6430/2020,Senegal/68458/2020,Senegal/66458/2020,Senegal/45436/2020,Senegal/45557/2020,Senegal/62368/2020 | - |
| Jun Li et al | 3 | Hangzhou/HZCDC0001/2020,Hangzhou/HZCDC0048L/2020,Hangzhou/HZCDC0091L/2020 | - |
| Baowen Du et al | 7 | Sichuan/SC-WCH4-290/2020,Sichuan/SC-WCH4-286/2020,Wuhan/HB-WH2-160/2020,Wuhan/HB-WH1-122/2020,Wuhan/HB-WH1-142/2020,Wuhan/HB-WH4-204/2020,Wuhan/HB-WH4-206/2020 | - |
| Tsuyoshi Sekizuka et al C | 1 | Japan/OS-20-07-1/2020 | Whole genome and phylogenetic analysis of two SARS-CoV-2 strains isolated in Italy in January and February 2020: additional clues on multiple introductions and further circulation in Europe |
| Ji-Rong Yang et al C | 1 | Taiwan/4/2020 | Whole genome and phylogenetic analysis of two SARS-CoV-2 strains isolated in Italy in January and February 2020: additional clues on multiple introductions and further circulation in Europe |
| Jianjun Chen et al | 1 | Wuhan/0126-C12/2020 | - |
| Shiou-Hwei Yeh et al | 8 | Taiwan/NTU29/2020,Taiwan/NTU27/2020,Taiwan/NTU31/2020,Taiwan/NTU32/2020,Taiwan/NTU35/2020,Taiwan/NTU30/2020,Taiwan/NTU34/2020,Taiwan/NTU33/2020 | - |
| Monika Slávikova et al | 4 | Slovakia/SK-BMC6/2020,Slovakia/SK-BMC2/2020,Slovakia/SK-BMC5/2020,Slovakia/SK-BMC1/2020 | - |
| Christian Beisel et al | 37 | Switzerland/BL-ETHZ-140032/2020,Switzerland/VS-ETHZ-100326/2020,Switzerland/SZ-ETHZ-100348/2020,Switzerland/TG-ETHZ-370466/2020,Switzerland/ZH-ETHZ-300236/2020,Switzerland/BE-ETHZ-300264/2020,Switzerland/ZH-ETHZ-310422/2020,Switzerland/VD-ETHZ-300248/2020,Switzerland/BE-ETHZ-120237/2020,Switzerland/BE-ETHZ-120221/2020,Switzerland/TI-ETHZ-190011/2020,Switzerland/SZ-ETHZ-190054/2020,Switzerland/BL-ETHZ-170006/2020,Switzerland/BE-ETHZ-210073/2020,Switzerland/VD-ETHZ-280118/2020,Switzerland/BL-ETHZ-310480/2020,Switzerland/VD-ETHZ-230041/2020,Switzerland/BE-ETHZ-190066/2020,Switzerland/BL-ETHZ-120174/2020,Switzerland/BE-ETHZ-120233/2020,Switzerland/GE-ETHZ-240043/2020,Switzerland/SZ-ETHZ-160009/2020,Switzerland/SG-ETHZ-130013/2020,Switzerland/BL-ETHZ-240000/2020,Switzerland/UR-ETHZ-190001/2020,Switzerland/BL-ETHZ-170015/2020,Switzerland/SH-ETHZ-350188/2020,Switzerland/BS-ETHZ-130005/2020,Switzerland/ZH-ETHZ-240075/2020,Switzerland/ZH-ETHZ-250107/2020,Switzerland/SO-ETHZ-300258/2020,Switzerland/BE-ETHZ-370487/2020,Switzerland/ZH-ETHZ-150025plus/2020,Switzerland/VS-ETHZ-321460/2020,Switzerland/BL-ETHZ-270079/2020,Switzerland/BL-ETHZ-280151/2020,Switzerland/GR-ETHZ-170019/2020 | - |
| OH consortium et al | 5 | mink/Netherlands/NB-EMC-8-20/2020,mink/Netherlands/NB-EMC-28-9/2020,mink/Netherlands/NB-EMC-28-2/2020,mink/Netherlands/NB-EMC-28-7/2020,mink/Netherlands/NB-EMC-28-3/2020 | - |

| | | | |
|---|---|---|---|
| Teemu Smura et al | 16 | Finland/1/2020,Finland/8May24S14/2020,Finland/5May49S6/2020,Finland/5May26S2/2020,Finland/FIN-266/2020,Finland/15A36S5/2020,Finland/14M77/2020,Finland/29M14S1/2020,Finland/29M77S3/2020,Finland/10A63S14/2020,Finland/5May56S6/2020,Finland/30M77S3/2020,Finland/12A46SVI/2020,Finland/17A54S4/2020,Finland/4May88S4/2020,Finland/15A602/2020 | - |
| Siyuan Yang et al | 3 | Beijing/DT-WH01/2020,Beijing/DT-travelIT04/2020,Beijing/DT-travelUSA01/2020 | - |
| Shengyue Wang et al | 1 | Shanghai/SH0119/2020 | - |
| Chen YYC et al | 13 | Singapore/17nan/2020,Singapore/1135/2020,Singapore/1154/2020,Singapore/1172/2020,Singapore/1173/2020,Singapore/1165/2020,Singapore/1249/2020,Singapore/1269/2020,Singapore/1274/2020,Singapore/1264/2020,Singapore/1385/2020,Singapore/759/2020,Singapore/268nan/2020 | - |
| Bal et al | 1 | France/ARA-739/2020 | Molecular characterization of SARS-CoV-2 in the first COVID-19 cluster in France reveals an amino acid deletion in nsp2 (Asp268del) |
| Caspar Gross et al | 1 | Germany/BW-UKT-a118/2020 | - |
| Ying Tao et al | 6 | USA/CA-CDC-2602/2020,USA/WI-CDC-0946/2020,Guatemala/CDC-1236/2020,Guatemala/CDC-1227/2020,Guatemala/CDC-1230/2020,Guatemala/CDC-1231/2020 | - |
| Xianding Deng et al | 1 | USA/CA-SCCPHD-UC101/2020 | - |
| Benjamin Pinksy et al | 1 | USA/CA-CZB-1209/2020 | - |
| Timms V et al | 1 | Australia/NSW219/2020 | Revealing COVID-19 transmission in Australia by SARS-CoV-2 genome sequencing and agent-based modeling |
| Frilasita A Yudhaputri et al | 10 | Indonesia/JK-EIJK-39/2020,Indonesia/BT-EIJK36/2020,Indonesia/JK-EIJK34/2020,Indonesia/JK-EIJK24/2020,Indonesia/JK-EIJK42/2020,Indonesia/JB-EIJK41/2020,Indonesia/JK-EIJK46/2020,Indonesia/JK-EIJK-40/2020,Indonesia/JK-EIJK-23/2020,Indonesia/PP-EIJK-11/2020 | - |
| Edison Johar et al | 3 | Indonesia/JK-EIJK-07/2020,Indonesia/JK-EIJK-03/2020,Indonesia/JK-EIJK-02/2020 | - |
| Mendieta-Condado Edgar et al | 1 | Mexico/JAL-InDRE-16/2020 | - |
| Garces-Ayala Fabiola. Taboada Ramírez Blanca. Ramirez-Gonzalez Ernesto et al | 1 | Mexico/JAL-InDRE-11/2020 | - |
| Xianding Deng et al | 1 | USA/CA-CDPH-UC4/2020 | Genomic surveillance reveals multiple introductions of SARS-CoV-2 into Northern California |
| Juan David Ramírez et al | 6 | Colombia/ANT-GUV-92252/2020,Colombia/ANT-GUV-92146/2020,Colombia/HUI-GVI-98032/2020,Colombia/NAR-GVI-97417/2020,Colombia/VAC-GVI-97321/2020,Colombia/MET-GVI-97769/2020 | The arrival and spread of SARS-CoV2 in Colombia |
| Krisnoadi Rahardjo et al | 1 | Indonesia/JI-ITD-136N/2020 | - |
| Allam M et al | 2 | SouthAfrica/R02827/2020,SouthAfrica/LR00149/2020 | - |
| Martin Sundqvist et al | 1 | Sweden/20-50125/2020 | - |

| | | | |
|---|---|---|---|
| Neta Zuckerman et al | 22 | Israel/CVL-n2820/2020,Israel/n14289/2020,Israel/CVL-s2064/2020,Israel/CVL-s2049/2020,Israel/CVL-s2057/2020,Israel/CVL-n17916/2020,Israel/CVL-n19050/2020,Israel/CVL-n1541/2020,Israel/CVL-n896/2020,Israel/CVL-n1750/2020,Israel/CVL-n2361/2020,Israel/CVL-n-6751/2020,Israel/CVL-n25924/2020,Israel/CVL1610/2020,Israel/CVL-n26134/2020,Israel/CVL1060/2020,Israel/CVL-n25904/2020,Israel/CVL1196/2020,Israel/CVL-n26122/2020,Israel/CVL-n-6750/2020,Israel/CVL-n-6735/2020,Israel/CVL-n-11594/2020 | - |
| Rasmus Kirkegaard et al | 5 | Denmark/ALAB-SSI-1378/2020,Denmark/ALAB-HH-128/2020,Denmark/ALAB-HH-195/2020,Denmark/ALAB-HH15/2020,Denmark/ALAB-SSI-1388/2020 | - |
| Kuo-Chien Tsao et al | 5 | Taiwan/CGMH-CGU-22/2020,Taiwan/CGMH-CGU-36/2020,Taiwan/CGMH-CGU-33/2020,Taiwan/CGMH-CGU-32/2020,Taiwan/CGMH-CGU-31/2020 | - |
| Siu et al | 17 | HongKong/HKPU-0613/2020,HongKong/HKPU-0157/2020,HongKong/HKPU-5119/2020,HongKong/HKPU-5102/2020,HongKong/HKPU-5183/2020,HongKong/HKPU-5161/2020,HongKong/HKPU-5068/2020,HongKong/HKPU-1142/2020,HongKong/HKPU-5054/2020,HongKong/HKPU-4896/2020,HongKong/HKPU-4328/2020,HongKong/HKPU-5052/2020,HongKong/HKPU-4954/2020,HongKong/HKPU-5116/2020,HongKong/HKPU-1326/2020,HongKong/HKPU-1253/2020,HongKong/HKPU-1166/2020 | - |
| David Perera et al | 13 | Malaysia/UNIMAS-C122/2020,Malaysia/UNIMAS-C121/2020,Malaysia/UNIMAS-11942/2020,Malaysia/UNIMAS-14960/2020,Malaysia/UNIMAS-M4061/2020,Malaysia/UNIMAS-C012N/2020,Malaysia/UNIMAS-M3482/2020,Malaysia/UNIMAS-15288/2020,Malaysia/UNIMAS-0217/2020,Malaysia/UNIMAS-1020/2020,Malaysia/UNIMAS-1197/2020,Malaysia/UNIMAS-1589/2020,Malaysia/UNIMAS-15784/2020 | - |
| Jezzy R Dewantari et al | 1 | Indonesia/JI-ITD-150Sp/2020 | - |
| Pawestri et al | 8 | Indonesia/PA-NIHRD-C0710717/2020,Indonesia/PA-NIHRD-C0710684/2020,Indonesia/PA-NIHRD-PME4306/2020,Indonesia/KS-NIHRD-PME4900/2020,Indonesia/JK-NIHRD-MI52946/2020,Indonesia/JK-NIHRD-MI43269/2020,Indonesia/JK-NIHRD-MI52885/2020,Indonesia/JK-NIHRD-MI41613/2020 | - |
| Daniel F Gudbjartsson et al | 6 | Iceland/447/2020,Iceland/13/2020,Iceland/53/2020,Iceland/209/2020,Iceland/241/2020,Iceland/282/2020 | Spread of SARS-CoV-2 in the Icelandic Population |
| Gunadi et al | 6 | Indonesia/YO-UGM-200927/2020,Indonesia/JT-UGM-47964/2020,Indonesia/JT-UGM-48660/2020,Indonesia/YO-UGM-48651/2020,Indonesia/YO-UGM-107727/2020,Indonesia/JT-UGM-47906/2020 | - |
| Marcelo Henrique Santos Paiva et al | 37 | Brazil/PE-IAM138/2020,Brazil/PE-IAM1264/2020,Brazil/PE-IAM307/2020,Brazil/PE-IAM991/2020,Brazil/PE-IAM1126/2020,Brazil/PE-IAM356/2020,Brazil/PE-IAM67/2020,Brazil/PE-IAM965/2020,Brazil/PE-IAM1082/2020,Brazil/PE-IAM1254/2020,Brazil/PE-IAM103/2020,Brazil/PE-IAM87/2020,Brazil/PE-IAM1468/2020,Brazil/PE-IAM48/2020,Brazil/PE-IAM30/2020,Brazil/PE-IAM961/2020,Brazil/PE-IAM914/2020,Brazil/PE-IAM109/2020,Brazil/PE-IAM84/2020,Brazil/PE-IAM957/2020,Brazil/PE-IAM889/2020,Brazil/PE-IAM1127/2020,Brazil/PE-IAM992/2020,Brazil/PE-IAM1315/2020,Brazil/PE-IAM958/2020,Brazil/PE-IAM939/2020,Brazil/PE-IAM719/2020,Brazil/PE-IAM221/2020,Brazil/PE-IAM08/2020,Brazil/PE-IAM1149/2020,Brazil/PE-IAM89/2020,Brazil/PE-IAM1309/2020,Brazil/PE-IAM355/2020,Brazil/PE-IAM383/2020,Brazil/PE-IAM662/2020,Brazil/PE-IAM900/2020,Brazil/PE-IAM1148/2020 | - |

| | | | |
|---|---|---|---|
| Suppiah J et al | 24 | Malaysia/IMR_WC14227/2020,Malaysia/IMR-WI194/2020,Malaysia/IMR-WI196/2020,Malaysia/IMR-W195I/2020,Malaysia/IMR-WC253176/2020,Malaysia/IMR-WC268890/2020,Malaysia/IMR-WC249886/2020,Malaysia/IMR-WI310/2020,Malaysia/IMR-WC272624/2020,Malaysia/IMR-WC281546/2020,Malaysia/IMR-CV140277/2020,Malaysia/IMR-WI109/2020,Malaysia/IMR-WI085/2020,Malaysia/IMR-WI080/2020,Malaysia/IMR-WI147/2020,Malaysia/IMR-WI205/2020,Malaysia/IMR-WI206/2020,Malaysia/IMR-WI203/2020,Malaysia/IMR-WI120/2020,Malaysia/IMR-WI112/2020,Malaysia/IMR-WI115/2020,Malaysia/IMR-WI123/2020,Malaysia/IMR-WI133/2020,Malaysia/IMR-WI132/2020 | - |
| Sakshi Shambhavi et al | 1 | India/TG-CCMB-J300/2020 | - |
| Mohammad Uzzal Hossain et al | 1 | Bangladesh/NIB-BCSIR-02/2020 | - |
| Angelov et al. et al | 2 | Germany/BW-UT-005/2020,Germany/BW-UT-004/2020 | - |
| Daryl Domman et al | 2 | USA/WY-UNM-00136/2020,USA/NM-UNM-00850/2020 | - |
| Chisha Sikazwe et al | 1 | Australia/WA34/2020 | - |
| Tsuyoshi Sekizuka et al D | 3 | Japan/DP0134/2020,Japan/DP0121/2020,Japan/DP0827/2020 | Haplotype networks of SARS-CoV-2 infections in the Diamond Princess cruise ship outbreak |
| Takayuki Murata et al | 2 | Japan/FHU-CS27-0221/2020,Japan/FHU-CS84-0228/2020 | - |
| Inbar Cohen-Gihon et al | 1 | Israel/ISR_JP0320/2020 | Full genome viral sequences inform patterns of SARS-CoV-2 spread into and within Israel |
| Javed et al | 1 | Pakistan/Gilgit1/2020 | Insights into The Codon Usage Bias of 13 Severe Acute Respiratory Syndrome Coronavirus 2 (SARS-CoV-2) Isolates from Different Geo-locations |
| Sirous Zeinali et al | 1 | Iran/KHGRC-1.1-IPI-8206/2020 | - |
| Anna Majer et al | 3 | Canada/MB-NML-1150/2020,Canada/MB-NML-818/2020,Canada/MB-NML-1017/2020 | - |
| Mohammad Ali Khosravi et al | 1 | Iran/KHGRC-2-2162/2020 | - |
| Zohreh Fattahi et al | 10 | Iran/7650/2020,Iran/GRC-70/2020,Iran/GRC-43/2020,Iran/1600/2020,Iran/2297/2020,Iran/GRC-10582/2020,Iran/1325/2020,Iran/2024/2020,Iran/GRC-9673/2020,Iran/GRC-9695/2020 | - |
| Chandima Jeewandara et al | 16 | SriLanka/CDR142/2020,SriLanka/CDR-SL7066/2020,SriLanka/CDR503/2020,SriLanka/CDR-KK224/2020,SriLanka/CDR-KK57/2020,SriLanka/CDR-KK230/2020,SriLanka/GQC12/2020,SriLanka/C31262/2020,SriLanka/C27337/2020,SriLanka/C27607/2020,SriLanka/cov3576/2020,SriLanka/C28324/2020,SriLanka/MIN01/2020,SriLanka/WB4/2020,SriLanka/BIN44/2020,SriLanka/CDR1885/2020 | - |
| Chandima Jeewandara et al | 4 | SriLanka/COV38/2020,SriLanka/COV486/2020,SriLanka/COV91/2020,SriLanka/COV53/2020 | Evolutionary and genomic analysis of four SARS-CoV-2 isolates circulating in March 2020 in Sri Lanka; Additional evidence on multiple introduction and further transmission |
| Harrigan et al | 8 | Canada/BC_17397/2020,Canada/BC_13297/2020,Canada/BC_69243/2020,Canada/BC_9345715/2020,Canada/BC_8486790/2020,Canada/BC_9574898/2020,Canada/BC_7553799/2020,Canada/BC_6997898/2020 | - |
| Harrigan et al | 1 | Canada/BC_37_0-2/2020 | A doubt of multiple introduction of SARS-CoV-2 in Italy: A preliminary overview |

| | | | |
|---|---|---|---|
| Alexandra Gerber et al | 54 | Brazil/MT-0225/2020,Brazil/GO-0207/2020,Brazil/GO-0208/2020,Brazil/MG-0288/2020,Brazil/RJ-0251/2020,Brazil/GO-0209/2020,Brazil/CE-0206/2020,Brazil/AM-0201/2020,Brazil/AM-0204/2020,Brazil/AM-0202/2020,Brazil/MG-0212/2020,Brazil/PA-0229/2020,Brazil/MG-0221/2020,Brazil/RJ-0252/2020,Brazil/RJ-0253/2020,Brazil/PR-0241/2020,Brazil/MG-0216/2020,Brazil/MG-0220/2020,Brazil/PA-0237/2020,Brazil/MG-0287/2020,Brazil/MG-0214/2020,Brazil/AM-0203/2020,Brazil/AM-0205/2020,Brazil/PA-0227/2020,Brazil/RJ-0247/2020,Brazil/MG-0291/2020,Brazil/MG-0286/2020,Brazil/MG-0222/2020,Brazil/MG-0289/2020,Brazil/PA-0232/2020,Brazil/PI-0239/2020,Brazil/PA-0235/2020,Brazil/MG-0285/2020,Brazil/MG-0217/2020,Brazil/RJ-0249/2020,Brazil/MG-0258/2020,Brazil/RJ-0250/2020,Brazil/MG-0218/2020,Brazil/GO-0210/2020,Brazil/MG-0290/2020,Brazil/SC-0246/2020,Brazil/MG-0219/2020,Brazil/PA-0228/2020,Brazil/PA-0231/2020,Brazil/PA-0234/2020,Brazil/PA-0236/2020,Brazil/PA-0230/2020,Brazil/PA-0226/2020,Brazil/PE-0238/2020,Brazil/SC-0245/2020,Brazil/SC-0244/2020,Brazil/RS-0242/2020,Brazil/MG-0211/2020,Brazil/SC-0243/2020 | Evolution and epidemic spread of SARS-CoV-2 in Brazil |
| Junyoung Kim et al | 9 | SouthKorea/KCDC2658/2020,SouthKorea/KCDC2811/2020,SouthKorea/KCDC2813/2020,SouthKorea/KCDC2712/2020,SouthKorea/KCDC2666/2020,SouthKorea/KCDC2669/2020,SouthKorea/KCDC2681/2020,SouthKorea/KCDC2736/2020,SouthKorea/KCDC2687/2020 | - |
| Fahd Al-Mulla et al | 5 | Kuwait/KU17/2020,Kuwait/KU008/2020,Kuwait/KU005/2020,Kuwait/KU001/2020,Kuwait/KU006/2020 | - |
| Zhibing Yun et al | 1 | Sweden/20-04631/2020 | - |
| Nato Kotaria et al | 4 | Georgia/Tb-82/2020,Georgia/Tb-468/2020,Georgia/Tb-54/2020,Georgia/Tb-477/2020 | - |
| Rita Feghali et al | 5 | Lebanon/S5_762/2020,Lebanon/S9_764/2020,Lebanon/S7_763/2020,Lebanon/S1_758/2020,Lebanon/S4_761/2020 | - |
| Shevtsov et al | 3 | Kazakhstan/NCB-1/2020,Kazakhstan/NCB-2/2020,Kazakhstan/NCB-3/2020 | - |
| Al Wasti et al | 1 | Bahrain/140008296/2020 | - |
| Jalees A. Nasir et al | 1 | Canada/ON_SHSC-01/2020 | - |
| Tata Imnadze et al | 1 | Georgia/Tb-7851/2020 | - |
| Matt Storey et al | 1 | NewZealand/20VR0174/2020 | An emergent clade of SARS-CoV-2 linked to returned travellers from Iran |
| Paola Stefanelli et al | 2 | Italy/LAZ-INMI-SPL1/2020,Italy/LOM-ASST-CDG1/2020 | Whole genome and phylogenetic analysis of two SARS-CoV-2 strains isolated in Italy in January and February 2020: additional clues on multiple introductions and further circulation in Europe |
| Cesare E.M. Gruber et al | 1 | Italy/LAZ-INMI2-N/2020 | The first two cases of 2019-nCoV in Italy: Where they come from? |
| Tomaž Mark Zorec et al | 6 | Slovenia/V0-6934/2020,Slovenia/V0-15921/2020,Slovenia/V0-2870/2020,Slovenia/V0-24263/2020,Slovenia/V0-5244/2020,Slovenia/V0-3902/2020 | - |
| Borges et al et al | 26 | Portugal/PT0846/2020,Portugal/PT0783/2020,Portugal/PT0784/2020,Portugal/PT0974/2020,Portugal/PT0588/2020,Portugal/PT0448/2020,Portugal/PT1532/2020,Portugal/PT1032a/2020,Portugal/PT0844/2020,Portugal/PT1739/2020,Portugal/PT1703/2020,Portugal/PT1701/2020,Portugal/PT1705/2020,Portugal/PT1702/2020,Portugal/PT1710/2020,Portugal/PT0497/2020,Portugal/PT0496/2020,Portugal/PT1593/2020,Portugal/PT1613/2020,Portugal/PT1627/2020,Portugal/PT1628/2020,Portugal/PT0493/2020,Portugal/PT0491/2020,Portugal/PT0492/2020,Portugal/PT1608/2020,Portugal/PT1554/2020 | - |

| | | | |
|---|---|---|---|
| Ann Machablishvili et al | 1 | Georgia/Tb-537/2020 | - |
| Maria R. Capobianchi et al | 1 | Italy/LAZ-INMI1-cs/2020 | The first two cases of 2019-nCoV in Italy: Where they come from? |
| Iglesias-Caballero et al | 2 | Spain/AN-ISCIII-201272/2020,Spain/CL-ISCIII-201061/2020 | Phylodynamics of SARS-CoV-2 transmission in Spain |
| Nguyễn Thanh Long et al | 1 | Vietnam/PIHCM-503/2020 | - |
| Hege Vangstein Aamot et al | 6 | Norway/CS-24/2020,Norway/CS-09/2020,Norway/CS-23/2020,Norway/CS-04/2020,Norway/CS-26/2020,Norway/CS-02/2020 | - |
| Soares da Silva et al | 6 | Timor-Leste/TL25/2020,Timor-Leste/TL17/2020,Timor-Leste/TL16/2020,Timor-Leste/TL07/2020,Timor-Leste/TL21/2020,Timor-Leste/TL22/2020 | - |
| Malta et al | 2 | Brazil/SP02cc/2020,Brazil/UN-HIAE-SP04/2020 | - |
| Jaqueline Goes de Jesus et al | 2 | Brazil/SP-02/2020,Brazil/SP-01/2020 | First cases of coronavirus disease (COVID-19) in Brazil, South America |
| Claudio Tavares Sacchi et al | 1 | Brazil/SP-10/2020 | COVID-19 in Latin America: Contrasting phylodynamic inference with epidemiological surveillance. |
| Eduardo Juscamayta Lopez et al | 15 | Peru/LIM-INS-141/2020,Peru/LIM-INS-123/2020,Peru/LIM-INS-156/2020,Peru/ARE-INS-164/2020,Peru/CAL-INS-103/2020,Peru/LIM-INS-148/2020,Peru/LIM-INS-099/2020,Peru/LIM-INS-077/2020,Peru/LIM-INS-117/2020,Peru/LIM-INS-120/2020,Peru/LIM-INS-100/2020,Peru/LIM-INS-110/2020,Peru/LIM-INS-086/2020,Peru/LIM-INS-159/2020,Peru/LIM-INS-093/2020 | - |
| PHE Covid Sequencing Team et al | 15 | Ukraine/203100357/2020,Ukraine/203100321/2020,Ukraine/203100333/2020,Ukraine/203100348/2020,Ukraine/203100319/2020,Ukraine/203100362/2020,Ukraine/203100360/2020,Ukraine/203100355/2020,Ukraine/203100337/2020,Ukraine/203100361/2020,Ukraine/203100324/2020,Ukraine/203100317/2020,Ukraine/203100334/2020,Ukraine/203100338/2020,England/20192085601/2020 | - |
| Paola Resende et al | 18 | Brazil/ES-225/2020,Brazil/SC-766/2020,Brazil/SC-769/2020,Brazil/RJ-314/2020,Brazil/BA-510/2020,Brazil/BA-312/2020,Brazil/RJ-899/2020,Brazil/RJ-763/2020,Brazil/RJ-818/2020,Brazil/RJ-872/2020,Brazil/DF-861/2020,Brazil/DF-619i/2020,Brazil/DF-615i/2020,Brazil/DF-891/2020,Brazil/RJ-1119/2020,Brazil/RJ-1111/2020,Brazil/DF-862/2020,Brazil/AL-837/2020 | Genomic surveillance of SARS-CoV-2 reveals community transmission of a major lineage during the early pandemic phase in Brazil |
| Yan Li et al | 4 | Jamaica/JM-CDC-0869/2020,Jamaica/JM-CDC-6365/2020,Jamaica/JM-CDC-5836/2020,Jamaica/JM-CDC-4376/2020 | - |
| Joyce M. Ngoi et al | 1 | Ghana/2230_S4/2020 | - |
| Oluniyi P.E. et al | 2 | Nigeria/OS016-CV3/2020,Nigeria/OG007-CV22/2020 | SARS-CoV-2 Genomes from Nigeria Reveal Community Transmission, Multiple Virus Lineages and Spike Protein Mutation Associated with Higher Transmission and Pathogenicity |
| Kuenyoul Park et al | 1 | SouthKorea/S4/2020 | - |
| Ma. Angelica Tujan et al | 1 | Philippines/PH-RITM-0020/2020 | - |
| Carlo M. Lapid et al | 2 | Philippines/PGC03/2020,Philippines/PGC06/2020 | - |
| Ali et al | 1 | Pakistan/KP-RMI-01/2020 | - |
| Logan Voegtly et al | 3 | Guam/GU_NHG_01/2020,Guam/GU_NHG_03/2020,Guam/GU_NHG_02/2020 | - |
| Medado et al | 3 | Philippines/RITM-03/2020,Philippines/RITM-07/2020,Philippines/RITM-05/2020 | - |
| Mak Tze Minn et al | 5 | Brunei/1/2020,Brunei/3/2020,Brunei/2/2020,Brunei/5/2020,Brunei/4/2020 | - |

| | | | |
|---|---|---|---|
| Christian Ranaivoson et al | 6 | Madagascar/VIRO-2722/2020,Madagascar/VIRO-2728/2020,Madagascar/IP-01650/2020,Madagascar/IP-01861/2020,Madagascar/VIRO-33698/2020,Madagascar/VIRO-5815/2020 | - |
| Sesay et al et al | 2 | Gambia/GC19-026/2020,Gambia/GC19-015/2020 | Origin of imported SARS-CoV-2 strains in The Gambia identified from Whole Genome Sequences |
| Myat Htut Nyunt et al | 1 | Myanmar/MMC_137/2020 | - |
| Voegtly et al | 1 | Guam/GU-NHG-02/2020 | - |
| Masahiro Suzuki et al | 1 | Japan/FHU-Cor529/2020 | - |
| Fahad Zadjali et al | 9 | Oman/205037397/2020,Oman/205037001/2020,Oman/205038017/2020,Oman/205042917/2020,Oman/205035503/2020,Oman/205041214/2020,Oman/205043002/2020,Oman/205024123/2020,Oman/205029065/2020 | - |
| Ivan-Christian Kurolt et al | 4 | Croatia/1761_Dubrovnik/2020,Croatia/592_Osijek/2020,Croatia/1560_Split/2020,Croatia/1146_Split/2020 | - |
| Sujay Krishna Maity et al | 11 | India/WB-IICB-009/2020,India/WB-IICB-008/2020,India/WB-IICB-013/2020,India/WB-IICB-034/2020,India/WB-IICB-016/2020,India/WB-IICB-032/2020,India/WB-IICB-038/2020,India/WB-IICB-015/2020,India/WB-IICB-036/2020,India/WB-IICB-017/2020,India/WB-IICB-031/2020 | - |
| Sajjad Asaf et al | 2 | Oman/RESP-20-B-15349/2020,Oman/C-2673/2020 | - |
| Samira Al-Mahruqi et al | 1 | Oman/RESP-20-B-6588/2020 | - |
| Boehmer et al | 4 | Germany/BY-ChVir-1019/2020,Germany/BY-ChVir-1017/2020,Germany/BY-ChVir-1020/2020,Germany/BY-ChVir-929-2/2020 | - |
| Arash Iranzadeh et al | 9 | SouthAfrica/NHLS-UCT-GS-7016-KRISP/2020,SouthAfrica/NHLS-UCT-GP-5276/2020,SouthAfrica/NHLS-UCT-GP-5326/2020,SouthAfrica/NHLS-UCT-GS-2087/2020,SouthAfrica/NHLS-UCT-GS-2089/2020,SouthAfrica/NHLS-UCT-GS-3552/2020,SouthAfrica/NHLS-UCT-GS-3515/2020,SouthAfrica/NHLS-UCT-GS-3097/2020,SouthAfrica/NHLS-UCT-GP-5742/2020 | - |
| Talita Adelino et al | 11 | Brazil/MG-CV9/2020,Brazil/MG-CV49/2020,Brazil/MG-CV13/2020,Brazil/MG-CV42/2020,Brazil/MG-CV48/2020,Brazil/MG-CV33/2020,Brazil/MG-CV19/2020,Brazil/MG-CV16/2020,Brazil/MG-CV26/2020,Brazil/MG-CV8/2020,Brazil/MG-CV32/2020 | The ongoing COVID-19 epidemic in Minas Gerais, Brazil: insights from epidemiological data and SARS-CoV-2 whole genome sequencing. |
| Victor M Corman et al | 1 | Germany/BY-ChVir-929/2020 | Whole genome and phylogenetic analysis of two SARS-CoV-2 strains isolated in Italy in January and February 2020: additional clues on multiple introductions and further circulation in Europe |
| Teemu Smura et al | 1 | Finland/FIN-25/2020 | A doubt of multiple introduction of SARS-CoV-2 in Italy: A preliminary overview |
| Gabriela Sevillano et al | 1 | Ecuador/ZZ-SARS-1/2020 | - |
| Kathrin Kattler et al | 1 | Germany/SL-SU-OM005/2020 | - |

| | | | |
|---|---|---|---|
| Brazil-UK Centre for Arbovirus Discovery Diagnosis Genomics et al | 35 | Brazil/MG-L29-CD720/2020,Brazil/PR-L30-CD764/2020,Brazil/PE-L27-CD596/2020,Brazil/MA-L27-CD591/2020,Brazil/MA-L28-CD616/2020,Brazil/MA-L27-CD588/2020,Brazil/MG-L45-CD1172/2020,Brazil/SP-L25-CD648/2020,Brazil/MA-L28-CD618/2020,Brazil/SP-L33-CD855/2020,Brazil/RS-L27-CD595/2020,Brazil/RS-L29-CD727/2020,Brazil/ES-L28-CD615/2020,Brazil/CE-L28-CD614/2020,Brazil/RS-L29-CD737/2020,Brazil/MG-L30-CD757/2020,Brazil/PR-L30-CD760/2020,Brazil/SP-L39-CD1032/2020,Brazil/SP-L39-CD1035/2020,Brazil/SP-L25-CD645/2020,Brazil/SP-L25-CD665/2020,Brazil/SP-L38-CD1011/2020,Brazil/SP-L37-CD990/2020,Brazil/SP-L39-CD1042/2020,Brazil/MG-L2-MG07/2020,Brazil/SP-L36-CD952/2020,Brazil/SP-L39-CD1036/2020,Brazil/SP-L39-CD1052/2020,Brazil/SP-L32-CD825/2020,Brazil/SP-L31-CD812/2020,Brazil/SP-L25-CD666/2020,Brazil/SP-L33-CD856/2020,Brazil/SP-L36-CD942/2020,Brazil/SP-L36-CD945/2020,Brazil/SP-L39-CD1037/2020 | - |
| Vidanovic et al | 29 | Serbia/Pancevo-007910104/2020,Serbia/Pancevo-009210204/2020,Serbia/Subotica-101360605/2020,Serbia/Subotica-125991705/2020,Serbia/UE38-04/2020,Serbia/Zrenjanin-033961404/2020,Serbia/KV27-04/2020,Serbia/Subotica-198541706/2020,Serbia/Pancevo-210032506/2020,Serbia/NoviSad-212282506/2020,Serbia/Vrsac-253781007/2020,Serbia/Sabac-000230907/2020,Serbia/Sabac-000490807/2020,Serbia/Sabac-000012108/2020,Serbia/Sabac-000020508/2020,Serbia/KM-09962407/2020,Serbia/KM-0370707/2020,Serbia/KosovskaMitrovica-05221607/2020,Serbia/NoviSad-196221606/2020,Serbia/NoviSad-158472905/2020,Serbia/NS838-04/2020,Serbia/Kikinda-000201303/2020_,Serbia/NoviPazar-17611405/2020,Serbia/NoviPazar-17641405/2020,Serbia/KosovskaMitrovica-08372107/2020,Serbia/NP-0440907/2020,Serbia/NP-0541007/2020,Serbia/KosovskaMitrovica-09872307/2020,Serbia/Pancevo-215572606/2020 | - |
| Lauren Cowley et al | 3 | Bangladesh/G-27/2020,Bangladesh/G-38/2020,Bangladesh/G-183/2020 | - |
| Zekri et al | 11 | Egypt/CUNCI-HGC6I029/2020,Egypt/CUNCI-HGC6I010/2020,Egypt/CUNCI-HGC7I029/2020,Egypt/CUNCI-HGC7I025/2020,Egypt/CUNCI-HGC7I030/2020,Egypt/CUNCI-7I026/2020,Egypt/CUNCI-7I028/2020,Egypt/CUNCI-HGC023/2020,Egypt/CUNCI-HGC014/2020,Egypt/CUNCI-HGC015/2020,Egypt/CUNCI-HGC6I031/2020 | - |
| Nihad Al-Rashedi et al | 1 | Iraq/ICGEB-5T/2020 | - |
| AlWasti et al | 10 | Bahrain/920319237_S17_L001/2020,Bahrain/023285711_S13_L001/2020,Bahrain/023287015_S14_L001/2020,Bahrain/340286642_S20_L001/2020,Bahrain/340564387_S8_L001/2020,Bahrain/090000572_S16_L001/2020,Bahrain/340513774_S1_L001/2020,Bahrain/340861254_S20/2020,Bahrain/340861247_S22/2020,Bahrain/340859209_S2/2020 | - |
| Murat Karamese et al | 7 | Turkey/KU-019/2020,Turkey/KU-009/2020,Turkey/KU-017/2020,Turkey/KU-030/2020,Turkey/KU-023/2020,Turkey/KU-011/2020,Turkey/KU-026/2020 | - |
| Ilhem Boutiba-Ben Boubaker et al | 7 | Tunisia/61627/2020,Tunisia/55006/2020,Tunisia/2909/2020,Tunisia/9111/2020,Tunisia/61624/2020,Tunisia/55304/2020,Tunisia/3913/2020 | - |
| Saleh et al | 1 | Egypt/C-VSVRI-SERVAC/2020 | - |
| Roshdy et al | 1 | Egypt/C-CEIRS-19MOH/2020 | - |
| Alberto Paniz-Mondolfi et al | 2 | Venezuela/VEN-95072/2020,Venezuela/VEN-89312/2020 | - |

| | | | |
|---|---|---|---|
| Katherine Laiton-Donato et al | 31 | Colombia/ATL-INS-01/2020,Colombia/AMA-INS-105161/2020,Colombia/AMA-INS-106856/2020,Colombia/ANT-INS-01/2020,Colombia/ANT-INS-5/2020,Colombia/ATL-INS-04/2020,Colombia/DC-INS-197/2020,Colombia/QUI-INS-85082/2020,Colombia/BOY-INS-88871/2020,Colombia/NSA-NS-79852/2020,Colombia/ATL-INS-88712/2020,Colombia/AMA-INS-109363/2020,Colombia/AMA-INS-105142/2020,Colombia/ANT-INS-02/2020,Colombia/ATL-INS-06/2020,Colombia/DC-INS-289/2020,Colombia/DC-INS-3334/2020,Colombia/DC-INS-2553/2020,Colombia/ATL-INS-07/2020,Colombia/INS-CUN-AG2/2020,Colombia/DC-INS-05-07/2020,Colombia/DC-INS-2624/2020,Colombia/DC-INS-2640/2020,Colombia/DC-INS-027/2020,Colombia/DC-INS-3569/2020,Colombia/CUN-INS-96791/2020,Colombia/CAU-INS-89002/2020,Colombia/COR-INS-95319/2020,Colombia/ATL-INS-10/2020,Colombia/MET-INS-95637/2020,Colombia/NAR-INS-103313/2020 | - |
| Laifa Hendarmin et al | 1 | Indonesia/BT-SHSIU-01-4941/2020 | - |
| Erike A Suwarsono et al | 1 | Indonesia/BT-SHSIU-01-2566/2020 | - |
| Chris Adhiyanto et al | 2 | Indonesia/BT-SHSIU-01-4920/2020,Indonesia/BT-SHSIU-01-3610/2020 | - |
| Nikita S. D. Sahadeo et al | 8 | Trinidad/TT3347/2020,Trinidad/TT6949/2020,Trinidad/TT4707/2020,Trinidad/TT7867/2020,Trinidad/TT2238/2020,Trinidad/TT9467/2020,Trinidad/TT1327/2020,Trinidad/TT7923/2020 | - |
| Sara Mfarrej et al | 8 | SaudiArabia/KAUST-RIYADH1263/2020,SaudiArabia/KAUST-RIYADH1258/2020,SaudiArabia/KAUST-RIYADH1264/2020,SaudiArabia/KAUST-JEDDAH560/2020,SaudiArabia/KAUST-JEDDAH583/2020,SaudiArabia/KAUST-Jeddah60/2020,SaudiArabia/KAUST-JEDDAH529/2020,SaudiArabia/KAUST-JEDDAH590/2020 | - |
| Muhammad Shuaib et al | 2 | SaudiArabia/KAUST-RIYADH1229/2020,SaudiArabia/KAUST-MAKKAH1410/2020 | - |
| Alosaimi et al | 5 | SaudiArabia/656/2020,SaudiArabia/596/2020,SaudiArabia/671/2020,SaudiArabia/627/2020,SaudiArabia/610/2020 | - |
| Zeti Harriyati et al | 1 | Indonesia/BT-SHSIU-01-3884/2020 | - |
| Mohamed Ahmed Ali et al | 2 | Egypt/NRC-03/2020,Egypt/NRC-01/2020 | SARS-CoV-2 Genome Analysis of Japanese Travelers in Nile River Cruise |
| Pilailuk Okada et al | 14 | Thailand/Chonburi-SQ-TB0062/2020,Thailand/Bangkok-14712/2020,Thailand/Bangkok-PH7928/2020,Thailand/phitsanulok-NT0076/2020,Thailand/Chonburi-SQ-TS0001/2020,Thailand/Chonburi-SQ-NT0002/2020,Thailand/Chonburi-SQ-KC0001/2020,Thailand/Bangkok-14918/2020,Thailand/Bangkok-14765/2020,Thailand/Bangkok-Tho07/2020,Thailand/Chonburi-SQ-NP0073/2020,Thailand/Bangkok-Am01/2020,Thailand/Chonburi-SQ-NT0031/2020,Thailand/Chonburi-SQ-RF0001/2020 | - |
| Souissi et al | 9 | Tunisia/ADAGE-24763/2020,Tunisia/ADAGE-17149/2020,Tunisia/ADAGE-24755/2020,Tunisia/ADAGE-18212/2020,Tunisia/ADAGE-18505/2020,Tunisia/ADAGE-17873/2020,Tunisia/ADAGE-15425/2020,Tunisia/ADAGE-12622/2020,Tunisia/ADAGE-18245/2020 | - |
| Alexandra Popa et al | 17 | Austria/CeMM1083/2020,Austria/CeMM0648/2020,Austria/CeMM0912/2020,Austria/CeMM1077/2020,Austria/CeMM1078/2020,Austria/CeMM0768/2020,Austria/CeMM0828/2020,Austria/CeMM0767/2020,Austria/CeMM0395/2020,Austria/CeMM1094/2020,Austria/CeMM1095/2020,Austria/CeMM1092/2020,Austria/CeMM1091/2020,Austria/CeMM1096/2020,Austria/CeMM0909/2020,Austria/CeMM0392/2020,Austria/CeMM1084/2020 | - |
| Samina Naz Mukry et al | 2 | Pakistan/NIBD-04-KHI/2020,Pakistan/NIBD-02-KHI/2020 | - |

| | | | |
|---|---|---|---|
| Xiaoyun Ren et al | 44 | NewZealand/20VR3494/2020,NewZealand/20CV0063/2020,NewZealand/20CV0628/2020,NewZealand/20CV0629/2020,NewZealand/20VR3068/2020,NewZealand/20VR3172/2020,NewZealand/20VR3128/2020,NewZealand/20VR3476/2020,NewZealand/20VR3755/2020,NewZealand/20VR3759/2020,NewZealand/20CV0637/2020,NewZealand/20VR1835/2020,NewZealand/20VR3909/2020,NewZealand/20CV0066/2020,NewZealand/20VR3879/2020,NewZealand/20VR3903/2020,NewZealand/20CV0229/2020,NewZealand/20CV0181/2020,NewZealand/20CV0217/2020,NewZealand/20CV0220/2020,NewZealand/20CV0246/2020,NewZealand/20CV0235/2020,NewZealand/20CV0230/2020,NewZealand/20CV0627/2020,NewZealand/20VR2060/2020,NewZealand/20CV0510/2020,NewZealand/20CV0587/2020,NewZealand/20VR3802/2020,NewZealand/20VR3804/2020,NewZealand/20VR3828/2020,NewZealand/20VR3060/2020,NewZealand/20CV0496/2020,NewZealand/20CV0631/2020,NewZealand/20CV0653/2020,NewZealand/20VR3066/2020,NewZealand/20VR3119/2020,NewZealand/20VR1888/2020,NewZealand/20CV0388/2020,NewZealand/20VR1962/2020,NewZealand/20CV0449/2020,NewZealand/20CV0515/2020,NewZealand/20CV0516/2020,NewZealand/20CV0541/2020,NewZealand/20CV0595/2020 | - |
| Foster et al | 6 | Australia/NSW3642/2020,Australia/NSW3644/2020,Australia/NSW3629/2020,Australia/NSW3628/2020,Australia/NSW2603/2020,Australia/NSW3647/2020 | - |
| Zahra Ahmadi et al | 1 | Iran/9GS/2020 | - |
| Kamolthip Atsawawaranunt et al | 4 | Thailand/Bangkok-CONI-0319/2020,Thailand/Bangkok-CONI-0329/2020,Thailand/Bangkok-CONI-0325/2020,Thailand/Bangkok-CONI-0331/2020 | - |
| Jing Zhang et al | 5 | Guatemala/CDC-1229/2020,Belize/CDC-6852/2020,Belize/CDC-6851/2020,Belize/CDC-6840/2020,Belize/CDC-6846/2020 | - |
| Robert Paulino-Ramirez et al | 4 | DominicanRepublic/ICGEB_UNIBE032.2/2020,DominicanRepublic/ICGEB_UNIBE032.7/2020,DominicanRepublic/ICGEB_UNIBE022/2020,DominicanRepublic/ICGEB_UNIBE258/2020 | - |
| Jorge González et al | 8 | Chile/MA-CADIUMAG-37/2020,Chile/MA-CADIUMAG-18/2020,Chile/MA-CADIUMAG-22/2020,Chile/MA-CADIUMAG-23/2020,Chile/MA-CADIUMAG-17/2020,Chile/MA-CADIUMAG-25/2020,Chile/MA-CADIUMAG-26/2020,Chile/MA-CADIUMAG-24/2020 | - |
| Seadawy et al | 6 | Egypt/Army-MCL001/2020,Egypt/HCoV2-Egy-002/2020,Egypt/EGY-028/2020,Egypt/EGY-029/2020,Egypt/EGY-046/2020,Egypt/EGY-050/2020 | - |
| Antoine Dara et al | 1 | Mali/M00960/2020 | - |
| Dominika Fričová et al | 1 | Slovakia/UKBA-101/2020 | - |
| Nagy et al | 38 | CzechRepublic/NRL-7322-3/2020,CzechRepublic/NRL-7322-4/2020,CzechRepublic/NRL_5815/2020,CzechRepublic/NRL-6047/2020,CzechRepublic/NRL_7000/2020,CzechRepublic/NRL-7322-2/2020,CzechRepublic/NRL-10712/2020,CzechRepublic/NRL-5593/2020,CzechRepublic/NRL_8460/2020,CzechRepublic/NRL_8972-5/2020,CzechRepublic/NRL-10717/2020,CzechRepublic/NRL-10720/2020,CzechRepublic/NRL_9893/2020,CzechRepublic/NRL_8972-3/2020,CzechRepublic/NRL_9448/2020,CzechRepublic/NRL-6849-8/2020,CzechRepublic/NRL_6849-10/2020,CzechRepublic/NRL_6849-7/2020,CzechRepublic/NRL_6849-11/2020,CzechRepublic/NRL-6847-2/2020,CzechRepublic/NRL_8459/2020,CzechRepublic/NRL-10710/2020,CzechRepublic/NRL_8972-1/2020,CzechRepublic/NRL_9713/2020,CzechRepublic/NRL-10733/2020,CzechRepublic/NRL_8458/2020,CzechRepublic/NRL_9654/2020,CzechRepublic/NRL-10549/2020,CzechRepublic/NRL_7917/2020,CzechRepublic/NRL-6833/2020,CzechRepublic/NRL_7164/2020,CzechRepublic/NRL-7913/2020,CzechRepublic/NRL-7915/2020,CzechRepublic/NRL-7950/2020,CzechRepublic/NRL_8118-4/2020,CzechRepublic/NRL_7949/2020,CzechRepublic/NRL_8118-3/2020,CzechRepublic/NRL_9883/2020 | - |

| | | | |
|---|---|---|---|
| Angel Angelov et al | 8 | Congo/UKT-008/2020,Congo/UKT-005/2020,Congo/UKT-014/2020,Congo/UKT-015/2020,Congo/UKT-013/2020,Congo/UKT-002/2020,Congo/UKT-009/2020,Germany/BW-UT-018/2020 | - |
| Allison McGeer et al | 12 | Canada/ON-S1285/2020,Canada/ON-S1317/2020,Canada/ON-S1821/2020,Canada/ON-S1852/2020,Canada/ON-S1805/2020,Canada/ON-S854/2020,Canada/ON-S1806/2020,Canada/ON-S1481/2020,Canada/ON-S1430/2020,Canada/ON-S1431/2020,Canada/ON-S1474/2020,Canada/ON-S1475/2020 | - |
| Biazzo et al | 8 | Malta/BAL-Sliema-7/2020,Malta/BAL-Sliema-5/2020,Malta/BAL-Sliema-1/2020,Malta/BAL-Sliema-2/2020,Malta/BAL-Sliema-3/2020,Malta/BAL-Sliema-4/2020,Malta/BAL-Sliema-9/2020,Malta/BAL-Sliema-8/2020 | - |
| Alwasti et al | 10 | Bahrain/920407807/2020,Bahrain/340685804/2020,Bahrain/920396988_S5_L001/2020,Bahrain/023341084/2020,Bahrain/340654690/2020,Bahrain/341017320/2020,Bahrain/250182773/2020,Bahrain/250182775/2020,Bahrain/250182774/2020,Bahrain/250182770_S11_L001/2020 | - |
| Anke Wienecke-Baldacchino et al | 16 | Luxembourg/LNS1586475/2020,Luxembourg/LNS1646752/2020,Luxembourg/LNS3156434/2020,Luxembourg/LNS4836560/2020,Luxembourg/LNS2128808/2020,Luxembourg/LNS2458673/2020,Luxembourg/LNS5413802/2020,Luxembourg/LNS6854244/2020,Luxembourg/LNS0113568/2020,Luxembourg/LNS4818581/2020,Luxembourg/LNS9470500/2020,Luxembourg/LNS0000001/2020,Luxembourg/LNS6733162/2020,Luxembourg/LNS7342327/2020,Luxembourg/LNS3059908/2020,Luxembourg/LNS1368952/2020 | - |
| Zaed et al | 3 | Bahrain/Hu-1/2020,Bahrain/110003611/2020,Bahrain/02/2020 | - |
| Marion Barbet et al | 19 | SaintBarthelemy/IPG-7358/2020,Guadeloupe/IPG-7331/2020,Guadeloupe/IPG-7323/2020,Guadeloupe/IPG-7319/2020,Guadeloupe/IPG-7336/2020,SaintMartin/IPG-7343/2020,SaintMartin/IPG-7356/2020,SaintBarthelemy/IPG-7329/2020,Guadeloupe/IPG-7324/2020,SaintMartin/IPG-7352/2020,SaintMartin/IPG-7354/2020,SaintMartin/IPG-7351/2020,SaintMartin/IPG-7348/2020,SaintMartin/IPG-7349/2020,SaintMartin/IPG-7355/2020,SaintMartin/IPG-7357/2020,France/IDF-9449/2020,France/BRE-BR9068/2020,France/BRE-IPP9983/2020 | - |
| Thanh Le Viet et al | 4 | Zimbabwe/ZW-25/2020,Zimbabwe/ZW-29/2020,Zimbabwe/ZW-6970/2020,Zimbabwe/ZW-1193B/2020 | - |
| Mélanie Albert et al C | 3 | Algeria/G0860_2262/2020,Algeria/G0640_2265/2020,Algeria/G0638_2264/2020 | - |
| Anthony LEVASSEUR et al | 12 | France/PAC-IHU-0941/2020,France/PAC-IHU-0928/2020,France/PAC-IHU-1131/2020,France/PAC-MEPHI-2018/2020,France/PAC-IHU-1448/2020,France/PAC-IHU-0955/2020,France/PAC-IHU-1632/2020,France/PAC-MEPHI-2394/2020,France/PAC-IHU-0971/2020,France/PAC-IHU-1589/2020,France/PAC-MEPHI-2375/2020,France/PAC-MEPHI-2390/2020 | - |
| Kefentse Arnold Tumedi et al | 6 | Botswana/BOT2283/2020,Botswana/BOT0455/2020,Botswana/BOT2624/2020,Botswana/BOT2254/2020,Botswana/BOT4161/2020,Botswana/BOT2855/2020 | - |

| | | | |
|---|---|---|---|
| Paola Resende et al | 104 | Brazil/SE-6591/2020,Brazil/SE-6555/2020,Brazil/SE-6583/2020,Brazil/SE-6568/2020,Brazil/PR-5623/2020,Brazil/PR-5618/2020,Brazil/PR-5622/2020,Brazil/SC-771/2020,Brazil/RS-2556/2020,Brazil/PR-5619/2020,Brazil/RS-2525/2020,Brazil/SE-6535/2020,Brazil/SE-6606/2020,Brazil/SE-6608/2020,Brazil/SE-6607/2020,Brazil/SE-6549/2020,Brazil/SE-6557/2020,Brazil/SE-6539/2020,Brazil/SE-6550/2020,Brazil/SE-6594/2020,Brazil/RS-6218/2020,Brazil/RS-6226/2020,Brazil/RS-15274/2020,Brazil/RS-6243/2020,Brazil/PR-5617/2020,Brazil/RS-2550/2020,Brazil/RS-2564/2020,Brazil/SE-6601/2020,Brazil/RS-6189/2020,Brazil/RS-6195/2020,Brazil/RS-6192/2020,Brazil/RS-2567/2020,Brazil/SE-6536/2020,Brazil/SE-6556/2020,Brazil/SC-770/2020,Brazil/SE-6579/2020,Brazil/RS-6205/2020,Brazil/RS-6196/2020,Brazil/RS-6184/2020,Brazil/RS-6177/2020,Brazil/RS-15280/2020,Brazil/RS-6203/2020,Brazil/RS-6188/2020,Brazil/RS-6208/2020,Brazil/RS-6222/2020,Brazil/RS-6231/2020,Brazil/RS-15273/2020,Brazil/RS-6169/2020,Brazil/RS-15288/2020,Brazil/RS-2544/2020,Brazil/RS-6240/2020,Brazil/RS-6227/2020,Brazil/RS-2546/2020,Brazil/RS-2554/2020,Brazil/RS-6179/2020,Brazil/RS-6242/2020,Brazil/RS-6228/2020,Brazil/RS-6197/2020,Brazil/RS-6198/2020,Brazil/RS-15287/2020,Brazil/RS-15276/2020,Brazil/RS-6180/2020,Brazil/RS-6187/2020,Brazil/RS-6190/2020,Brazil/RS-6241/2020,Brazil/SE-6563/2020,Brazil/RS-2533/2020,Brazil/RS-15282/2020,Brazil/RS-15283/2020,Brazil/RS-6219/2020,Brazil/RS-6220/2020,Brazil/RS-2565/2020,Brazil/RJ-1953/2020,env/Brazil/RJ-4736/2020,Brazil/SE-6529/2020,Brazil/RS-15270/2020,Brazil/RS-15284/2020,Brazil/SE-6567/2020,Brazil/RS-15275/2020,Brazil/RS-15279/2020,Brazil/RS-2553/2020,Brazil/RS-2529/2020,Brazil/PR-5620/2020,Brazil/RS-2539/2020,Brazil/PR-5621/2020,Brazil/RS-15286/2020,Brazil/RS-2528/2020,Brazil/RS-6232/2020,Brazil/RS-15290/2020,Brazil/RS-15289/2020,Brazil/RS-15281/2020,Brazil/RS-15292/2020,Brazil/SE-6530/2020,Brazil/SE-6603/2020,Brazil/SE-6561/2020,Brazil/RS-6215/2020,Brazil/RS-6183/2020,Brazil/RS-6213/2020,Brazil/RS-15285/2020,Brazil/RS-15291/2020,Brazil/RS-15278/2020,Brazil/RS-2549/2020,Brazil/SE-6533/2020,Brazil/SE-6544/2020 | - |
| Cristian Rohr et al | 2 | Argentina/Heritas_HG009/2020,Argentina/Heritas-HG023/2020 | - |
| Dalmacio Pereyra et al | 5 | Argentina/Heritas_HG019/2020,Argentina/Heritas_HG015/2020,Argentina/Heritas_HG014/2020,Argentina/Heritas_HG018/2020,Argentina/Heritas_HG001/2020 | - |
| Joaquín Ezpeleta et al | 10 | Argentina/argenTAG-127F2009/2020,Argentina/argenTAG-12926008/2020,Argentina/argenTAG-12CF5045/2020,Argentina/argenTAG-1252C02E/2020,Argentina/argenTAG-12568054/2020,Argentina/argenTAG-1283F02A/2020,Argentina/argenTAG-12CB103F/2020,Argentina/argenTAG-12C0A026/2020,Argentina/argenTAG-12E7E043/2020,Argentina/argenTAG-12BBC055/2020 | - |
| Githinji et al 2020 et al | 8 | Kenya/C21582/2020,Kenya/C4214/2020,Kenya/C3739/2020,Kenya/C5289/2020,Kenya/C4691/2020,Kenya/C6380/2020,Kenya/C4195/2020,Kenya/C6255/2020 | - |
| Rongbao Gao et al | 1 | Beijing/NPRC0004/2020 | - |
| Nabaes Jodar et al | 4 | Argentina/PAIS-A0003/2020,Argentina/PAIS-A0008/2020,Argentina/PAIS-A0015/2020,Argentina/PAIS-A0022/2020 | - |
| Gvantsa Brachveli et al | 2 | Georgia/Tb-3118/2020,Georgia/Tb-6572/2020 | - |
| Ana Papkiauri et al | 1 | Georgia/Tb-7856/2020 | - |
| Nino Berishvili et al | 1 | Georgia/Tb-5727/2020 | - |
| Meri Pantsulaia et al | 2 | Georgia/Tb-6598/2020,Georgia/Tb-27822/2020 | - |
| Meriem LAAMARTI et al | 2 | Morocco/RMPS-06/2020,Morocco/RMPS-05/2020 | Genetic analysis of SARS-CoV-2 strains collected from North Africa: viral origins and mutational spectrum |
| Rivera NR et al | 1 | ElSalvador/INS-01/2020 | - |

| | | | |
|---|---|---|---|
| Ismael N et al | 5 | Mozambique/KRISP-MZ157469/2020,Mozambique/KRISP-MZ154276/2020,Mozambique/KRISP-MZ158971/2020,Mozambique/KRISP-MZ155093/2020,Mozambique/KRISP-MZ157470/2020 | - |
| Giorgi Tomashvili et al | 1 | Georgia/Tb-22208/2020 | - |
| Filip Rokić et al | 4 | Croatia/I7-S21new/2020,Croatia/OY-S1new/2020,Croatia/LG-S2new/2020,Croatia/AU-S10new/2020 | - |
| Francisco Duarte et al | 17 | CostaRica/INC-0123/2020,CostaRica/INC-0128/2020,CostaRica/INC-0052/2020,CostaRica/INC-0048/2020,CostaRica/INC-0064/2020,CostaRica/INC-0094/2020,CostaRica/INC-0055/2020,CostaRica/INC-0078/2020,CostaRica/INC-0056/2020,CostaRica/INC-0129/2020,CostaRica/INC-0130/2020,CostaRica/INC-0136/2020,CostaRica/INC-0067/2020,CostaRica/INC-0060/2020,CostaRica/INC-0061/2020,CostaRica/INC-0082/2020,CostaRica/INC-0069/2020 | - |
| Danish Covid-19 Genome Consortium et al | 12 | Denmark/DCGC-12971/2020,Denmark/DCGC-12023/2020,Denmark/DCGC-10565/2020,Denmark/DCGC-12910/2020,Denmark/DCGC-11369/2020,Denmark/DCGC-11684/2020,Denmark/DCGC-9851/2020,Denmark/DCGC-13007/2020,Denmark/DCGC-10672/2020,Denmark/DCGC-12992/2020,Denmark/DCGC-12982/2020,Denmark/DCGC-10522/2020 | - |
| David Brandt et al | 1 | Germany/NW-UBI-25/2020 | - |
| Thomas Günther et al | 1 | Germany/NW-MPP-26/2020 | - |
| Anna-Malin Linde et al | 27 | Sweden/20-23091/2020,Sweden/20-50088/2020,Sweden/20-52462/2020,Sweden/20-22115/2020,Sweden/20-50220/2020,Sweden/20-52045/2020,Sweden/20-08987/2020,Sweden/20-08830/2020,Sweden/20-50257/2020,Sweden/20-08988/2020,Sweden/20-51952/2020,Sweden/20-22323/2020,Sweden/20-08986/2020,Sweden/20-52053/2020,Sweden/20-08950/2020,Sweden/20-53266/2020,Sweden/20-52827/2020,Sweden/20-52288/2020,Sweden/20-53091/2020,Sweden/20-53168/2020,Sweden/20-53166/2020,Sweden/20-52391/2020,Sweden/20-52653/2020,Sweden/20-52769/2020,Sweden/20-52767/2020,Sweden/20-52766/2020,Sweden/20-52425/2020 | - |
| Morten Rasmussen et al | 4 | Denmark/SSI-04/2020,Denmark/SSI-09/2020,Denmark/SSI-02/2020,Denmark/SSI-01/2020 | - |
| Mélnie Albert et al | 2 | France/NOR-N1620/2020,France/NOR-N2223/2020 | Introductions and early spread of SARS-CoV-2 in France |
| LAUBSCHER Florian et al. et al | 1 | Switzerland/TI-SNRCI-29919486/2020 | - |
| Madlen Stange et al | 5 | Switzerland/BL-UHB-42192884/2020,Switzerland/BS-UHB-42202167/2020,Switzerland/BS-UHB-42203665/2020,Switzerland/BS-42216750/2020,Switzerland/BS-42232763/2020 | - |
| Max Muenchhoff et al | 6 | Germany/BY-MVP-0113/2020,Germany/BY-MVP-0059/2020,Germany/BY-MVP-0259/2020,Germany/BY-MVP-0101/2020,Germany/BY-MVP-0100/2020,Germany/BY-MVP-0253/2020 | - |
| Dr V A Potdar et al | 1 | India/MH-CCMB-NIV5/2020 | A distinct phylogenetic cluster of Indian SARS-CoV-2 isolates |
| Jan Richter et al | 8 | Cyprus/007/2020,Cyprus/001/2020,Cyprus/002/2020,Cyprus/004/2020,Cyprus/003/2020,Cyprus/006/2020,Cyprus/008/2020,Cyprus/005/2020 | - |
| Rawlinson et al | 3 | Australia/NSW2158/2020,Australia/NSW2435/2020,Australia/NSW2411/2020 | - |
| Olympia E. Anastasiou et al | 1 | Germany/NW-HHU-163/2020 | - |
| Vicente Soriano Chirona et al | 1 | Spain/VC-FISABIO-305/2020 | - |
| Laurens Lambrechts et al | 4 | Belgium/UGent-129/2020,Belgium/UGent-297/2020,Belgium/UGent-251/2020,Belgium/UGent-165/2020 | - |
| Philippe Selhorst et al | 1 | Belgium/ITM_C237/2020 | - |

| | | | |
|---|---|---|---|
| David Nieuwenhuijse et al | 3 | Netherlands/Utrecht_3/2020,Netherlands/Diemen_1363454/2020,Netherlands/Helmond_1363548/2020 | Rapid SARS-CoV-2 whole genome sequencing for informed public health decision making in the Netherlands |
| Laubscher F. et al | 4 | Switzerland/GE-HUG-1184/2020,Switzerland/GE-HUG-1422/2020,Switzerland/GE-HUG-1402/2020,Switzerland/AG-HUG-7120/2020 | - |
| Lorusso A et al | 11 | Italy/ABR-IZSGC-TE5551/2020,Italy/ABR-IZSGC-TE99753/2020,Italy/ABR-IZSGC-TE5268/2020,Italy/ABR-IZSGC-TE6202/2020,Italy/ABR-IZSGC-TE6218/2020,Italy/ABR-IZSGC-TE18513/2020,Italy/LOM-IZSGC-TE35432/2020,Italy/LOM-IZSGC-TE35441/2020,Italy/LOM-IZSGC-TE30933/2020,Italy/LOM-IZSGC-TE35578/2020,Italy/LOM-IZSGC-TE35566/2020 | - |
| Ana da Silva Filipe et al | 2 | Scotland/CVR355/2020,Scotland/CVR3415/2020 | Preliminary analysis of SARS-CoV-2 importation & establishment of UK transmission lineages |
| Luke W Meredith et al | 2 | England/CAMB-743FF/2020,England/CAMB-78407/2020 | Rapid implementation of SARS-CoV-2 sequencing to investigate cases of health-care associated COVID-19: a prospective genomic surveillance study |
| Guillermo Martín Gutiérrez et al | 1 | Spain/AN-IBV-001106/2020 | - |
| CZB Cliahub Consortium et al | 4 | USA/CA-CZB089/2020,USA/CA-CZB-3699/2020,USA/CA-CZB-6987/2020,USA/CA-CZB-13465/2020 | - |
| Kassela K. et al | 2 | Greece/56_37161/2020,Greece/264_32497/2020 | Dominant and rare SARS-Cov2 variants responsible for the COVID-19 pandemic in Athens, Greece. |
| Jonas Schmidt et al | 1 | Germany/BW-MVZ-LS-12/2020 | - |
| Tanya Golubchik et al | 10 | England/OXON-B2C90/2020,England/OXON-B1E22/2020,England/OXON-AF7C2/2020,England/QEUH-A4C147/2020,England/ALDP-9BCA48/2020,England/ALDP-9BC9B4/2020,England/ALDP-9D7810/2020,England/ALDP-9D9034/2020,England/ALDP-9D8FED/2020,England/QEUH-A619F9/2020 | - |
| Blankenship HM et al | 1 | USA/MI-MDHHS-SC20833/2020 | - |
| Abi Habib et al | 2 | Lebanon/LB-R8/2020,Lebanon/LB-LR9/2020 | - |
| Sergey A. Bodnev et al | 2 | Crimea/SRC-80603/2020,Russia/Tatarstan-82208/2020 | - |
| David Nieuwenhuijse et al | 1 | Luxembourg/Lux1/2020 | - |
| Naranzul Ts et al | 1 | Mongolia/Imp3989/2020 | - |
| Juscamayta et al | 11 | Peru/UN-INS-044/2020,Peru/UN-INS-025/2020,Peru/UN-INS-021/2020,Peru/UN-INS-004/2020,Peru/UN-INS-001/2020,Peru/UN-INS-038/2020,Peru/UN-INS-041/2020,Peru/UN-INS-022/2020,Peru/UN-INS-032/2020,Peru/UN-INS-083/2020,Peru/UN-INS-015/2020 | - |
| Belén Prado-Vivar et al | 32 | Ecuador/USFQ-139/2020,Ecuador/USFQ-039/2020,Ecuador/USFQ-110/2020,Ecuador/USFQ-1112/2020,Ecuador/USFQ-197/2020,Ecuador/USFQ-127/2020,Ecuador/USFQ-126/2020,Ecuador/USFQ-220/2020,Ecuador/USFQ-344/2020,Ecuador/USFQ-114/2020,Ecuador/USFQ-111/2020,Ecuador/USFQ-118/2020,Ecuador/USFQ-119/2020,Ecuador/USFQ-109/2020,Ecuador/USFQ-108/2020,Ecuador/USFQ-105/2020,Ecuador/USFQ-381/2020,Ecuador/USFQ-193/2020,Ecuador/USFQ-203/2020,Ecuador/USFQ-201/2020,Ecuador/USFQ-054/2020,Ecuador/USFQ-066/2020,Ecuador/USFQ-120/2020,Ecuador/USFQ-433/2020,Ecuador/USFQ-253/2020,Ecuador/USFQ-346/2020,Ecuador/USFQ-405/2020,Ecuador/USFQ-558/2020,Ecuador/USFQ-065/2020,Ecuador/USFQ-147/2020,Ecuador/USFQ-1188/2020,Ecuador/USFQ-322/2020 | - |
| Gianguglielmo Zehender et al | 1 | Italy/LOM-UniMI03/2020 | Genomic characterization and phylogenetic analysis of SARS-COV-2 in Italy |
| Antonino Di Caro et al | 2 | Italy/LOM-INMI-5925/2020,Italy/LOM-INMI-9491/2020 | - |

| | | | |
|---|---|---|---|
| Hajar Lemriss et al | 1 | Morocco/refstage1/2020 | - |
| Samo Zakotnik et al | 5 | Slovenia/5028/2020,Slovenia/4915/2020,Slovenia/4584/2020,Slovenia/7360/2020,Slovenia/5070/2020 | - |
| Duarte et al | 1 | CostaRica/06/2020 | - |
| Broňa Brejová et al | 7 | Slovakia/UKBA-210/2020,Slovakia/UKBA-321/2020,Slovakia/UKBA-411/2020,Slovakia/UKBA-209/2020,Slovakia/UKBA-319/2020,Slovakia/UKBA-212/2021,Slovakia/UKBA-409/2020 | - |
| Githinji G. et al 2020 et al | 10 | Kenya/C22/2020,Kenya/C54/2020,Kenya/C58/2020,Kenya/C787/2020,Kenya/C1751/2020,Kenya/C2826/2020,Kenya/C2469/2020,Kenya/NIC_196/2020,Kenya/NIC_569/2020,Kenya/C358/2020 | Introduction and local transmission of SARS-CoV-2 cases in Kenya |
| Gaete A et al | 6 | Chile/RM-CMM-25669/2020,Chile/RM-CMM-05230/2020,Chile/RM-CMM-23741/2020,Chile/AT-CMM-00063/2020,Chile/AT-CMM-00064/2020,Chile/AT-CMM-00007/2020 | - |
| M.Lazar et al | 4 | Romania/284508/2020,Romania/285388/2020,Romania/284783/2020,Romania/284762/2020 | Whole-Genome Sequences of the Severe Acute Respiratory Syndrome Coronavirus-2 obtained from Romanian patients between March and June of 2020 |
| Andres Moreira-Soto et al | 6 | CostaRica/CRV-0023/2020,CostaRica/CRV-0020/2020,CostaRica/CRV-0018/2020,CostaRica/CRV-0024/2020,CostaRica/CRV-0022/2020,CostaRica/CRV-0017/2020 | - |
| Viktória Hodorová et al | 8 | Slovakia/UKBA-410/2020,Slovakia/UKBA-403/2020,Slovakia/UKBA-204/2020,Slovakia/UKBA-207/2020,Slovakia/UKBA-322/2020,Slovakia/UKBA-313/2020,Slovakia/UKBA-201/2020,Slovakia/UKBA-412/2020 | - |
| Belen Prado-Vivar et al | 1 | Ecuador/USFQ-004/2020 | - |
| Abdul Karim sesay et al | 21 | Gambia/NPHL-1209/2020,Gambia/NPHL-1208/2020,Gambia/NPHL-2727/2020,Gambia/NPHL-2759/2020,Gambia/NPHL-2892/2020,Gambia/NPHL-1248/2020,Gambia/NPHL-1230/2020,Gambia/NPHL-1446/2020,Gambia/GC19-3373/2020,Gambia/3735/2020,Gambia/NPHL-1224/2020,Gambia/NPHL-1645/2020,Gambia/NPHL-1711/2020,Gambia/NPHL-2615/2020,Gambia/GC19-0544/2020,Gambia/GC19-1387/2020,Gambia/GC19-1385/2020,Gambia/GC19-1706/2020,Gambia/NPHL-1216/2020,Gambia/GC19-1814/2020,Gambia/GC19-3266/2020 | - |
| Giandhari J et al | 31 | SouthAfrica/KRISP-0041/2020,SouthAfrica/KRISP-0131/2020,SouthAfrica/KRISP-0035/2020,SouthAfrica/KRISP-0040/2020,SouthAfrica/KRISP-0115/2020,SouthAfrica/KRISP-0067/2020,SouthAfrica/KRISP-0017/2020,SouthAfrica/KRISP-0055/2020,SouthAfrica/KRISP-0019/2020,SouthAfrica/KRISP-0160/2020,SouthAfrica/KRISP-0159/2020,SouthAfrica/KRISP-K000591/2020,SouthAfrica/KRISP-K002322/2020,SouthAfrica/KRISP-K002890/2020,SouthAfrica/KRISP-K003364/2020,SouthAfrica/KRISP-K003367/2020,SouthAfrica/KRISP-0385/2020,SouthAfrica/KRISP-K004641/2020,SouthAfrica/KRISP-K004638/2020,SouthAfrica/KRISP-0284/2020,SouthAfrica/KRISP-0285/2020,SouthAfrica/KRISP-0149/2020,SouthAfrica/KRISP-K002853/2020,SouthAfrica/KRISP-K004589/2020,SouthAfrica/KRISP-0564/2020,SouthAfrica/KRISP-K004631/2020,SouthAfrica/KRISP-K002393/2020,SouthAfrica/KRISP-K002765/2020,SouthAfrica/KRISP-K002755/2020,SouthAfrica/KRISP-K002757/2020,SouthAfrica/KRISP-K002825/2020 | - |
| Nabaes Jodar et al | 13 | Argentina/PAIS-A0004/2020,Argentina/PAIS-A0012/2020,Argentina/PAIS-A0007/2020,Argentina/PAIS-A0009/2020,Argentina/PAIS-A0013/2020,Argentina/PAIS-A0001/2020,Argentina/PAIS-A0026/2020,Argentina/PAIS-A0024/2020,Argentina/PAIS-A0023/2020,Argentina/PAIS-A0011/2020,Argentina/PAIS-A0014/2020,Argentina/PAIS-A0005/2020,Argentina/PAIS-A0016/2020 | COVID-19 in Latin America: Contrasting phylodynamic inference with epidemiological surveillance. |
| Marion Barbet et al | 7 | Morocco/6897/2020,Morocco/6889/2020,Morocco/6899/2020,Morocco/6887/2020,Morocco/6895/2020,Morocco/6905/2020,Morocco/6906/2020 | Genetic Diversity and Genomic Epidemiology of SARS-COV-2 in Morocco |

| | | | |
|---|---|---|---|
| Robert Belužić et al | 15 | Croatia/Zagreb-19/2020,Croatia/Zagreb-12/2020,Croatia/Zagreb-29/2020,Croatia/Zagreb-11/2020,Croatia/Zagreb-1/2020,Croatia/Zagreb-22/2020,Croatia/Zagreb-14/2020,Croatia/Zagreb-4/2020,Croatia/Zagreb-5/2020,Croatia/Zagreb-8/2020,Croatia/Zagreb-21/2020,Croatia/Zagreb-26/2020,Croatia/Zagreb-9/2020,Croatia/Zagreb-15/2020,Croatia/Zagreb-7/2020 | - |
| Salihefendic L. et al | 2 | BosniaandHerzegovina/AGC-MOS-002/2020,BosniaandHerzegovina/AGC-SAR-002/2020 | - |
| Leontina Banica et al | 11 | Romania/Suceava-5894/2020,Romania/Suceava-5816/2020,Romania/Bucuresti-4284/2020,Romania/Bucuresti-4105/2020,Romania/Bucuresti-6451/2020,Romania/Bucuresti-12546/2020,Romania/Mioveni-24095/2020,Romania/Bucuresti-13907/2020,Romania/Bucuresti-14251/2020,Romania/Bucuresti-15503/2020,Romania/Bucuresti-14545/2020 | - |
| Gvantsa Chanturia et al | 2 | Georgia/Tb-712/2020,Georgia/Tb-1679/2020 | - |
| Wbeimar Aguilar-Jimenez et al | 1 | Colombia/ANT-UdeA-200325-01/2020 | - |
| Juan José Guadalupe et al | 2 | Ecuador/USFQ-162/2020,Ecuador/USFQ-184/2020 | - |
| Echeverría C et al | 15 | Chile/AT-CMM-ATAC07/2020,Chile/AT-CMM-ATAC08/2020,Chile/AT-CMM-ATAC20/2020,Chile/AT-CMM-ATAC10/2020,Chile/AT-CMM-ATAC13/2020,Chile/AT-CMM-ATAC12/2020,Chile/AT-CMM-ATAC19/2020,Chile/AT-CMM-ATAC03/2020,Chile/AT-CMM-ATAC11/2020,Chile/AT-CMM-ATAC09/2020,Chile/AT-CMM-ATAC02/2020,Chile/AT-CMM-ATAC06/2020,Chile/AT-CMM-ATAC05/2020,Chile/AT-CMM-ATAC18/2020,Chile/AT-CMM-ATAC14/2020 | - |
| Carla López-Causapé et al | 3 | Spain/IB-IBV-99010784/2020,Spain/IB-IBV-99010811/2020,Spain/IB-IBV-99010812/2020 | - |
| Lobiuc Andrei et al | 1 | Romania/ROSV_12618/2020 | - |
| Derly Andrade et al | 4 | Ecuador/USFQ-133/2020,Ecuador/USFQ-129/2020,Ecuador/USFQ-207/2020,Ecuador/USFQ-528/2020 | - |
| Endre Gábor Tóth et al | 28 | Hungary/MH-789/2020,Hungary/MH-6633/2020,Hungary/MH-6603/2020,Hungary/MH-6657/2020,Hungary/MH-8268/2020,Hungary/MH-9653/2020,Hungary/MH-13211/2020,Hungary/UD-91623/2020,Hungary/UD-57955/2020,Hungary/UD-94400/2020,Hungary/UD-9953/2020,Hungary/UD-57964/2020,Hungary/US-60432w/2020,Hungary/UD-94347/2020,Hungary/US-57523w/2020,Hungary/MH-7051/2020,Hungary/UD-7039/2020,Hungary/UD-7417/2020,Hungary/UD-8157/2020,Hungary/US-482w/2020,Hungary/US-31273w/2020,Hungary/US-30412w/2020,Hungary/US-32618w/2020,Hungary/US-31885w/2020,Hungary/US-30242w/2020,Hungary/US-31248w/2020,Hungary/US-32311w/2020,Hungary/US-32315w/2020 | - |
| Andrea Macias et al | 4 | Ecuador/USFQ-213/2020,Ecuador/USFQ-523/2020,Ecuador/USFQ-249/2020,Ecuador/USFQ-524/2020 | - |
| Fabian Aguilar et al | 3 | Ecuador/USFQ-209/2020,Ecuador/USFQ-208/2020,Ecuador/USFQ-527/2020 | - |
| Nouar Qutob et al | 20 | Palestine/13/2020,Palestine/15/2020,Palestine/19/2020,Palestine/16/2020,Palestine/28/2020,Palestine/44/2020,Palestine/66/2020,Palestine/68/2020,Palestine/69/2020,Palestine/73/2020,Palestine/36/2020,Palestine/11/2020,Palestine/82/2020,Palestine/90/2020,Palestine/45/2020,Palestine/86/2020,Palestine/3/2020,Palestine/55/2020,Palestine/46/2020,Palestine/54/2020 | - |
| Rijad Konjhodzic et al | 1 | BosniaandHerzegovina/04-Sarajevo/2020 | Phylogenetic pattern of SARS-CoV-2 from COVID-19 patients from Bosnia and Herzegovina: lessons learned to optimize future molecular and epidemiological approaches |
| Baumeister E. et al | 2 | Argentina/C3013/2020,Argentina/C121/2020 | COVID-19 in Latin America: Contrasting phylodynamic inference with epidemiological surveillance. |

| | | | |
|---|---|---|---|
| Santos et al | 19 | Brazil/PA-IEC-166687/2020,Brazil/PB-IEC-161853/2020,Brazil/AP-IEC-165513/2020,Brazil/MA-IEC-166716/2020,Brazil/MA-IEC-164827/2020,Brazil/RN-IEC-162277/2020,Brazil/MA-IEC-162157/2020,Brazil/MA-IEC-165425/2020,Brazil/AP-IEC-162620/2020,Brazil/AP-IEC-165669/2020,Brazil/MA-IEC-165398/2020,Brazil/MA-IEC-162466/2020,Brazil/AP-IEC-163359/2020,Brazil/AP-IEC-164920/2020,Brazil/MA-IEC-166867/2020,Brazil/PA-IEC-165313/2020,Brazil/PA-IEC-164747/2020,Brazil/PA-IEC-165302/2020,Brazil/PA-IEC-163469/2020 | - |
| Luan Felipo Botelho-Souza et al | 8 | Brazil/RO-04/2020,Brazil/RO-06/2020,Brazil/RO-01/2020,Brazil/RO-03/2020,Brazil/RO-07/2020,Brazil/RO-08/2020,Brazil/RO-05/2020,Brazil/RO-02/2020 | - |
| Antonin Bal et al | 23 | France/ARA-43052/2020,France/ARA-33082/2020,France/ARA-SC370/2020,France/PAC-SC455/2020,France/ARA-SC005/2020,France/ARA-SC003/2020,France/ARA-107987/2020,France/ARA-111718/2020,France/ARA-SC135/2020,France/ARA-SC050/2020,France/ARA-SC054/2020,France/ARA-22132/2020,France/ARA-SC075/2020,France/PAC02004/2020,France/ARA-109965/2020,France/ARA97063/2020,France/OCC-SC531/2020,France/ARA-59557/2020,France/OCC-SC520/2020,France/ARA-SC179/2020,France/NAQ-isl2/2020,France/ARA-107339/2020,France/ARA-18437/2020 | - |
| Nick Vereecke et al | 3 | Belgium/UGent-122/2020,Belgium/UGent-302/2020,Belgium/UGent-225/2020 | - |
| Meriem LAAMARTI et al | 2 | Morocco/RMPS-15/2020,Morocco/RMPS-12/2020 | - |
| Keith Durkin et al | 31 | Belgium/ULG-11052/2020,Belgium/ULG-11055/2020,Belgium/ULG-11053/2020,Belgium/ULG-10177/2020,Belgium/ULG-10178/2020,Belgium/ULG-10299/2020,Belgium/ULG-10210/2020,Belgium/ULG-10247/2020,Belgium/ULG-10246/2020,Belgium/ULG-10175/2020,Belgium/ULG-10532/2020,Belgium/ULG-10258/2020,Belgium/ULG-10259/2020,Belgium/ULG-10254/2020,Belgium/ULG-10291/2020,Belgium/ULG-10951/2020,Belgium/ULG-10345/2020,Belgium/ULG-10348/2020,Belgium/ULG-10375/2020,Belgium/ULG-10374/2020,Belgium/ULG-10399/2020,Belgium/ULG-10966/2020,Belgium/ULG-10968/2020,Belgium/ULG-10999/2020,Belgium/ULG-11029/2020,Belgium/ULG-10654/2020,Belgium/ULG-10850/2020,Belgium/ULG-10716/2020,Belgium/ULG-10715/2020,Belgium/ULG-11039/2020,Belgium/ULG-10793/2020 | - |
| Senjuti Saha et al | 13 | Bangladesh/CHRF-0011/2020,Bangladesh/CHRF-0067/2020,Bangladesh/CHRF-0071/2020,Bangladesh/CHRF-0009/2020,Bangladesh/CHRF-0075/2020,Bangladesh/CHRF-0080/2020,Bangladesh/CHRF-0087/2020,Bangladesh/CHRF-0085/2020,Bangladesh/CHRF-0012/2020,Bangladesh/CHRF-0079/2020,Bangladesh/CHRF-0059/2020,Bangladesh/CHRF-0081/2020,Bangladesh/CHRF-0069/2020 | - |
| Junsong Zhang et al | 1 | Guangdong/SYSU-IHV/2020 | - |
| Antonio Piralla et al | 1 | Italy/LOM-INMI-5314-B/2020 | - |
| Adam Kotorashvili et al | 1 | Georgia/Tb/2020 | - |
| Fries et al | 2 | Cuba/USAFSAM-S030/2020,Cuba/USAFSAM-S031/2020 | - |
| Katherine Laiton-Donato et al | 4 | Colombia/ANT-INS-82457/2020,Colombia/VAC-INS-81279/2020,Colombia/VAC-INS-81251/2020,Colombia/SAN-INS-94514/2020 | Substitutions in Spike and Nucleocapsid proteins of SARS-CoV-2 circulating in Colombia |
| Gisela Barrera-Badillo et al | 10 | Mexico/BCS-InDRE-25/2020,Mexico/GRO-InDRE-24/2020,Mexico/TLA-InDRE-57/2020,Mexico/ZAC-InDRE-72/2020,Mexico/AGS-InDRE-82/2020,Mexico/AGS-InDRE-78/2020,Mexico/AGS-InDRE-80/2020,Mexico/COA-InDRE-36/2020,Mexico/COA-InDRE_40/2020,Mexico/NLE-InDRE-60/2020 | - |
| Sylvie Behillil et al | 8 | France/BRE-8930/2020,France/BRE-8931/2020,France/BRE-8932/2020,France/BRE-8943/2020,France/IDF-8679/2020,France/IDF-9137/2020,France/IDF-9230/2020,France/IDF-8702/2020 | - |
| David Navarro Ortega et al | 1 | Spain/VC-IBV-98007309/2020 | - |

| | | | |
|---|---|---|---|
| Gustavo Cilla et al | 5 | Spain/PV-IBV-98007662/2020,Spain/PV-IBV-98007637/2020,Spain/PV-IBV-2122/2020,Spain/PV-IBV-004225/2020,Spain/PV-IBV-004227/2020 | - |
| Georgi Merhi et al | 3 | Lebanon/LAU1-53249/2020,Lebanon/LAU3-53451/2020,Lebanon/LAU4-53460/2020 | - |
| SEARCH Alliance San Diego with Samuel Navarro Alvarez et al | 5 | Mexico/BCN-ALSR-1704/2020,Mexico/BCN-ALSR-1706/2020,Mexico/BCN-ALSR-1479/2020,Mexico/BCN-ALSR-1693/2020,Mexico/BCN-ALSR-1713/2020 | - |
| Luiza Ustea et al | 4 | Romania/Bucuresti-294285/2020,Romania/Buzau-291946/2020,Romania/Buzau-292711/2020,Romania/Buzau-293197/2020 | - |
| Andreas Henschel et al | 1 | UnitedArabEmirates/581/2020 | - |
| SEARCH Alliance San Diego with Idanya Rubí Serafín Higuera et al | 9 | Mexico/BCN-ALSR-2462/2020,Mexico/BCN-ALSR-2563/2020,Mexico/BCN-ALSR-2516/2020,Mexico/BCN-ALSR-2542/2020,Mexico/BCN-ALSR-2557/2020,Mexico/BCN-ALSR-2537/2020,Mexico/BCN-ALSR-2568/2020,Mexico/BCN-ALSR-2494/2020,Mexico/BCN-ALSR-2559/2020 | - |
| Ernesto Ramirez-Gonzalez et al | 4 | Mexico/QRO-InDRE-88/2020,Mexico/BCS-InDRE_30/2020,Mexico/QRO-InDRE-87/2020,Mexico/GRO-InDRE-93/2020 | - |
| Erin Young et al | 4 | USA/UT-UPHL-201202700/2020,USA/UT-UPHL-201230028/2020,USA/UT-UPHL-201204198/2020,USA/UT-UPHL-201216059/2020 | - |
| Claudio Tavares Sacchi et al | 2 | Brazil/SP-06/2020,Brazil/SP-05/2020 | Importation and early local transmission of COVID-19 in Brazil, 2020 |
| Vasiliki Pogka et al | 13 | Greece/127_HPI/2020,Greece/5884_HPI/2020,Greece/8540_HPI/2020,Greece/14999_HPI/2020,Greece/22565_HPI/2020,Greece/8334_HPI/2020,Greece/26072_HPI/2020,Greece/24440_HPI/2020,Greece/HPI-6465/2020,Greece/16034_HPI/2020,Greece/19553_HPI/2020,Greece/HPI-6478/2020,Greece/15005_HPI/2020 | - |
| Joan Marti-Carreras et al | 1 | Belgium/JK-0305110/2020 | A phylodynamic workflow to rapidly gain insights into the dispersal history and dynamics of SARS-CoV-2 lineages |
| Nagamani K et al | 1 | India/TG-GMC-KN318/2020 | Phylogenetic clustering of the Indian SARS-CoV-2 genomes reveals the presence of distinct clades of viral haplotypes among states |
| E Giombini et al | 2 | Italy/LAZ-INMI-96/2020,Italy/LAZ-INMI-106/2020 | - |
| M Rueca et al | 1 | Italy/LAZ-INMI-111/2020 | - |
| Department of Microbiology et al | 10 | Sweden/20-53631/2020,Sweden/20-09861/2020,Sweden/20-53570/2020,Sweden/20-09864/2020,Sweden/20-09865/2020,Sweden/20-53067/2020,Sweden/20-53532/2020,Sweden/20-21492/2020,Sweden/20-53635/2020,Sweden/20-53054/2020 | - |
| Spott et al | 4 | Germany/TH-IIMK-CaSe-18/2020,Germany/TH-IIMK-CaSe-17/2020,Germany/TH-IIMK-CaSe-16/2020,Germany/TH-IIMK-CaSe-7/2020 | - |
| B Bartolini et al | 1 | Italy/LAZ-INMI-108/2020 | - |
| F Messina et al | 1 | Italy/LAZ-INMI-112/2020 | - |
| Rafael Benito et al | 1 | Spain/AR-IBV-97009527/2020 | - |
| Parisi A. et al | 19 | Italy/APU-IZSPB-333PT/2020,Italy/APU-IZSPB_295PT/2020,Italy/APU-IZSPB_291PT/2020,Italy/APU-UniMI-5428/2020,Italy/APU-UniMI-5429/2020,Italy/APU-UniMI-5433/2020,Italy/APU-IZSPB-153APT/2020,Italy/APU-IZSPB-156APT/2020,Italy/APU-IZSPB_306PT/2020,Italy/APU-IZSPB-172APT/2020,Italy/APU-IZSPB-179APT/2020,Italy/APU-IZSPB-183PT/2020,Italy/APU-IZSPB-196PT/2020,Italy/APU-IZSPB-191APT/2020,Italy/BAS-IZSPB-339PT/2020,Italy/BAS-IZSPB-379PT/2020,Italy/APU-IZSPB_311PT/2020,Italy/APU-IZSPB_308PT/2020,Italy/APU-IZSPB_309PT/2020 | - |

| | | | |
|---|---|---|---|
| Antonio Rezusta López et al | 6 | Spain/AR-IBV-98010577/2020,Spain/AR-IBV-98010737/2020,Spain/AR-IBV-98010736/2020,Spain/AR-IBV-98005657/2020,Spain/AR-IBV-98005684/2020,Spain/AR-IBV-98005659/2020 | - |
| Elisa Martró et al | 1 | Spain/CT-IBV-97010161/2020 | - |
| Bastias M et al | 8 | Chile/RM-CMM-0000E/2020,Chile/RM-CMM-0184M/2020,Chile/RM-CMM-0263M/2020,Chile/RM-CMM-0166M/2020,Chile/RM-CMM-0069M/2020,Chile/RM-CMM-0096M/2020,Chile/RM-CMM-0257M/2020,Chile/RM-CMM-0007M/2020 | - |
| Oleg V. Pyankov et al | 2 | Russia/Samara-73406/2020,Russia/Buryatia-84508/2020 | - |
| Enatha Mukantwari et al | 1 | Rwanda/AF37991-RD/2020 | - |
| Gage Moreno et al | 7 | USA/WI-UW-1914/2020,USA/WI-UW-1916/2020,USA/WI-UW-2100/2020,USA/WI-UW-1921/2020,USA/WI-UW-1922/2020,USA/WI-UW-1291/2020,USA/WI-UW-1307/2020 | - |
| Hidayat Trimarsanto et al | 1 | Indonesia/SA-EIJK50/2020 | - |
| RCGEB - MASA et al | 21 | NorthMacedonia/6497/2020,NorthMacedonia/7238/2020,NorthMacedonia/7245/2020,NorthMacedonia/7813/2020,NorthMacedonia/9133/2020,NorthMacedonia/9360/2020,NorthMacedonia/2819/2020,NorthMacedonia/3101/2020,NorthMacedonia/2175/2020,NorthMacedonia/5220/2020,NorthMacedonia/11633/2020,NorthMacedonia/11602/2020,NorthMacedonia/9359/2020,NorthMacedonia/9304/2020,NorthMacedonia/883/2020,NorthMacedonia/12026/2020,NorthMacedonia/11567/2020,NorthMacedonia/9073/2020,NorthMacedonia/3122/2020,NorthMacedonia/11805/2020,NorthMacedonia/8000/2020 | - |
| Petr Brož et al | 4 | CzechRepublic/IAB_18/2020,CzechRepublic/IAB_21/2020,CzechRepublic/IAB_14/2020,CzechRepublic/IAB_17/2020 | - |
| Paul Cotter et al | 2 | Ireland/MH-NVRL-20G49748/2020,Ireland/MH-NVRL-20G49952/2020 | - |
| Dinesh Kumar et al | 1 | India/GJ-GBRC287/2020 | - |
| Arindam Maitra et al | 5 | India/WB-IK4/2020,India/WB-S13/2020,India/WB-IK18/2020,India/HR-NIBG-TF94/2020,India/MH-GA19/2020 | - |
| Danish Covid-19 Genome Consortia et al | 31 | Denmark/DCGC-1293/2020,Denmark/DCGC-1258/2020,Denmark/DCGC-1569/2020,Denmark/DCGC-3448/2020,Denmark/DCGC-2463/2020,Denmark/DCGC-7260/2020,Denmark/DCGC-7678/2020,Denmark/DCGC-7261/2020,Denmark/DCGC-2936/2020,Denmark/DCGC-5598/2020,Denmark/DCGC-5508/2020,Denmark/DCGC-5519/2020,Denmark/DCGC-2973/2020,Denmark/DCGC-5208/2020,Denmark/DCGC-7576/2020,Denmark/DCGC-1785/2020,Denmark/DCGC-2060/2020,Denmark/DCGC-5766/2020,Denmark/DCGC-2024/2020,Denmark/DCGC-2033/2020,Denmark/DCGC-2032/2020,Denmark/DCGC-1919/2020,Denmark/DCGC-3758/2020,Denmark/DCGC-3501/2020,Denmark/DCGC-1872/2020,Denmark/DCGC-873/2020,Denmark/DCGC-1254/2020,Denmark/DCGC-1864/2020,Denmark/DCGC-1213/2020,Denmark/DCGC-1195/2020,Denmark/DCGC-1488/2020 | - |
| Sharif Hala et al | 2 | SaudiArabia/KAUST-Jeddah61/2020,SaudiArabia/KAUST-Jeddah66/2020 | - |
| Ray Izquierdo-Lara et al | 1 | env/Netherlands/NH-92852-N1/2020 | - |
| Alberto Delgado-Iribarren et al | 1 | Spain/MD-IBV-97010011/2020 | - |
| Ahmad Abou Tayoun et al | 4 | UnitedArabEmirates/L2004034680/2020,UnitedArabEmirates/L920632696/2020,UnitedArabEmirates/L116596767/2020,UnitedArabEmirates/L2004034634/2020 | - |
| Kairavi Joshi et al | 1 | India/GJ-GBRC50/2020 | Phylogenetic clustering of the Indian SARS-CoV-2 genomes reveals the presence of distinct clades of viral haplotypes among states |

| | | | |
|---|---|---|---|
| CIDM-PH et al. et al | 12 | Australia/NSW527/2020,Australia/NSW518/2020,Australia/NSW529/2020,Australia/NSW673/2020,Australia/NSW294/2020,Australia/NSW332/2020,Australia/NSW606/2020,Australia/NSW617/2020,Australia/NSW1206/2020,Australia/NSW1202/2020,Australia/NSW1151/2020,Australia/NSW1217/2020 | - |
| Disha Patel et al | 1 | India/GJ-GBRC70/2020 | Phylogenetic clustering of the Indian SARS-CoV-2 genomes reveals the presence of distinct clades of viral haplotypes among states |
| Lex Leong et al | 3 | Australia/SAP212/2020,Australia/SAP550/2020,Australia/SAP551/2020 | - |
| Sunil Raghav et al | 2 | India/OR-ILSCV29215/2020,India/OR-ILSCV32593/2020 | - |
| Pramod Kumar# et al | 1 | India/DL-NCDC7715-CSIR-IGIB/2020 | - |
| Fathia Ben Rached et al | 2 | SaudiArabia/KAUST-JEDDAH1056/2020,SaudiArabia/KAUST-JEDDAH1057/2020 | - |
| Mahesh S. Dhar1* et al | 2 | India/DL-57013-CSIR-IGIB/2020,India/DL-57083-CSIR-IGIB/2020 | - |
| Souza et al | 11 | Brazil/RJ-DCV5/2020,Brazil/RJ-DCVN3/2020,Brazil/RJ-DCV05/2020,Brazil/RJ-DCV3/2020,Brazil/RJ-DCVTV507Q/2020,Brazil/RJ-DCV1/2020,Brazil/RJ-DCVN2/2020,Brazil/RJ-0920/2020,Brazil/RJ-01020R2/2020,Brazil/RJ-0720R2/2020,Brazil/RJ-0720R1/2020 | - |
| Oluniyi P.E. et al | 1 | Nigeria/Lagos01/2020 | First African SARS-CoV-2 genome sequence from Nigerian COVID-19 case |
| Chu et al et al | 3 | USA/WA-S1163/2020,USA/WA-S1179/2020,USA/WA-S373/2020 | - |
| Simulundu et al | 1 | Zambia/29/2020 | - |
| Sachin Chalapati et al | 2 | Ireland/KY-NVRL-20G35628/2020,Ireland/D-NVRL-20G35041/2020 | - |
| Sanaâ Lemriss et al | 2 | Morocco/moh100/2020,Morocco/AZ183/2020 | - |
| Caly et al | 2 | Australia/VIC1387/2020,Australia/VIC10431/2020 | - |
| Kelsey R. Florek et al | 1 | USA/WI-WSLH-200084/2020 | - |
| Ereqat et al | 1 | Palestine/WB-AQU-P3/2020 | - |
| Nasereddin et al | 1 | Palestine/WB-AQU-1/2020 | - |
| Ramzi Fattouh et al | 18 | Canada/ON-UHTC_0149/2020,Canada/ON-UHTC-0298/2020,Canada/ON-UHTC_0122/2020,Canada/ON-UHTC_0345/2020,Canada/ON-UHTC_0340/2020,Canada/ON-UHTC-0366/2020,Canada/ON-UHTC-0373/2020,Canada/ON-UHTC-0363/2020,Canada/ON-UHTC-0376/2020,Canada/ON-UHTC-0377/2020,Canada/ON-UHTC_0381/2020,Canada/ON-UHTC-0258/2020,Canada/ON-UHTC-0222/2020,Canada/ON-UHTC-0232/2020,Canada/ON-UHTC-0287/2020,Canada/ON-UHTC_0318/2020,Canada/ON-UHTC-0288/2020,Canada/ON-UHTC-0188/2020 | - |
| Khan et al | 4 | Bangladesh/DNAS-BSH-isl-9/2020,Bangladesh/DNAS-RAB-isl-114/2020,Bangladesh/DNAS-RAB-isl-82/2020,Bangladesh/DNAS-RAB-isl-86/2020 | - |
| Dahdouh et al | 1 | Spain/MD-H12-LP28-2522/2020 | - |
| Ulinici M et al | 3 | Moldova/ICGEB_MD7/2020,Moldova/ICGEB_MD6/2020,Moldova/ICGEB_MD1/2020 | - |
| Franceschi et al. | 21 | Brazil/RS-25833,Brazil/RS-13979,Brazil/RS-22838,Brazil/RS-11569,Brazil/RS-26977,Brazil/RS-24277,Brazil/RS-11069,Brazil/RS-13368,Brazil/RS-24285,Brazil/RS-31799,Brazil/RS-9881,Brazil/RS-11262,Brazil/RS-11574,Brazil/RS-19337,Brazil/RS-31784,Brazil/RS-31800,Brazil/RS-15371,Brazil/RS-31786,Brazil/RS-24565,Brazil/RS-27623,Brazil/RS-13367 | - |

| | | | |
|---|---|---|---|
| Artem Fadeev et al | 13 | Russia/OMS-ORINFI-5912S/2020,Russia/OMS-ORINFI-1157S/2020,Russia/OMS-ORINFI-2166S/2020,Russia/OMS-ORINFI-864S/2020,Russia/OMS-ORINFI-2128S/2020,Russia/OMS-ORINFI-1141S/2020,Russia/OMS-ORINFI-120S/2020,Russia/OMS-ORINFI-60S/2020,Russia/OMS-ORINFI-78S/2020,Russia/OMS-ORINFI-8296S/2020,Russia/OMS-ORINFI-56S-2/2020,Russia/OMS-ORINFI-4832S/2020,Russia/OMS-ORINFI-3970S/2020 | - |
| Tulasi Nagabandi et al | 1 | India/TG-CCMB-M5/2020 | - |
| Namami Gaur et al | 1 | India/TG-CCMB-X103/2020 | - |
| Nikhil Hajirnis et al | 1 | India/TG-CCMB-X115/2020 | - |
| Potdar V et al | 2 | India/MH-NIV-SARI-4375/2020,India/MH-NIV-45071/2020 | - |
| Tujan et al | 1 | Philippines/PH-RITM-0019/2020 | - |
| Jouali Farah et al | 1 | Morocco/15N/2020 | SARS-CoV-2 Genome Sequence from Morocco, Obtained Using Ion AmpliSeq Technology |
| Vinayasekhar Aedula et al | 1 | India/TG-CCMB-GC20/2020 | - |
| Lamuk Zaveri et al | 2 | India/TG-CCMB-AC338/2020,India/TG-CCMB-X116/2020 | - |
| Onur TOKGUN et al. et al | 10 | Turkey/DEN-PAU01/2020,Turkey/DEN-PAU07/2020,Turkey/DEN-PAU19/2020,Turkey/DEN-PAU12/2020,Turkey/DEN-PAU14/2020,Turkey/DEN-PAU08/2020,Turkey/DEN-PAU15/2020,Turkey/DEN-PAU03/2020,Turkey/DEN-PAU05/2020,Turkey/DEN-PAU13/2020 | - |
| Kenjiro Kosaki et al | 14 | Japan/Donner30/2020,Japan/Donner34/2020,Japan/Donner33/2020,Japan/Donner37/2020,Japan/Donner41/2020,Japan/Donner42/2020,Japan/Donner49/2020,Japan/Donner43/2020,Japan/Donner86/2020,Japan/Donner93/2020,Japan/Donner84/2020,Japan/Donner66/2020,Japan/Donner68/2020,Japan/Donner78/2020 | - |
| Yaqub et al | 5 | Pakistan/UN-UVAS-Lahore-IV/2020,Pakistan/UN-UVAS-Lahore-III/2020,Pakistan/UN-UVAS-Lahore-I/2020,Pakistan/UN-UVAS-Lahore-II/2020,Pakistan/UN-UVAS-Sialkot/2020 | - |
| Jorge Gonzalez et al | 7 | Chile/MA-UMAG-Catg-7/2020,Chile/MA-UMAG-Catg-5/2020,Chile/MA-UMAG-Catg-21/2020,Chile/MA-UMAG-Catg-6/2020,Chile/MA-UMAG-Catg-20/2020,Chile/MA-UMAG-Catg-5-2/2020,Chile/MA-UMAG-Catg-4/2020 | - |
| Maurizio Viscardi et al | 5 | Italy/CAM-IZSM-183/2020,Italy/CAM-IZSM-166/2020,Italy/CAM-IZSM-172/2020,Italy/CAM-IZSM-112/2020,Italy/CAM-IZSM-129/2020 | - |
| Gray K et al | 1 | Australia/NSW165/2020 | Revealing COVID-19 transmission in Australia by SARS-CoV-2 genome sequencing and agent-based modeling |
| Fernando L Melo et al | 1 | Brazil/DF-0001/2020 | Evolution and epidemic spread of SARS-CoV-2 in Brazil |
| Santos et al | 14 | Brazil/AC-IEC-162535/2020,Brazil/MA-IEC-163069/2020,Brazil/AP-IEC-162966/2020,Brazil/PA-IEC-164684/2020,Brazil/AP-IEC-162741/2020,Brazil/AP-IEC-164346/2020,Brazil/AP-IEC-161167/2020,Brazil/PA-IEC-161548/2020,Brazil/AP-IEC-164082/2020,Brazil/AP-IEC-163972/2020,Brazil/PA-IEC-164218/2020,Brazil/PA-IEC-164239/2020,Brazil/PA-IEC-162802/2020,Brazil/PA-IEC-164173/2020 | Genomic surveillance of SARS-CoV-2 reveals community transmission of a major lineage during the early pandemic phase in Brazil |
| Gustavo D. P. Silva et al | 2 | Brazil/RJ-MD63-1a/2020,Brazil/RJ-MD63-1b/2020 | - |
| Jolene Bowers et al | 1 | USA/AZ-TG569537/2020 | - |

| | | | |
|---|---|---|---|
| Carolina M Voloch et al | 110 | Brazil/RJ-00451/2020,Brazil/RJ-00520/2020,Brazil/RJ-00461/2020,Brazil/RJ-00450/2020,Brazil/RJ-00470/2020,Brazil/RJ-00532/2020,Brazil/RJ-00475/2020,Brazil/RJ-00474/2020,Brazil/MG-00302/2020,Brazil/RJ-00316/2020,Brazil/RJ-00355/2020,Brazil/RJ-00525/2020,Brazil/RJ-00456/2020,Brazil/RJ-00438/2020,Brazil/RJ-00505/2020,Brazil/MG-00301/2020,Brazil/RJ-00463/2020,Brazil/RJ-00471/2020,Brazil/RJ-00476/2020,Brazil/RJ-00477/2020,Brazil/RJ-00466/2020,Brazil/RJ-00468/2020,Brazil/RJ-00457/2020,Brazil/RJ-00338/2020,Brazil/RJ-00517/2020,Brazil/RJ-00452/2020,Brazil/RJ-00453/2020,Brazil/RJ-00455/2020,Brazil/RJ-00460/2020,Brazil/RJ-00575/2020,Brazil/RJ-00534/2020,Brazil/RJ-00511/2020,Brazil/RJ-00521/2020,Brazil/RJ-00478/2020,Brazil/RJ-00454/2020,Brazil/RJ-00348/2020,Brazil/RJ-00362/2020,Brazil/RJ-00522/2020,Brazil/RJ-00358/2020,Brazil/RJ-00443/2020,Brazil/RJ-00337/2020,Brazil/RJ-00340/2020,Brazil/RJ-00469/2020,Brazil/RJ-00472/2020,Brazil/RJ-00473/2020,Brazil/RJ-00467/2020,Brazil/RJ-00526/2020,Brazil/RJ-00441/2020,Brazil/RJ-00305/2020,Brazil/RJ-00339/2020,Brazil/RJ-00345/2020,Brazil/RJ-00524/2020,Brazil/RJ-00439/2020,Brazil/RJ-00311/2020,Brazil/RJ-00359/2020,Brazil/RJ-00346/2020,Brazil/RJ-00509/2020,Brazil/RJ-00504/2020,Brazil/RJ-00429/2020,Brazil/RJ-00413/2020,Brazil/RJ-00412/2020,Brazil/RJ-00327/2020,Brazil/RJ-00328/2020,Brazil/RJ-00544/2020,Brazil/RJ-00546/2020,Brazil/RJ-00540/2020,Brazil/RJ-00560/2020,Brazil/RJ-00561/2020,Brazil/RJ-00562/2020,Brazil/RJ-00555/2020,Brazil/RJ-00570/2020,Brazil/RJ-00553/2020,Brazil/RJ-00563/2020,Brazil/RJ-00572/2020,Brazil/RJ-00571/2020,Brazil/RJ-00569/2020,Brazil/RJ-00564/2020,Brazil/RJ-00559/2020,Brazil/RJ-00539/2020,Brazil/RJ-00545/2020,Brazil/RJ-00550/2020,Brazil/RJ-00557/2020,Brazil/RJ-00541/2020,Brazil/RJ-00537/2020,Brazil/RJ-00542/2020,Brazil/RJ-00551/2020,Brazil/RJ-00536/2020,Brazil/RJ-00547/2020,Brazil/RJ-00558/2020,Brazil/RJ-00565/2020,Brazil/RJ-00554/2020,Brazil/RJ-00566/2020,Brazil/RJ-00573/2020,Brazil/RJ-00543/2020,Brazil/RJ-00549/2020,Brazil/RJ-00548/2020,Brazil/RJ-00556/2020,Brazil/RJ-00567/2020,Brazil/RJ-00568/2020,Brazil/RJ-00552/2020,Brazil/RJ-00405/2020,Brazil/RJ-00410/2020,Brazil/RJ-00430/2020,Brazil/RJ-00411/2020,Brazil/RJ-00402/2020,Brazil/RJ-00407/2020,Brazil/RJ-00406/2020,Brazil/RJ-00414/2020,Brazil/RJ-00415/2020,Brazil/RJ-00428/2020 | - |
| Amgarteb et al | 1 | Brazil/UN-HIAE-SP03/2020 | - |
| Prameet M. Sheth et al | 2 | Canada/ON-OICR-SLB2820/2020,Canada/ON-OICR-SLB2821/2020 | - |
| Bianca Catarina Azevedo Cabral et al | 3 | Brazil/RJ-UFRJ-58271/2020,Brazil/RJ-UFRJ-32317/2020,Brazil/RJ-UFRJ-9331/2020 | - |
| Gaggero A et al | 7 | Chile/RM-CMM-LRF3924/2020,Chile/RM-CMM-ADC1858/2020,Chile/RM-CMM-NA4204/2020,Chile/RM-CMM-EDB244K/2020,Chile/RM-MSHS-RSV7306/2020,Chile/RM-CMM-MCD9529/2020,Chile/RM-CMM-LAE7918/2020 | - |
| Juliana D. Siqueira et al | 17 | Brazil/RJ-INCA-I34/2020,Brazil/RJ-INCA-I57/2020,Brazil/RJ-INCA-I51/2020,Brazil/RJ-INCA-I30/2020,Brazil/RJ-INCA-I35/2020,Brazil/RJ-INCA-C41/2020,Brazil/RJ-INCA-C33/2020,Brazil/RJ-INCA-C22/2020,Brazil/RJ-INCA-C63/2020,Brazil/RJ-INCA-C68/2020,Brazil/RJ-INCA-C69/2020,Brazil/RJ-INCA-C77/2020,Brazil/RJ-INCA-I43/2020,Brazil/RJ-INCA-I39/2020,Brazil/RJ-INCA-C07/2020,Brazil/RJ-INCA-C24/2020,Brazil/RJ-INCA-C64/2020 | - |
| Sarah Schmedes et al | 7 | USA/FL-BPHL-0618/2020,USA/FL-BPHL-0820/2020,USA/FL-BPHL-0829/2020,USA/FL-BPHL-0863/2020,USA/FL-BPHL-0869/2020,USA/FL-BPHL-1793/2020,USA/FL-BPHL-0961/2020 | - |
| Virginia DCLS et al | 6 | USA/VA-DCLS-0584/2020,USA/VA-DCLS-2187/2020,USA/VA-DCLS-2185/2020,USA/VA-DCLS-2191/2020,USA/VA-DCLS-1775/2020,USA/VA-DCLS-1935/2020 | - |
| Svetoslav N Slavov et al | 1 | Brazil/SP-RP4/2020 | - |
| Samples: Ingra Morales Claro et al | 6 | Brazil/SP-L9-CD139/2020,Brazil/SP-L8-CD101/2020,Brazil/SC-L15-CD265/2020,Brazil/SP-L14-CD240/2020,Brazil/SP-L22-CD474/2020,Brazil/SP-L9-CD90/2020 | Evolution and epidemic spread of SARS-CoV-2 in Brazil |

| | | | |
|---|---|---|---|
| Claudio Tavares Sacchi et al C | 74 | Brazil/SP-625/2020,Brazil/SP-341/2020,Brazil/SP-384/2020,Brazil/SP-624/2020,Brazil/SP-390/2020,Brazil/SP-551/2020,Brazil/SP-122/2020,Brazil/SP-124/2020,Brazil/SP-388/2020,Brazil/SP-300/2020,Brazil/SP-225/2020,Brazil/SP-844R1/2020,Brazil/SP-163/2020,Brazil/SP-600/2020,Brazil/SP-427/2020,Brazil/SP-256/2020,Brazil/SP-692/2020,Brazil/SP-438/2020,Brazil/SP-666/2020,Brazil/SP-429/2020,Brazil/SP-577/2020,Brazil/SP-694/2020,Brazil/SP-719/2020,Brazil/SP-415/2020,Brazil/SP-249/2020,Brazil/SP-696/2020,Brazil/SP-317/2020,Brazil/SP-668/2020,Brazil/SP-324/2020,Brazil/SP-591/2020,Brazil/SP-356/2020,Brazil/SP-321/2020,Brazil/SP-688/2020,Brazil/SP-400/2020,Brazil/SP-437/2020,Brazil/SP-433/2020,Brazil/SP-436/2020,Brazil/SP-401/2020,Brazil/SP-389/2020,Brazil/SP-639/2020,Brazil/SP-537/2020,Brazil/SP-272/2020,Brazil/SP-370/2020,Brazil/SP-590/2020,Brazil/SP-673/2020,Brazil/SP-361/2020,Brazil/SP-844R2/2020,Brazil/SP-149/2020,Brazil/SP-147/2020,Brazil/SP-125/2020,Brazil/SP-154/2020,Brazil/SP-190/2020,Brazil/SP-294/2020,Brazil/SP-253/2020,Brazil/SP-461/2020,Brazil/SP-283/2020,Brazil/SP-459/2020,Brazil/SP-209/2020,Brazil/SP-516/2020,Brazil/SP-203/2020,Brazil/SP-243/2020,Brazil/SP-439/2020,Brazil/SP-255/2020,Brazil/SP-417/2020,Brazil/SP-383/2020,Brazil/SP-394/2020,Brazil/SP-398/2020,Brazil/SP-440/2020,Brazil/SP-376/2020,Brazil/SP-589/2020,Brazil/SP-353/2020,Brazil/SP-393/2020,Brazil/SP-579/2020,Brazil/SP-425/2020 | - |
| Valiente F et al | 4 | Chile/RM-CMM-IN533883977/2020,Chile/RM-CMM-A5P533884002/2020,Chile/RM-CMM-IN233883987/2020,Chile/RM-CMM-A2P533884024/2020 | - |
| José Javier Costa Alcalde et al | 2 | Spain/GA-IBV-98006031/2020,Spain/GA-IBV-98006079/2020 | - |
| Roberta Crespo et al | 1 | Argentina/Heritas_HG006/2020 | - |
| Andrea Mangano et al | 1 | Argentina/Heritas_HG007/2020 | - |
| Pavitra Roychoudhury et al | 1 | USA/WA-UW-4015/2020 | - |
| Son Nguyen et al. et al | 1 | Australia/QLD1196/2020 | - |
| Suzukawa et al | 1 | Brazil/PR-LRV-01/2020 | - |
| Alfredo Bruno Caicedo et al | 9 | Ecuador/36230/2020,Ecuador/36226/2020,Ecuador/51908/2020,Ecuador/30755/2020,Ecuador/52389/2020,Ecuador/52872/2020,Ecuador/36192/2020,Ecuador/23055/2020,Ecuador/23890/2020 | - |
| Belen Prado-Vivar et al | 1 | Ecuador/HGSQ-USFQ-018/2020 | Genome sequencing of the first SARS-CoV-2 reported from patients with COVID-19 in Ecuador. |
| Rosario Moreno et al | 1 | Spain/VC-IBV-98003572/2020 | - |
| Andrey Komissarov et al | 10 | Russia/SPE-RII-MH1628S/2020,Russia/SVE-RII-MH1640S/2020,Russia/IN-RII-MH2745S/2020,Russia/SVE-RII-MH231S/2020,Russia/ORL-RII-MH2815S/2020,Russia/StPetersburg-RII11250S/2020,Russia/StPetersburg-RII11151S/2020,Russia/SPE-RII-MH971S/2020,Russia/NVS-RII-MH1137S/2020,Russia/StPetersburg-RII21769S/2020 | - |
| Martin Smith et al | 2 | Canada/QC-AL5/2020,Canada/QC_AY8/2020 | - |
| The Lighthouse Lab in Alderley Park et al | 8 | England/ALDP-6BCFB5/2020,England/ALDP-49876E/2020,England/ALDP-49E391/2020,England/ALDP-92578F/2020,England/ALDP-68E0E6/2020,England/ALDP-5B7935/2020,England/ALDP-92841C/2020,England/ALDP-5B5EFC/2020 | - |
| Sam Haldenby et al | 5 | England/LIVE-A254D/2020,England/LIVE-A02FD/2020,England/LIVE-AA3F0/2020,England/LIVE-DC4751/2020,England/ALDP-98C391/2020 | - |
| Abu Sayeed Mohammad Mahmud et al | 4 | Bangladesh/BCSIR-NILMRC-305/2020,Bangladesh/BCSIR-NILMRC-311/2020,Bangladesh/BCSIR-NILMRC-005/2020,Bangladesh/BCSIR-NILMRC-076/2020 | - |
| Marek Sanak et al | 8 | Poland/PL_P23/2020,Poland/PL_P12/2020,Poland/PL_P24/2020,Poland/PL_P21/2020,Poland/PL_P10/2020,Poland/PL_P14/2020,Poland/PL_P19/2020,Poland/PL_P20/2020 | - |
| Katarzyna Pancer et al | 2 | Poland/PL_P35/2020,Poland/PL_P28/2020 | - |

| | | | |
|---|---|---|---|
| Kristína Boršová et al | 4 | Slovakia/UKBA-208/2020,Slovakia/UKBA-203/2020,Slovakia/UKBA-205/2020,Slovakia/UKBA-318/2020 | - |
| Maximilian Damagnez et al | 10 | Germany/NW-HHU-298/2020,Germany/NW-HHU-D871/2020,Germany/NW-HHU-348/2020,Germany/NW-HHU-273/2020,Germany/NW-HHU-271/2020,Germany/NW-HHU-105/2020,Germany/NW-HHU-101/2020,Germany/NW-HHU-104/2020,Germany/NW-HHU-289/2020,Germany/NW-HHU-315/2020 | - |
| Joseph Fauver et al | 1 | USA/CT-Yale-453/2020 | - |
| WILSON JOSE DA SILVA JUNIOR et al | 2 | Brazil/PE-COV0260/2020,Brazil/PE-AMU0036/2020 | - |
| Rob Howes et al | 4 | England/CAMC-C46B6B/2020,England/CAMC-A81CBE/2020,England/CAMC-C3E586/2020,England/CAMC-A3F147/2020 | - |
| Harper VanSteenhouse et al | 9 | England/QEUH-B806FC/2020,England/QEUH-768C1A/2020,England/QEUH-960524/2020,England/QEUH-962D66/2020,Scotland/QEUH-96F9FC/2020,England/QEUH-96FA62/2020,England/QEUH-96FA17/2020,England/QEUH-B2E942/2020,England/QEUH-BA320E/2020 | - |
| Jade Wang et al | 1 | USA/NY-NYCPHL-001334/2020 | - |
| José Luiz Proença-Modena et al | 1 | Brazil/SP-L5-CAMPI83/2020 | Evolution and epidemic spread of SARS-CoV-2 in Brazil |
| Stefan Schmutz et al | 1 | Switzerland/ZH-UZH-1000477796/2020 | A doubt of multiple introduction of SARS-CoV-2 in Italy: A preliminary overview |
| Joan Marti-Carerras et al | 1 | Belgium/DHWM-03041/2020 | A phylodynamic workflow to rapidly gain insights into the dispersal history and dynamics of SARS-CoV-2 lineages |
| Nguyen Thi Tam et al | 6 | Vietnam/VNHN_022/2020,Vietnam/VNHN_0299/2020,Vietnam/VNHN_4868/2020,Vietnam/VNHN_4189/2020,Vietnam/VNHN_0419/2020,Vietnam/VNHN_4851/2020 | - |
| Griselda De Marco et al | 1 | Spain/VC-FISABIO-132/2020 | - |
| Lenka Kramna et al | 1 | CzechRepublic/Seq7/2020 | - |
| Tsuyoshi Sekizuka et al E | 5 | Japan/P4-5/2020,Japan/P4-3/2020,Japan/P4-1/2020,Japan/P5-1/2020,Japan/P4-2/2020 | SARS-CoV-2 Genome Analysis of Japanese Travelers in Nile River Cruise |
| Sesay et al et al | 8 | Gambia/0268/2020,Gambia/0548/2020,Gambia/0214/2020,Gambia/0278/2020,Gambia/0547/2020,Gambia/0536/2020,Gambia/0528/2020,Gambia/0531/2020 | - |
| Souad KARTTI et al | 1 | Morocco/RMPS-22/2020 | - |
| Alla Mironenko et al | 8 | Ukraine/Ivano-Frankivsk-706/2020,Ukraine/Kharkiv-877/2020,Ukraine/Kharkiv-782/2020,Ukraine/Kharkiv-705/2020,Ukraine/Kyiv-785/2020,Ukraine/Chernivtsi_861/2020,Ukraine/Lutsk-889/2020,Ukraine/Kharkiv-882/2020 | - |
| Samoilov AE et al | 8 | Russia/MOS-CRIE-13385510/2020,Russia/MOS-CRIE-13455107/2020,Russia/MOS-CRIE-13449451/2020,Russia/MOS-CRIE7157442/2020,Russia/MOS-CRIE7165818/2020,Russia/MOS-CRIE7162018/2020,Russia/MOS-CRIE-13297130/2020,Russia/MOS-CRIE-13379339/2020 | - |
| Matt Storey et al | 5 | NewZealand/20VR0712/2020,NewZealand/20VR1133/2020,NewZealand/20VR2062/2020,NewZealand/20VR2541/2020,NewZealand/20VR2659/2020 | - |
| Alghoribi et al | 1 | SaudiArabia/KAIMRC03/2020 | - |
| Jacquelyn Wynn et al | 3 | Wales/ALDP-C06842/2020,England/ALDP-A51D87/2020,England/ALDP-B5B6FA/2020 | - |
| Md. Moniruzzaman et al | 1 | Bangladesh/NIB-01/2020 | Coding-Complete Genome Sequence of SARS-CoV-2 Isolate from Bangladesh by Sanger Sequencing |
| Seemann T. et al | 8 | Australia/VIC5995/2020,Australia/VIC3877/2020,Australia/VIC4011/2020,Australia/VIC14300/2020,Australia/VIC16787/2020,Australia/VIC15897/2020,Australia/VIC14684/2020,Australia/VIC15878/2020 | - |

| | | | |
|---|---|---|---|
| Barbara Bartolini et al | 2 | Italy/LAZ-INMI-29/2020,Italy/LAZ-INMI-66/2020 | - |
| Iffat Jahan et al | 3 | Bangladesh/BCSIR-NILMRC_423/2020,Bangladesh/BCSIR-NILMRC-100/2020,Bangladesh/BCSIR-NILMRC_422/2020 | - |
| Barna Goswami et al | 2 | Bangladesh/BCSIR-NILMRC_420/2020,Bangladesh/BCSIR-NILMRC_421/2020 | - |
| Tanjina Akhter Banu et al | 2 | Bangladesh/BCSIR-NILMRC_397/2020,Bangladesh/BCSIR-NILMRC-223/2020 | - |
| Md. Murshed Hasan Sarkar et al | 1 | Bangladesh/BCSIR-NILMRC-103/2020 | - |
| Shahina Akter et al | 2 | Bangladesh/BCSIR-NILMRC-226/2020,Bangladesh/BCSIR-NILMRC-165/2020 | - |
| Salceanu et al | 1 | Romania/BUCHAREST-IMT-1/2020 | - |
| Justinas Slikas et al | 6 | Lithuania/C20-05-R7/2020,Lithuania/C20-05-R5/2020,Lithuania/C20-05-R11/2020,Lithuania/C20-05-R19/2020,Lithuania/C20-06-R20/2020,Lithuania/C20-05-R15/2020 | - |
| Janine Michel et al | 1 | Germany/BE-RKI-Z-0023/2020 | - |
| Rahul P Salunke et al | 3 | SaudiArabia/KAUST-JEDDAH927/2020,SaudiArabia/KAUST-JEDDAH877/2020,SaudiArabia/KAUST-JEDDAH886/2020 | - |
| Shehab et al | 1 | Bahrain/920268866/2020 | - |
| Goletic et al | 1 | BosniaandHerzegovina/01-Livno/2020 | Phylogenetic pattern of SARS-CoV-2 from COVID-19 patients from Bosnia and Herzegovina: lessons learned to optimize future molecular and epidemiological approaches |
| Raeece Naeem et al | 2 | SaudiArabia/KAUST-JEDDAH999/2020,SaudiArabia/KAUST-JEDDAH1020/2020 | - |
| Hematopathology Laboratory et al | 1 | India/MH-ACTREC-521/2020 | - |
| Yoshihiro Nakata et al | 12 | Japan/NGY-NNH-034/2020,Japan/NGY-NNH-054/2020,Japan/NGY-NNH-050/2020,Japan/NGY-NNH-051/2020,Japan/NGY-NNH-055/2020,Japan/NGY-NNH-047/2020,Japan/NGY-NNH-028/2020,Japan/NGY-NNH-049/2020,Japan/NGY-NNH-052/2020,Japan/NGY-NNH-031/2020,Japan/NGY-NNH-009/2020,Japan/NGY-NNH-053/2020 | - |
| Alexandra Lorentz et al | 1 | USA/MN-MDH-2187/2020 | - |
| Johan Ringlander et al | 1 | Sweden/20-11321/2020 | - |
| Samiha Al-Kharusi et al | 3 | Oman/24647/2020,Oman/C-7102/2020,Oman/C-5174/2020 | - |
| Chun Hang AU et al | 1 | HongKong/HKSH0007/2020 | Genome Sequences of SARS-CoV-2 Strains Detected in Hong Kong |
| Szymon Hryhorowicz et al | 2 | Poland/IHG-PAS-8-29/2020,Poland/IHG-PAS-8-30/2020 | - |
| Ramirez-Gonzalez Ernesto et al | 1 | Mexico/CMX-InDRE-07/2020 | - |
| Stern Lab et al | 1 | Israel/2116859/2020 | Full genome viral sequences inform patterns of SARS-CoV-2 spread into and within Israel |
| Filipe Romero et al | 1 | Brazil/MG-0101/2020 | - |
| Stephanie Hutchings et al | 1 | England/BRIS-124E38/2020 | Preliminary analysis of SARS-CoV-2 importation & establishment of UK transmission lineages |
| Ozer et al | 1 | Turkey/KOC-IST-OD5/2020 | - |
| Alexey Shchetinin et al | 2 | Russia/Moscow-PMVL-12/2020,Russia/Moscow_PMVL-2/2020 | - |
| Oliver Lung et al | 4 | mink/Canada/A/2020,mink/Canada/B/2020,mink/Canada/D/2020,mink/Canada/C/2020 | - |
| Pence et al | 4 | Turkey/IMU-SP-03/2020,Turkey/IMU-SP-04/2020,Turkey/IMU-SP-02/2020,Turkey/IMU-SP-01/2020 | - |

| | | | |
|---|---|---|---|
| Dovrolis N. et al | 1 | cat/Greece/2K/2020 | - |
| Xiang Zhao et al | 6 | Liaoning/IVDC-02/2020,Liaoning/IVDC-01/2020,Liaoning/IVDC-04/2020,Liaoning/IVDC-03/2020,Beijing/IVDC-02-06/2020,env/Beijing/IVDC-03-06/2020 | - |
| Leandro Patino Patino et al | 3 | Ecuador/2343/2020,Ecuador/2479/2020,Ecuador/2512/2020 | - |
| Ramirez-Gonzalez Ernesto et al | 1 | Mexico/CMX-InDRE-01/2020 | Full genome sequence of the first SARS-CoV-2 detected in Mexico |
| Neris Garcia-Gonzalez et al | 1 | Spain/VC-FISABIO-25/2020 | - |
| Silvia Fillo et al | 2 | Italy/LAZ-AMC3-5015/2020,Italy/SAR-ATS-AMC1-4634/2020 | - |
| Calm Walsh et al | 1 | Ireland/CE-NVRL-MPW96382/2020 | - |
| Chaoran Chen et al | 7 | Switzerland/ZH-ETHZ-400173/2020,Switzerland/SO-ETHZ-400044/2020,Switzerland/SO-ETHZ-400040/2020,Switzerland/AG-ETHZ-360224/2020,Switzerland/AG-ETHZ-400138/2020,Switzerland/ZH-ETHZ-400158/2020,Switzerland/BE-ETHZ-360199/2020 | - |
| LIC et al | 1 | Latvia/test1/2020 | - |
| Thrilok Chander B et al | 1 | India/TG-GMC-TC469/2020 | Phylogenetic clustering of the Indian SARS-CoV-2 genomes reveals the presence of distinct clades of viral haplotypes among states |
| Péter Urbán et al | 4 | Hungary/SRC-00105w/2020,Hungary/SRC-00278w/2020,Hungary/SRC-02801w/2020,Hungary/SRC-03670w/2020 | - |
| Azhar et al | 1 | SaudiArabia/86327/2020 | - |
| Afrah Alsomali et al | 3 | SaudiArabia/KAUST-JEDDAH840/2020,SaudiArabia/KAUST-JEDDAH867/2020,SaudiArabia/KAUST-MADINAH859/2020 | - |
| Rajesh Pandey# et al | 2 | India/DL-MaxCov0014-CSIR-IGIB/2020,India/DL-MaxCov0022-CSIR-IGIB/2020 | - |
| Andrey Komissarov et al | 3 | Russia/StPetersburg-RII4328S/2020,Russia/StPetersburg-RII6065S/2020,Russia/StPetersburg-RII7039V/2020 | Genomic epidemiology of the early stages of SARS-CoV-2 outbreak in Russia |
| Fatma Bayrakdar et al | 2 | Turkey/HSGM-302/2020,Turkey/HSGM-1428/2020 | - |
| Nurtop et al | 1 | Turkey/KOC-IST-B316/2020 | - |
| Ilker Karacan et al | 3 | Turkey/GLAB-CoV155/2020,Turkey/GLAB-CoV195/2020,Turkey/GLAB-CoV176/2020 | - |
| The Lighthouse Lab in Milton Keynes et al | 1 | England/MILK-826585/2020 | - |
| Krasnov et al | 1 | Russia/SAM-RARI-1011/2020 | - |
| Jean-Jacques Muyembe Tamfum et al | 1 | DRC/D09-0700/2020 | - |
| Brendan L. O'Connell et al | 4 | USA/OR-OHSU-2477/2020,USA/OR-OHSU-2449/2020,USA/OR-OHSU-2859/2020,USA/OR-OHSU-2553/2020 | - |
| Darren L Smith et al | 2 | England/NORT-1B726CD/2020,England/NORT-1B63243/2020 | - |
| Đỗ Thái Hùng et al | 1 | Vietnam/22/2020 | - |
| Gabriela Sevillano Camilo Zurita-Salinas Karen Loaiza David Ortega-Paredes Jeannete Zurita et al | 1 | Ecuador/ZZL-4/2020 | - |
| Galia Zaide et al | 1 | Israel/ISR_NH_M2_0614/2020 | - |
| Al-Jawabreh et al | 2 | Palestine/AAS15/2020,Palestine/AAS16/2020 | - |
| Farhan Ali et al | 1 | India/KA-InStem-NCBS-0042/2020 | - |

| | | | |
|---|---|---|---|
| AlAbbas et al | 2 | Bahrain/920457374_S4/2020,Bahrain/340862225_S35/2020 | - |
| Kei Miyakawa et al | 1 | Japan/YCU02/2020 | - |
| Francesco Messina et al | 2 | Italy/LAZ-INMI-50/2020,Italy/LAZ-INMI-42/2020 | - |
| Cesare E.M. Gruber et al | 3 | Italy/LAZ-INMI-40/2020,Italy/LAZ-INMI-37/2020,Italy/LAZ-INMI-27/2020 | - |
| Mustafa HASOKSUZ et al | 2 | Turkey/Pen07-2/2020,Turkey/Pen07/2020 | - |
| Sandra Carbo et al | 1 | Spain/VC-FISABIO-597/2020 | - |
| Lương Chấn Quang et al | 1 | Vietnam/PIHCM-638/2020 | - |
| Keith Durkin et al | 2 | Belgium/ULG-10072/2020,Belgium/ULG-10082/2020 | A phylodynamic workflow to rapidly gain insights into the dispersal history and dynamics of SARS-CoV-2 lineages |
| Tanya Golubchik et al | 1 | England/OXON-AEFF8/2020 | Preliminary analysis of SARS-CoV-2 importation & establishment of UK transmission lineages |
| Wenjie Tan et al | 1 | Beijing/IVDC-01-06/2020 | - |
| Annika Nilsson et al | 1 | Sweden/20-07237/2020 | - |
| Piras Giovanna et al | 1 | Italy/SAR-ASSL-02/2020 | - |
| Yin Chen et al | 1 | Zhejiang/SX0715/2020 | - |
| Huilai Ma et al | 6 | env/Qingdao/IVDC-04-10/2020,env/Qingdao/IVDC-09-10/2020,env/Qingdao/IVDC-07-10/2020,env/Qingdao/IVDC-08-10/2020,env/Qingdao/IVDC-010-10/2020,env/Qingdao/IVDC-011-10/2020 | - |
| Lovisa Hjerten et al | 1 | Sweden/20-14094/2020 | - |
| João Costa et al | 1 | Portugal/IGC3484/2020 | - |
| Cathy Paulino et al | 1 | Portugal/IGC4024/2020 | - |
| Joao Sobral et al | 3 | Portugal/IGC4103/2020,Portugal/IGC4251/2020,Portugal/IGC4479/2020 | - |
| Alexander Nagy et al | 1 | CzechRepublic/2308/2020 | - |
| Lukasz Rabalski et al | 5 | Poland/Sla6/2020,Poland/Sla1/2020,Poland/Sla4/2020,Poland/Sla8/2020,Poland/Sla5/2020 | - |
| Leanne Mortimer et al | 1 | Canada/ON-E62/2020 | - |
| Benoit et al | 2 | Canada/UN-CRCHUM-PreFreeze-2/2020,Canada/UN-CRCHUM-PostFreeze-2/2020 | - |
| Alexa K Dowdell et al | 1 | USA/OR-PROV-122/2020 | Genomic heterogeneity and clinical characterization of SARS-CoV-2 in Oregon |
| Anik Budhi Dharmayanthi et al | 1 | Indonesia/JK-LIPI002/2020 | - |
| Kowalski et al | 5 | Poland/PL_MCB_80/2020,Poland/PL_MCB_82/2020,Poland/PL_MCB_83/2020,Poland/PL_MCB_79/2020,Poland/PL_MCB_110/2020 | - |
| Olga Douvropoulou et al | 2 | SaudiArabia/KAUST-MAKKAH1426/2020,SaudiArabia/KAUST-MAKKAH1427/2020 | - |
| Md. Saddam Hossain et al | 1 | Bangladesh/BCSIR-NILMRC-233/2020 | - |
| Md. Ahasan Habib et al | 1 | Bangladesh/BCSIR-NILMRC-154/2020 | - |
| Azzania Fibriani et al | 1 | Indonesia/JB-TFRIC19-R48562/2020 | - |
| Susan Engelbrecht et al | 3 | SouthAfrica/Tygerberg_360_CC2/2020,SouthAfrica/Tygerberg_362_CC2/2020,SouthAfrica/Tygerberg_358_CC2/2020 | - |

Supplemental File 3

All submitters of data may be contacted directly via www.gisaid.org

Shu Y., McCauley, J. (2017) GISAID: from vision to reality EuroSurveillance 22(13) doi:10.2807/1560-7917.ES.2017.22.13.30494 PMCID: PMC5388101
